# Climate change, hygiene, and health: a research roadmap for climate adaptation

**DOI:** 10.1101/2025.08.25.25334069

**Authors:** Jessica Gerard, Lauren D’Mello-Guyett, Jane Falconer, Camille Heylen, Sari Kovats, Robert Dreibelbis

## Abstract

Climate change is intensifying hazards that threaten population health, including those that undermine hygiene behaviours, services, and infrastructure. While water and sanitation feature in climate–health agendas, hygiene, and the behaviours and enabling conditions that interrupt disease transmission, remains under-specified. We identified critical research priorities at this intersection through a structured, consensus-driven process involving global experts. Using the Child Health and Nutrition Research Initiative (CHNRI) method, we generated potential research questions via a rapid scoping review and key informant interviews. After screening and refinement, 57 questions were classified using the “4Ds” framework (description, delivery, development, discovery) and scored by global stakeholders against four weighted criteria: relevancy, answerability, potential impact, and potential for translation. The multilingual survey was disseminated globally and completed by 141 respondents from 40 countries representing academic, non-governmental, multilateral, and government sectors.

The top 20 research priorities spanned all aspects of hygiene and climate-related hazards and impact drivers, with 75% focussing on descriptive research to quantify disease burdens, understand hygiene practices and coping strategies under climate stress. Climate-related hazards, such as extreme weather events, dominated the questions (55%) with water scarcity as the second most frequently cited climate impact driver. Delivery-focused priorities (20%) addressed preparedness strategies, targeted support for vulnerable groups, and hygiene promotion during hazards. Only one top-ranked question related to intervention development. Equity considerations, particularly for people with disabilities, women, and girls, featured prominently.

This research agenda addresses a critical gap in climate–health priorities by explicitly linking climate hazards to hygiene practices, infrastructure, and behaviour change needs. By directing attention and resources to these priorities, we aim to guide future research investments, inform policy, and support programming responsive to climate-driven hygiene challenges. The agenda complements global initiatives on climate and health, emphasising the need for evidence-based strategies to safeguard health in an increasingly unstable climate.

## Introduction

Hygiene encompasses a diverse set of behaviours aimed at reducing the risk of disease transmission, spanning domestic and institutional settings. Modifying these behaviours can reduce the multiple exposure routes important for disease transmission through hands, food, and surfaces. Inadequate hygiene remains a major contributor to the global burden of disease: poor hand hygiene was linked with nearly three-quarters of a million deaths from diarrheal disease and acute respiratory infections in 2019 (1). Inadequate food hygiene is a major source of the estimated 600 million cases of foodborne illness and 420,000 deaths in 2010, the last year for which global estimates are available (2). Studies in healthcare and public settings have shown how specific pathogens move through environments via high-touch surfaces and a lack of robust hygiene protocols (3).

Climate change poses an escalating threat to population health. Over half of known human pathogenic diseases are aggravated by climatic factors, due to the alteration of behaviour, range, and longevity of vectors, hosts and reservoirs, and increased propagation of viruses, bacteria, and protozoa (4, 5). As these disease pressures rise, effective hygiene practices become increasingly critical to reduce infectious and chemical risks (6).

However, the same climatic changes that heighten the need for hygiene also undermine the infrastructure, resources and services that enable it. Slow-onset events, such as drought and land degradation, and acute shocks, such as floods and cyclones, are undermining water and sanitation systems and disrupting critical supply chains (7–12). Climate change is reducing overall water availability, for example, through the reduction of glaciers and snowpack that are crucial for freshwater supply (13). Seasonal and annual rainfall patterns are becoming less predictable, contributing to household water scarcity and heightened perceptions of insecurity that drive changes in household water use (14–16). Without reliable access to water and sanitation services or hygiene products, the likelihood of unsafe hygiene behaviours and disease exposure increases, further exacerbating the health risks posed by climate change (1, 12).

Climate hazards threaten all aspects of hygiene. Shifts in water resources and service reliability can trigger coping mechanisms that have direct implications for hygiene and health. Reduced water availability often forces households to prioritise water use for drinking and cooking over hygiene needs, reducing handwashing, bathing, or cleaning (17–23). Households may also use more contaminated sources for washing, cleaning, or food preparation (24–26). Water scarcity can decrease the frequency of handwashing and bathing, heightening risks of enteric and dermatological infections (19, 27, 28). In coastal areas, saltwater intrusion further limits freshwater availability, increasing reliance on brackish water for household cleaning and bathing (29, 30)

Alongside these slow-onset challenges, extreme weather events, such as floods and storms, can also reduce household access to water by damaging infrastructure. This can drive behaviours such as reduced hygiene and washing or laundering in contaminated waters (31, 32). Menstruating women and girls face specific challenges during disasters due to compounded barriers in accessing water and sanitation services or hygiene products (33–35). People with disabilities may be at greater risk because of existing vulnerabilities and limited ability to express their hygiene needs or access safety information (36, 37). Floods and cyclones can also destroy or overwhelm sanitation facilities, leading to increased open defecation or faecal contamination of the home environment (38, 39). Households are also exposed to pathogens and chemical pollutants through contamination of surfaces and stagnant water left behind from floods (40–43).

Food hygiene is similarly threatened. Changes in ambient temperature, precipitation, and humidity affect food safety during storage and preparation, facilitating the growth of and spread of foodborne pathogens and toxins (44, 45). Ocean warming, climate-driven acidification, and shifts in salinity and precipitation alter the biochemical properties of water, impacting microflora and the persistence and patterns of pathogenic bacteria, harmful algal blooms, and chemical contaminants in foods (46–48). Extreme weather events can damage or contaminate household food storage facilities, increasing the risk of cross-contamination and outbreaks of foodborne illness.

Research on the role of climate determinants in population health is increasing (49, 50), supported by national adaptation plans and strategic research agendas that emphasise actions to reduce the impacts of climate change (51, 52). Recent frameworks, including the National Academies of Medicine’s research agenda (53), the European Commission’s Strategic Research and Innovation Agenda on Health and Climate Change (54), and the National Institute of Health’s Climate Change and Health Initiative (55), identify priority domains such as health impacts, adaptation and mitigation, infrastructure and capacity, and evidence-to-policy translation. The World Health Organization’s (WHO) latest global research agenda on climate change and health similarly highlights these domains, with a strong emphasis on equity and action priorities. While water and sanitation services are recognised as a critical component across these agendas, hygiene, encompassing behaviours, services and enabling determinants, remains comparatively under-specified (56).

Hygiene does feature in a limited number of climate adaptation strategies and research agendas. In 2024, WHO and UNICEF released a joint review identifying new indicators to enhance national and global monitoring of climate-resilient WASH, including monitoring of hygiene-related infrastructure and behavioural elements (57, 58). Similarly, guidance on climate-resilient WASH from UNICEF acknowledges the need to change behaviours as well as address infrastructure (59). In Europe, there are new frameworks assessing the emerging climate-related risks to food safety, including fungal toxins and seafood contamination, and identifying vulnerabilities in the microbiological safety of food under scenarios of extreme weather, supply chain disruption and changing production practice (60). Meanwhile, WHO’s hand hygiene research agenda for 2023-2030 and related initiatives mention the potential of behaviour change to enhance hygiene under climate stress in both domestic and institutional settings (61–63).

Understanding the interplay between climate hazards, hygiene practices, and health outcomes is challenging due to the complex, multi-sectoral, and context-specific pathways involved. Factors such as social norms, poor infrastructure resilience, service delivery models, and the lack of prioritisation of adaptation undermine action. This highlights why dedicated research is urgently needed to move beyond acknowledging the problem to identifying specific, evidence-based priorities for action.

This study sought to identify consensus-based research priorities at the intersection of climate change, hygiene, health, and well-being. By systematically identifying and ranking key research gaps through global expert engagement, the aim is to guide future research investments, inform policy and support programming that is responsive to the climate-driven hygiene challenges faced by diverse communities. These priorities are intended to ensure that research efforts align with the needs articulated by national and local practitioners, policymakers and researchers working across disciplines and sectors.

## Methods

To define research priorities on climate change, hygiene, and health, we used a consultative approach based on the Child Health and Nutrition Research Initiative (CHNRI) method to identify research questions and prioritise them in a transparent, consultative, comprehensive and replicable manner (64). The CHNRI methodology has been used previously in prioritising health research investments. We combined the full CHNRI process into three steps: question identification, question refinement, and scoring and prioritisation. The entire process was overseen by the study team, four individuals who represent a diversity of expertise in areas of climate impacts, adaptation, and hygiene (LDG, JG, SK, RD).

### Step 1. Question identification

The scope of the research questions and research prioritisation exercise was agreed upon by the study team at the outset (**Table 1**).

**Table 1.** Scope of the research prioritisation exercise.

| Table 1. Scope of the research prioritisation exercise |  |
| --- | --- |
| Area | Scope |
| Target population | All global regions are experiencing climate hazards, recognising that while exposure is universal, research should prioritise communities with the greatest vulnerability and lowest adaptive capacity. |
| Geographical scope | Global |
| Time horizon | Questions should be answerable by 2035, capturing both short-term, medium-term, and longer-term evidence needs |
| Outcomes of interest | Health outcomes (e.g., disease burdens, morbidity and mortality, wellbeing, and quality of life); behavioural outcomes (e.g., hygiene practices, social norms, coping strategies); human rights-based outcomes (e.g., right to adequate hygiene, wellbeing, dignity, privacy); and inclusive health outcomes (e.g., consideration of people with disabilities, women/ girls, older adults). |

An initial set of potential questions was identified through a rapid scoping review and key informant interviews. Full details on these methods are provided in Supplemental Material 1 (Tables 1 and 2). A total of 116 peer-reviewed manuscripts and 55 key informant interviews formed this initial dataset. The study team reviewed manuscripts and interview transcripts and extracted relevant research questions related to any combination of the intersection of climate change, hygiene, health, and well-being.

**Table 2.** Top 20 research priorities on climate change, hygiene, and health.

| Rank # | Climate Change Hazard Category | 4Ds framework | Research question | n | Weighted RPS (%) | Scaled RPS (%) |
| --- | --- | --- | --- | --- | --- | --- |
| 1 | Climate-related hazards | Delivery | What hygiene promotion actions and preparedness strategies are needed before or during extreme weather events to prepare populations for increased risks to health? | 117 | 75.4% | 100.0% |
| 2 | Climate impact drivers | Description | How does climate-induced water scarcity, drought or precipitation variability affect the infectious disease burden, and how is this mediated by changes to hygiene practices? | 120 | 74.7% | 96.5% |
| 3 | Climate impact drivers | Description | How do climate-induced water scarcity and drought affect an individual's use and consumption of water for hygiene practices? | 122 | 74.4% | 95.1% |
| 4 | Climate impact drivers | Description | What is the association between higher ambient temperatures, precipitation variability and humidity with the incidence of diarrhoeal diseases, including cholera? | 121 | 74.0% | 93.3% |
| 5 | Climate-related hazards | Description | During extreme weather events, how does solid waste contaminate the domestic environment and increase exposure to infectious diseases and/or chemical pollutants? | 129 | 73.5% | 90.6% |
| 6 | Climate-related hazards | Delivery | What support is needed for people with disabilities and their caregivers during extreme weather events to maintain personal hygiene? | 115 | 73.1% | 89.0% |
| 7 | Climate impact drivers | Development | Are different approaches needed for hygiene promotion and hygiene behaviour change for populations living in water-scarce or drought conditions? | 122 | 72.4% | 85.1% |
| 8 | Climate-related hazards | Description | How do extreme weather events affect the capability of people with disabilities to maintain personal hygiene? | 123 | 72.0% | 83.3% |
| 9 | Climate impact drivers | Description | Does climate-induced water scarcity or drought affect an individual's capability, opportunity, or motivation to practice effective hygiene behaviours? | 122 | 72.0% | 83.2% |
| 10 | Climate impact drivers | Description | How do extreme heat events affect an individual's use and consumption of water for hygiene practices? | 126 | 71.9% | 83.0% |
| 11 | Climate-related hazards | Delivery | Given the microbial and chemical contamination risks of floodwater, what messages should be included in health promotion and hygiene behaviour change campaigns? | 129 | 71.7% | 82.0% |
| 12 | Climate impact drivers | Description | What effect will an increase in ambient temperature and humidity associated with climate change have on pathogen levels, with implications for human health, in water, food, soil, surfaces, and the environment? | 121 | 71.6% | 81.5% |
| 13 | Climate-related hazards | Description | What are the determinants of hygiene behaviours during extreme weather events? | 112 | 71.6% | 81.3% |
| 14 | Climate-related hazards | Description | What challenges are faced by women and girls concerning menstrual health and hygiene during extreme weather events, periods of climate-induced water scarcity, drought, and saltwater intrusion? | 119 | 71.3% | 80.1% |
| 15 | Climate-related hazards | Description | What is the association between extreme weather events, climate-induced water scarcity, drought, or saltwater intrusion with the incidence of neglected tropical diseases (NTDs), and how are these risks mediated by hygiene practices? | 119 | 71.2% | 79.6% |
| 16 | Climate-related | Description | How does chronic disruption of water and sanitation services due to extreme weather | 123 | 70.9% | 77.8% |
|  | hazards |  | events impact personal hygiene behaviours? |  |  |  |
| **17** | Climate impact drivers | Description | How will extreme heat affect water collection practices and the water available for hygiene practices? | 126 | 70.8% | 77.5% |
| **18** | Climate-related hazards | Description | What challenges are there in maintaining adequate personal hygiene during extreme weather events, periods of climate-induced water scarcity, drought, and saltwater intrusion? | 119 | 70.6% | 76.4% |
| **19** | Climate-related hazards | Description | How does acute disruption of water and sanitation services due to extreme weather events impact personal hygiene behaviours? | 123 | 70.5% | 75.9% |
| **20** | Climate impact drivers | Delivery | Given the food contamination risks resulting from higher temperatures and extreme weather events, what messages should be included in the promotion of safe food hygiene practices? | 126 | 70.3% | 74.8% |

The study team screened, de-duplicated and refined questions to ensure relevance to study aims. Questions were organised to address climate impact drivers and specific climate hazards following the Intergovernmental Panel on Climate Change (IPCC) framework: (Figure 1). Climate impact drivers include changing precipitation patterns, prolonged dry spells or intensified rainfall, sea level rise, which contributes to inundation, coastal change, and increased salinisation, rising average temperatures, reduced soil moisture, and increased atmospheric humidity. Climate-related hazards are specific weather or climate events, and include hydrometeorological hazards (floods, tropical cyclones, heavy rainfall events); temperature extremes (heatwaves and cold waves), and compound events, like concurrent drought (65–67).

**Figure 1.**
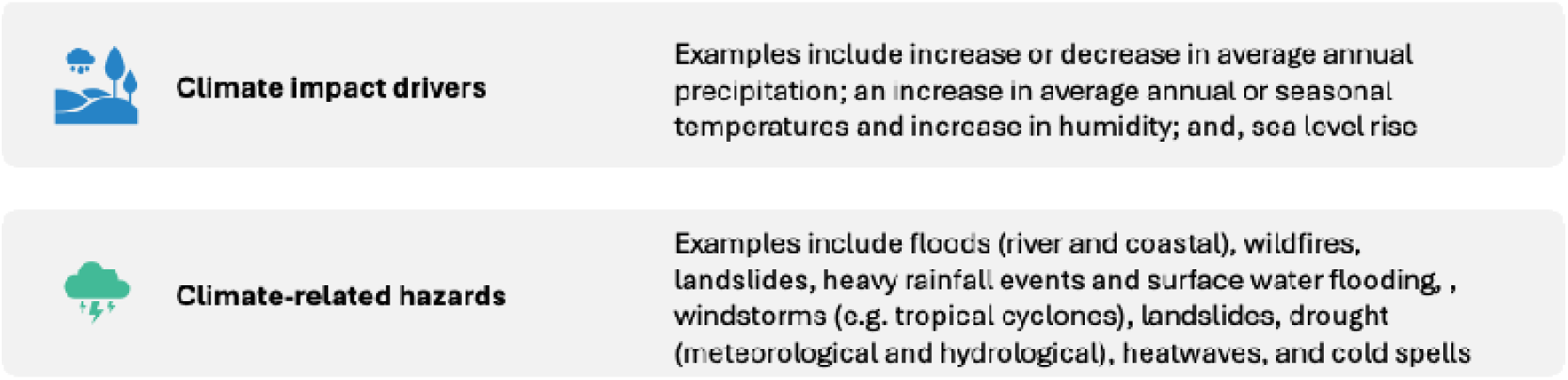
Definitions and examples of climate impact drivers and climate-related hazards, adapted from the Intergovernmental Panel on Climate Change (IPCC) framework.

We further organised the research questions by three broad hygiene domains (68, 69), drawing on definitions of behaviour from the Capability, Opportunity and Motivation Behaviour (COM-B) model (70). We defined *personal hygiene* as behaviours focused on maintaining the cleanliness of the body to support health and well-being. Examples include bathing, handwashing, toilet use, menstrual hygiene, hygiene material access, and incontinence care. We defined *domestic hygiene* as behaviours that clean and sanitise the household and peri-domestic environment. Examples include washing clothes and bedding; cleaning the toilet, floor, and surfaces; water storage practices; domestic animal, livestock, and vector habitat management; post-flood cleaning; and managing solid waste. We defined *food hygiene* as behaviours related to safe food handling. This includes food storage, food preparation and washing, cross-contamination prevention in the home, and ensuring food safety from preparation to consumption.

### Step 2. Question refinement

Question identification (Step 1) resulted in a total of 141 research questions. These questions were reviewed by six technical advisors from academic institutions (see Acknowledgements). The advisors assessed the questions for relevance to climate change, hygiene, and health; checked for duplication; and provided editorial feedback. This review process refined and consolidated the list to 57 questions.

The study team classified these 57 questions according to definitions from the “4Ds framework” of description, delivery, development, or discovery (see Supplementary Material 1, Table 4) (71). *Description* includes research to assess the existing situation. *Delivery* refers to research to evaluate existing interventions. *Development* describes research to improve or develop existing interventions. *Discovery* includes research that may lead to innovations and new interventions. This categorisation ensured that the final set of research questions not only aligned with the climate and hygiene scope established in Step 1 but also reflected expert judgement on the translational pipeline of the intended research.

### Step 3: Scoring and prioritisation

To allow a ranking of the 57 questions refined (Step 2), we first established the criteria for scoring. The original CHNRI methodology includes 15 possible criteria for scoring research questions (see Supplementary Material 1, Table 3) (72). These were shared with four members of the technical advisory group, who were then asked to allocate 100 points across the criteria based on their perceived importance. Weights were calculated by dividing the mean points allocated by 25, yielding relative weights for each criterion. The four highest-scoring criteria, which served as the basis of subsequent scoring, were:

- *Relevancy (criteria weight= 0.88)*: Will the research answer relevant evidence gaps?
- *Answerability (criteria weight= 0.84):* Can the research be feasibly completed and answered between now and 2035?
- *Impact (criteria weight= 0.80):* Is the proposed research likely to improve population health and well-being?
- *Potential for translation* (*criteria weight=* 0.80): Will the research generate knowledge that can be translated into feasible interventions?

An online survey was developed using Qualtrics (Provo, Utah, USA). A targeted dissemination approach was taken, sharing the survey through existing mailing lists, professional networks, participant resharing and social media platforms were used to make the survey widely available.

Respondents were asked to score each question by the four priority criteria - relevancy, answerability, impact, and potential for translation. They indicated their assessment by selecting “Yes” (allocated 1 point), “Maybe” (0.5 points), “No” (0 points), or “Not my Area of Expertise” (excluded from the scoring). The survey was made available in Arabic, English, French, Portuguese, and Spanish, and was open between November 2024 and January 2025. Additional information was collected on respondent gender, organisation affiliation, country, and region (according to WHO classification); the geographic level and focus of their work; years of experience in climate change and/or hygiene or water or sanitation; and areas of expertise. Finally, respondents were invited to suggest thematic areas they felt were missing or needed more specific questions, which are listed in Supplementary Material 2, Table 1

For each research question, the research priority score (RPS) was calculated (see Supplementary Material 1, Table 5 for details). Scores were converted into a scaled and weighted RPS ranging from 0 to 100%. To support decision-making at different levels, results were also stratified by WHO region and by organisation affiliation, allowing us to explore differences in research priorities within different contexts.

### Ethical approval

Ethical approval was received from the LSHTM Research Ethics Committee (No. 30443) for the survey and interviews. Before KIIs, respondents received a respondent information sheet and signed informed consent forms, which outlined confidentiality and risk of the procedure, as well as the voluntary nature of participation and that data would be anonymised before analysis. Survey respondents and all answers were anonymous, as no personal identifiers were required from respondents. However, survey respondents were asked at the end of the survey if they would like to receive further updates on the project by sharing a professional email address.

## Results

A total of 141 people from 40 countries completed the online survey, representing a diverse cross-section of stakeholders, including academic institutions, international and local non-governmental organisations (NGOs), multilateral agencies, and government bodies, and reflecting a broad set of geographic and disciplinary perspectives (see Supplementary Material 2, Table 2).

The top 20 highest-ranked research questions, shown in **Table 2**, achieved consistently high weighted RPS ranging from 75–100%, indicating high consensus on their importance. These questions spanned the hygiene domains and reflected a range of climate-impact drivers and hazards, including water scarcity, extreme weather events, and rising temperatures. The rank order list of all 57 research questions and questions stratified by organisational affiliation or region of focus can be found in the Supplementary Materials 3 and 4.

The top-ranked questions revealed a clear emphasis on building the foundational evidence base at the intersection of climate change, hygiene, and health. A substantial majority (75%) of the top 20 questions focused on *descriptive* research, aiming to better characterise the burden of infectious diseases linked to climate-related hazards, understand existing hygiene practices and coping strategies, and identify barriers to maintaining safe hygiene under climate stress. Examples include exploring how household water scarcity, drought, and precipitation variability affect infectious disease burdens through impacts on hygiene behaviours (Q2, Q3, Q9, Q15) and assessing how extreme weather events disrupt hygiene practices (Q14, Q18) or the capability of people with disabilities to maintain personal hygiene (Q8). Other descriptive questions addressed contamination pathways, such as the role of solid waste in spreading pathogens and pollutants during floods (Q5), and the determinants of hygiene behaviours in the context of extreme weather events (Q13, Q16, Q19).

One fifth (20%) of the top-ranked questions related to *delivery*, focusing on how to provide practical support measures and preparedness messaging to protect health during climate hazards. Examples included identifying effective hygiene promotion actions before or during extreme weather events (Q1, Q11), tailoring support for people with disabilities (Q6), and developing communication strategies to promote safe hygiene during higher temperatures (Q10) and extreme weather events (Q20). Only a single (5%) question fell under *development*, exploring how to adapt or refine existing hygiene interventions (Q7). This included assessing whether tailored approaches are needed for hygiene promotion in water-scarce or drought-prone settings.

Across the top 20, climate hazards were a dominant focus, appearing in 11 questions. Many sought to understand immediate disruptions from floods, cyclones, and other extreme weather events on hygiene behaviours; the cascading and long-term impacts from damaged infrastructure; and the impact of chronic service disruptions (Q5, Q11, Q16, Q19). Reduced rainfall and water availability emerged as the most prominent climate impact driver of interest, while there were relatively few prioritised questions focusing on high temperatures or heavy rainfall as direct hygiene-related health risks (Q2, Q7). However, some priorities addressed heat- and humidity-related effects, including their influence on water use for hygiene (Q10, Q17) and on pathogen dynamics across water, food, and domestic surfaces (Q4, Q12, Q20).

## Discussion

This work complements and strengthens other global initiatives to identify evidence needs on the links between climate and health. By explicitly focusing on hygiene, a domain often underrepresented in climate-health discussions, this exercise fills a critical gap and provides actionable priorities to guide investments and research strategies over the next decade. By applying a structured, consensus-driven CHNRI process, our study has identified these priorities through a transparent and credible process that relies on extensive engagement (72). Taken together, this prioritisation provides a clear, stakeholder-informed roadmap for funders, researchers, and policymakers seeking to support the most critical research needs at this intersection (73). By directing attention and resources to these priorities, the sector can build the evidence base needed to inform adaptive strategies, strengthen hygiene promotion, and safeguard health against the accelerating impacts of climate change.

The findings underline the importance of multidisciplinary and cross-sector collaboration (74). Climate change, hygiene, and health are interlinked through complex pathways that span environmental systems, infrastructure, human behaviour, and social determinants. Addressing the identified research gaps will require integrating expertise across public health, water and sanitation service delivery, climate science, behavioural research, and policy. Such collaboration is essential to design resilient interventions that are both scientifically grounded and practically feasible, ensuring that adaptation efforts meaningfully protect population health in an increasingly unstable climate (75).

The strong leaning toward descriptive research underscores that the field remains at a stage where robust empirical data on exposure pathways, disease burdens, and behavioural responses are urgently needed (76, 77). This foundational understanding appears to be viewed by experts as essential before effective interventions and large-scale programmatic adjustments can be designed and tested. The relatively smaller number of delivery and development-focused questions suggests that while there is recognition of the need to adapt hygiene strategies to climate realities, there is still limited evidence to inform how best to operationalise these adaptations. However, the presence of high-ranking delivery questions, such as those on preparedness strategies and messaging during extreme weather, points to an emerging readiness to move toward implementation research in some areas (78).

Equity concerns were evident across the priorities. Several top-ranked questions explicitly addressed the compounded vulnerabilities faced by people with disabilities, women and girls managing menstrual hygiene, and communities confronting multiple overlapping climate hazards. This aligns with broader calls in climate and health research to centre the experiences of groups who disproportionately bear the impacts of climate change and to design interventions that do not exacerbate existing inequalities (37).

Consistent with patterns observed in other climate-health research agendas (49, 50, 53–55), our findings show a stronger priority given to climate-related hazards, such as extreme weather events, than on slow-onset changes such as chronic heat exposure or progressive water salinisation. In our prioritisation, 11 of the top 20 questions addressed the impacts of floods, cyclones, and storms on hygiene behaviours, infrastructure, and domestic environments. These events are highly visible, cause acute disruption, and frequently trigger disaster response, making them more salient to policymakers, the media, and the public than gradual climatic shifts (79). Similarly, much of the climate–health evidence base focuses on water-related pathways (e.g., floods, droughts, water scarcity) because these have direct, well-recognised links to health outcomes and can be more readily measured and modelled than other drivers such as land degradation, biodiversity loss, or shifting disease ecologies (50, 76). Water insecurity was another dominant theme in our results, with reduced rainfall and drought highly ranked research priorities, often linked to changes in water use for hygiene and consequent impacts on disease burden. The less visible, slow-onset hazards and their long-term implications for hygiene, however, remain important, and our findings demonstrate the need to expand research and intervention portfolios to address the full spectrum of climate change-related risks.

The research questions that emerged from this process are necessarily broad and require further specification and contextualization before they are readily translated into research protocols. It is important to note that our research prioritisation process did not differentiate between research questions that reflect a lack of data or information around a specific topic and questions that reflect a lack of knowledge about a specific evidence base. For example, the links between climate hazards and infectious disease burden were the second-highest prioritised research question in our exercise. There are multiple reviews, both estimating climate-related infectious disease burden (80) and infectious disease transmission pathways related to climate change (81), but these studies may not explicitly include hygiene in their discussion of potential transmission pathways or prevention strategies. Addressing research questions related to climate, hygiene, and health will require structured engagement with existing scientific evidence to extract relevant insights and findings. Changing water use practices was our third highest-ranked research question. Multiple studies describe context-specific individual and household changes in water use and water prioritisation in the context of water scarcity (82–84), and the research gaps may reflect a lack of evidence synthesis. There is a wide range of evidence synthesis methods (85) that should be explored for their potential relevance to addressing the stated research priorities.

### Integration with global policy and existing literature

These findings resonate with, but also extend, the growing international focus on climate change and health. This research agenda helps fill this key gap by providing explicit priorities that link climate hazards to hygiene practices, infrastructure, and behaviour change needs. It also complements broader calls for climate-resilient health systems by operationalising what this means in the context of everyday hygiene (86), a domain critical for preventing infectious diseases but often absent from high-level adaptation plans (87).

Moreover, by highlighting equity concerns, particularly the compounded risks for women, girls, people with disabilities, and those facing multiple hazards, these priorities align closely with international commitments such as the Sustainable Development Goals (SDGs), which emphasise leaving no one behind (88), and the Sendai Framework for Disaster Risk Reduction (DRR) (89), which calls for inclusive approaches to managing risk. As climate hazards intensify, this targeted roadmap provides a timely contribution to guide research investments that can underpin both policy and practice, ensuring that hygiene is not overlooked in the global climate-health dialogue.

#### Limitations

This study has several limitations related to both the CHNRI methodology and the complexity of assessing the intersection between climate change, hygiene, and health. A key methodological challenge was the potential difficulty respondents faced in conceptualising hygiene as a distinct area, separate from broader WASH definitions, which may have influenced how research gaps were articulated by key informants and later prioritised.

There are also important considerations related to potential bias. There may be selection bias, as respondents were predominantly individuals already working in hygiene, climate adaptation, or DRR. While this means perspectives reflect those most actively engaged in addressing these challenges, it may also limit input from adjacent sectors or communities not directly involved. However, given the complex linkages between climate, hygiene, and health, having respondents with technical expertise arguably strengthens the relevance and practicality of identified priorities. There is a potential bias associated with the relatively low participation of government representatives, the private sector, and donors. This may narrow the direct applicability of the findings for policy and investment decisions, underlining the need for continued engagement with these groups in future work. Additionally, many respondents had varying familiarity with climatological terms and frameworks, which could have influenced how they interpreted and scored questions related to specific climate drivers and hazards. Lastly, the structured nature of the CHNRI process, while providing transparency and rigour, can sometimes reduce complex, multi-faceted research topics into short-form questions and quantitative scores (90). This approach risks oversimplifying nuanced topics and may dilute some of the interdisciplinary discussions that would emerge in more qualitative approaches.

Despite these limitations, the process engaged a broad, multidisciplinary pool of stakeholders who helped to refine and validate the research questions. This lends credibility to the findings and ensures that the priorities identified are grounded in diverse, practice-based perspectives.

#### Conclusions

This research prioritisation exercise has identified critical knowledge gaps at the intersection of climate change, hygiene, and health, emphasising the urgent need for multi-sectoral action. The findings highlight the necessity of integrating hygiene promotion into climate preparedness, addressing water scarcity impacts on hygiene behaviours, and investigating climate-driven disease transmission pathways. Despite growing recognition of these links, hygiene remains underrepresented in climate change research and policy, underscoring the importance of targeted funding and interdisciplinary collaboration. Without a more comprehensive evidence base, effective public health strategies and climate adaptation efforts will be limited. Strengthening research collaboration across disciplines, geographies, and sectors is essential to ensuring that adaptations and climate change resilience efforts are evidence-based, context-specific, and equitable.

## Data Availability

All data produced in the present work are contained in the manuscript

https://doi.org/10.17037/DATA.00004222

## Acknowledgements

This research agenda was supported by six technical advisors:

- Akanksha Marphatia (London School of Hygiene & Tropical Medicine, UK)
- Belen Torondel-Lopez (London School of Hygiene & Tropical Medicine, UK)
- Julia Sobolik (London School of Hygiene & Tropical Medicine, UK)
- Karen Levy (University of Washington, USA)
- Kondwani Chidziwisano (Malawi University of Business and Applied Sciences (MUBAS), Malawi)
- Oliver Cumming (London School of Hygiene & Tropical Medicine, UK)

The study team would like to acknowledge and thank the survey respondents, interviewees, and all observers who provided time and feedback throughout this process, particularly in identifying, developing, and refining the research questions. Throughout the entire process, numerous experts provided their time and input to shape this research agenda.

## Supporting information

- Supplementary Material 1. Tables 1-5: Detailed CHNRI methodology
- Supplementary Material 2. Tables 1: Additional Research Questions, and Table 2: Demographic Characteristics of Respondents
- Supplementary Material 3. List of all 57 research questions, with research prioritisation scores (RPS)
- Supplementary Material 4. Research priorities stratified by organisational affiliation and region

