## Supplementary Material 1 for "Climate change, hygiene, and health: a research roadmap for climate adaptation"

| **Table 1. Rapid scoping review**  We conducted a rapid scoping review of the literature from 2000 to March 2024 and extracted research questions related to the intersection of climate change, hygiene, and health. Research questions were extracted from a total of 114 published manuscripts from the 11,270 records from a database search. The full search strategy, terms and database search results are hosted in an open-access digital repository maintained by LSHTM, https://doi.org/10.17037/DATA.00004222 (52).  **Search concepts**  The search strategy included strings of terms, synonyms, and controlled vocabulary terms (where available) to reflect the following concepts:  • Concept 1: climate change  • Concept 2: hygiene  • Concept 3: adaptations or perceptions  Terms for the three concepts were combined using the Boolean operator AND to find items discussing some or all concepts.  Terms for concept 1 and concept 2 were initially derived from existing literature and further informed by expert process managers in both subjects. After discussion with the project team, a draft strategy was compiled in the OvidSP Medline database by an experienced information specialist (JF). The search strategy was refined with the project team, with the addition of concept 3, until the results retrieved reflected the scope of the project. The agreed OvidSP Medline search was adapted for each database to incorporate database-specific syntax and controlled vocabularies.  **Limits**  Searches were run with no limits to retrieve the widest range of material possible. However, after the searches were run and duplicates were removed, it was decided to limit the search by date; items published before 2000 were removed. No language restrictions were specified. No geographical limits were specified to make sure items from across the world were included in the review.  **Search strategy and inclusion criteria**  Information sources were chosen to search as wide a selection of sources as possible. Care was taken to choose sources that included locally published titles and a variety of publication types.  **Databases**  The following bibliographic databases were searched on 28 March 2024:  • OvidSP Medline ALL – 1946 to 27 March 2024  • OvidSP Embase Classic + Embase – 1947 to 27 March 2024  • OvidSP Global Health, 1910 to 2024, week 12  • EBSCOhost GreenFILE – complete database to 28 March 2024  Information management  All citations identified by our searches were imported into EndNote 21 software. Duplicates were identified and removed using the method described on the London School of Hygiene & Tropical Medicine Library & Archives Service blog (Falconer, 2018).  Items published before 2000 were removed in EndNote.  Articles were eligible for inclusion if they adhered to the criteria below:   - Date: published during or after 2000 - Language: any language - Types of population/participants: people of all ages from any country - Type of setting: country or areas affected by or at-risk of climate change hazards - Types of studies: all study designs - Types of interventions: not applicable, as studies were not excluded based on the presence or absence of an intervention. | | |
| --- | --- | --- |
| **Results from each database** | | |
| **Database name** | **Total number of results** | **Number of results once duplicates are removed** |
| OvidSP Medline ALL | 5,296 | 5,289 |
| Embase Classic + Embase | 5,197 | 1,936 |
| Global Health | 2,349 | 1,745 |
| GreenFILE | 3,632 | 2,300 |
| **Total** | **16,474** | **11,270** |

| **Table 2. Key informant interviews with key stakeholders**  The study team collectively identified individuals for in-depth interviews who could discuss research questions related to climate change, WASH and global health. People were identified through existing or known research collaborations, recommendations from the study team, and manually searching author lists from the rapid scoping review. Initial respondents included a mix of researchers, governments, non-governmental organisations (NGOs), donor government agencies, private sector companies, and multilateral agencies.  Individuals were invited to participate in a key informant interview (KIIs) between April to September 2024, based on the above eligibility. Interviews followed a broad topic guide on the intersection of climate risks, hygiene, and health. Interviews were conducted over Zoom and lasted for approximately 45-60 minutes. Interviews were recorded and auto-transcribed via the Zoom platform. Respondents were asked to detail any existing research questions related to hygiene, health and climate change, research or programmatic questions within their agency or organisation, and any published, ongoing, or planned research. Respondents were also asked to articulate climate, hygiene, and health research gaps. 146 individuals were invited to participate in a KII, of which 55 responded and participated, resulting in a total of 52 interviews. The interviewees represented diverse sectors, including academia, NGOs, multilateral agencies, the private sector and governments. Data from KIIs was then analysed and used to inform the listing of research questions. | | | | | | | | |
| --- | --- | --- | --- | --- | --- | --- | --- | --- |
| **Characteristics of KII respondents** | | | | | | | | |
| **Organisation type** | **Gender** | **African Region** | **Region of the Americas** | **Eastern Mediterranean Region** | **European Region** | **South-East Asian Region** | **Western Pacific Region** | **Total** |
| Academic | Female | 3 | 2 | – | 5 | 1 | 2 | **13** |
|  | Male | – | 2 | – | 6 | 2 | – | **10** |
| Government | Female | 1 | – | – | – | – | – | **1** |
|  | Male | – | – | – | – | – | – | **0** |
| International non-governmental organisation | Female | – | 1 | – | 6 | 2 | 3 | **12** |
|  | Male | 2 | – | 1 | 4 | – | 1 | **8** |
| Multilateral agency (i.e., United Nations) | Female | – | – | – | 4 | – | 1 | **5** |
|  | Male | 1 | – | – | 2 | 1 | – | **4** |
| Private sector | Female | – | – | – | 2 | – | – | **2** |
|  | Male | – | – | – | – | – | – | **0** |
| **Total** |  | **7** | **5** | **1** | **29** | **6** | **7** | **55** |

**Organisations contacted for KIIs**

Action for Global Health UK

African International University

AMREF Health Africa

Boston University

Bridge Network

CARE International

Canadian International Community Association

Chatham House

Christian Blind Mission (CBM)

CIDRZ (Centre for Infectious Disease Research in Zambia)

CREHPA (Center for Research on Environment Health and Population Activities)

EAWAG (Swiss Federal Institute of Aquatic Science and Technology)

Emory University

European Food Safety Authority (EFSA)

Fiji National University

FINDDx

Geneva Graduate Institute

Global Menstrual Collective

Groupe URD (Urgence, Réhabilitation et Développement)

GTO (German Toilet Organisation)

Hasanuddin University

ICRC (International Committee of the Red Cross)

ICDDR Research Bangladesh (ICDDRB)

ICIMOD (International Centre for Integrated Mountain Development)

IFRC (International Federation of Red Cross and Red Crescent Societies)

Imperial College London

Independent Consultant(s)

International Organization for Migration (IOM)

International Rescue Committee (IRC)

IRC WASH (International Water and Sanitation Centre)

IRSS (Institut de Recherche en Sciences de la Santé)

Johns Hopkins University

London School of Hygiene & Tropical Medicine (LSHTM)

Médecins Sans Frontières (MSF)

Ministère de la Santé (French Ministry of Health)

Monash University

MRC Gambia (Medical Research Council Unit The Gambia)

North Carolina State University (NC State)

Norwegian Church Aid

Norwegian Refugee Council (NRC)

Oxfam

Oxford Policy Management

Queen's University

Reckitt

Red Cross Red Crescent Climate Centre

SAMRC (South African Medical Research Council)

Save the Children International

SIMAD University

Swiss Tropical and Public Health Institute (Swiss TPH)

TDR (Special Programme for Research and Training in Tropical Diseases, WHO)

The Behavioural Insights Team

The George Institute for Global Health

The Leprosy Mission

Tulane University

UCL (University College London)

UK Foreign, Commonwealth & Development Office (UK FCDO)

UK Health Security Agency (UKHSA)

UK Research and Innovation (UKRI)

Universidade Federal da Bahia (UFBA)

University of Bristol

University of Cape Town (UCT)

University of East Anglia

University of Ghana

University of Leeds

University of Manchester

University of Michigan

University of Nebraska–Lincoln (UNL)

University of North Carolina (UNC)

University of Queensland

University of South Carolina

University of Sussex

University of Technology Sydney

University of Washington

UNICEF

UNICEF Pacific

UNICEF WASH Cluster

Unlimit Health

Water for Women Fund

WaterAid

WHO AFRO (WHO Regional Office for Africa)

WHO EURO (WHO Regional Office for Europe)

World Health Organization (WHO)

| **Table 3. Potential prioritisation criteria** | |
| --- | --- |
| **Criterion** | **Definition** |
| 1. Answerability | Some health research options will be more likely to be answerable than others |
| 2. Attractiveness | Some health research options will be more likely to lead to publication in high-impact journals |
| 3. Novelty | Some health research options will be more likely to generate truly novel knowledge that did not exist previously |
| 4. Potential for translation | Some health research options will be more likely to generate knowledge that will be translated into health interventions |
| 5. Effectiveness/ impact | Some health research options will be more likely to generate or improve truly effective health interventions |
| 6. Affordability | Translation or implementation of knowledge generated through some health research options will not be affordable within the context |
| 7. Implementability/ deliverability | Some health research options will lead to or impact health interventions that will not be deliverable within the context |
| 8. Sustainability | Some health research options will lead to or impact health interventions that will not be sustainable within the context |
| 9. Public opinion | Some health research options will seem more justifiable and acceptable to the public than others |
| 10. Ethical aspects | Some health research options will seem more justified and acceptable to the public than others |
| 11. Maximum potential impact | Some health research options will have the theoretical potential to reduce much larger portions of the existing disease burden than others |
| 12. Equity | Some health research options will lead to health interventions that will only be accessible to the privileged in the society or context, thus increasing inequity |
| 13. Community involvement | Some health research options will have additional positive side‑effects through community involvement |
| 14. Cost and feasibility | All other criteria being equal, some research options will still require more funding than others and thus be less feasible investments |
| 15. Likelihood of generating patents/ lucrative products | Some research options will be more likely to generate patents or other potentially lucrative products, thus promising greater financial return on investments, regardless of their impact on disease burden |
| Source: (Rudan, 2008) | |

| **Table 4. The 4Ds framework and example question types for climate change, hygiene and health research prioritisation exercise** | |
| --- | --- |
| Description | Measuring the burden of health and social outcomes |
|  | Understanding the risk factors of health and social outcomes |
|  | Understanding the risk to practices, routines, behaviours |
|  | Measuring prevalence of exposure to risk factors for health and social outcomes |
| Delivery | Evaluating the efficacy of interventions in a laboratory setting |
|  | Evaluating the efficacy and effectiveness of interventions in place |
|  | Evaluating the financial/cost analysis of interventions in place |
|  | Evaluating the provision of infrastructure or system strengthening |
|  | Evaluating human resources or coordination constraints, or requirements |
|  | Evaluating the responsiveness and operational feasibility of interventions in place |
| Development | Improving existing interventions (affordability) |
|  | Improving existing interventions (deliverability) |
|  | Improving existing interventions (effectiveness) |
|  | Improving the responsiveness and operational feasibility of interventions in place |
| Discovery | Basic, clinical and public health research to advance existing knowledge to develop new capacities |
|  | Basic, clinical and public health research to explore entirely novel ideas to develop new capacities |
|  | Basic, clinical, and public health research to explore entirely novel ideas to develop new interventions |

| **Table 5. Research questions organised by the 4Ds framework: description, delivery, development, and discovery** | | | |
| --- | --- | --- | --- |
|  | Questions (n=57) | Questions in the top 20 (n=20) | Proportion in the top 20 research questions (%) |
| Description | 35 | 13 | 65 |
| Delivery | 13 | 2 | 10 |
| Development | 8 | 5 | 25 |
| Discovery | 1 | 0 | 0 |

**Table 5. Calculation of Weighted Research Prioritisation Scores (RPS) and Weighted Average Expert Agreement (AEA) scores**

Respondents were asked to judge whether each of the 58research questions met each criterion by indicating “Yes” (1 point), “Maybe” (0.5 points), “No” (0 points), or “Not my area of expertise” (no input), respectively.

Two scores were calculated for each research question.

**Weighted research priority score**

The following formula was used, where *c* is the criterion used to evaluate the research question; *a*, is the number of criteria selected to prioritise the research question; *W* is the weight for each criterion; and *N* is the number of answers by answer type.

$$s_{Weighted RPS}= \frac{1}{a}\times\sum_{c=1}^{a} W_{c}\times\frac{{(N}_{\mathrm{Yes}}\times1)+{(N}_{\mathrm{Maybe}}\times0.5)}{N_{\mathrm{Yes}}+N_{\mathrm{No}}+N_{\mathrm{Maybe}}}$$

**Weighted average expert agreement scores**

The level of agreement between respondents was then assessed through average expert agreement (AEA), based on the proportion of scorers who gave the most common score (mode) for a question, divided by the total number of scorers who scored that question. This method of validation is unaffected by the varying number of scorers per criterion and differences in scorer composition for the different criteria. In this validation exercise, all four possible responses (Yes, No, Undecided or Insufficiently informed) are treated as valid. Therefore, if a substantial proportion of the experts respond as insufficiently informed, the agreement score will reflect this and reduce the level of agreement rather than increase it.

AEA is the average proportion of scorers who agreed on the five questions asked. The following formula was used, where c is the four criteria used to evaluate the research question and w is the criteria weight developed (step 2):

$$s_{Weighted AEA}=\frac{1}{a}\times\sum_{c=1}^{a} W_{c}\times\frac{N_{Most frequent reponse}}{N_{\mathrm{Yes}}+N_{\mathrm{No}}+N_{\mathrm{Maybe}}{+ N}_{Not my area of expertise}}$$

Weights, which were derived by the initial allocation of 100 points by process managers (step 2), were then applied to both the research priority score (RPS) and AEA. These included: impact 0.80, answerability 0.88, relevancy 0.84, and potential for translation 0.80, respectively.

The weighted scores for AEA and RPS were calculated for each criterion for each research question. As weights were similar across prioritisation criteria, the unweighted RPS or AEA are not presented in this report.

Lastly, the final score (by method of scoring) was converted into a Weighted AEA ranging from 0% to 100%. Based on the final score, a rank was assigned to each research question, where the highest research priority score was ranked 1 and the lowest research priority score 58.
