## Supplementary Material 2 for "Climate change, hygiene, and health: a research roadmap for climate adaptation"

| **Table 1. Topics and associated questions that were potentially missing or could have been elaborated on in the climate change, hygiene, health and wellbeing outcomes research agenda** | |
| --- | --- |
| **Topics** | **Question(s)** |
| Antimicrobial Resistance (AMR) | What are the links between hygiene practices, antimicrobial resistance (AMR) and climate change, and what interventions can reduce risks? |
|  | How does wastewater use and heavy metal contamination in agriculture contribute to antimicrobial resistance (AMR), and what mitigation strategies can be implemented? |
| Behavioural adaptation | The question is not will climate change will make a difference, but how do we help people change and adapt? |
|  | How does climate change make people change their behaviours, as it relates to good hygiene practices? i.e., handwashing with soap |
|  | How do climate change-related hygiene practices impact different age groups within a community? |
|  | How do hygiene priorities change during extreme weather events, e.g., is food hygiene still a priority, but personal hygiene is not? |
|  | How do climate-related disruptions affect the care practices for newborns and infants, including food preparation, diapering, and bathing? |
|  | What is the impact of wildfires on hygiene practices? |
| Displacement, migration and nomadic populations | What strategies can effectively include displaced people in host communities, ensuring access to essential services needed to maintain hygiene, health and wellbeing? |
|  | How do hygiene practices differ between camp settings and host community settings? |
|  | How can climate-affected communities be supported in adapting to forced migration, and accordingly, improving hygiene and health outcomes for climate migrants? |
|  | What are the best practices for promoting hygiene in climate-vulnerable nomadic and pastoralist communities? |
| Engagement, collaboration and coordination (community, stakeholders, etc.) | What are effective strategies for engaging communities and other stakeholders in climate change awareness and action initiatives? |
|  | What are effective strategies for engaging younger populations in climate change awareness and action initiatives? |
|  | What are the most effective ways to improve coordination between sectors (environment, shelter, health, and WASH) to address climate-related health challenges? |
|  | How do governance structures and the political economy influence the integration of hygiene into global health and climate change policies? |
|  | How can government ministries/departments improve coordination (particularly between health and WASH)? |
|  | How do and can health-insurance models or financial support vulnerable populations against climate shocks? |
|  | What are the most effective approaches for costing and financing inclusive, climate-resilient hygiene interventions? |
| Extreme heat | What are the specific health needs of older populations in the context of increasing heatwaves, and how can health systems adapt to better support them? |
|  | What type of cooling stations work best for vulnerable populations, but are inexpensive and easy to build? |
| Food (production, consumption, safety) | What is the impact of wastewater contamination on food crops, particularly in terms of heavy metal accumulation, and how can this be addressed through improved agricultural practices? |
| Health (infectious disease, nutrition, mental health, etc.) | What is the relationship between hygiene practices, climate change, and the emergence of new diseases? |
|  | How do climate-related disruptions affect maternal health? |
|  | What is the relationship between climate change, hygiene and mental health? |
|  | How does climate change impact mental health, particularly in vulnerable populations, and what strategies can support resilience in affected communities? |
|  | How can climate change adaptation strategies utilise domestic hygiene improvements to reduce the burden of vector-borne and other NTDs in vulnerable regions? |
|  | How does climate change impact neonatal and infant health? |
|  | Where can we anticipate the effects of climate change on respiratory disease? |
|  | How do health outcomes and access to care differ between urban and rural areas, particularly regarding the impacts of climate change on vulnerable populations such as older adults?" |
|  | What are the risks of pathogenic (re)contamination of household stored water (used for drinking, cooking, domestic and personal hygiene) under extreme weather events and water-related climate effects? |
|  | What is the relationship between extreme weather events and waterborne diseases? |
|  | How do extreme weather events contribute to child stunting and stress, and what interventions can reduce these impacts in vulnerable populations? |
|  | What is the relationship between the impact of climate change, water/hygiene scarcity and nutrition? |
|  | What are the health and well-being impacts of climate change on female subsistence workers? |
| Infrastructure and technology adaptation | What emerging technologies and infrastructure innovations are most effective in improving hygiene in the context of climate change? |
|  | What are the most effective and scalable waste management strategies for climate-vulnerable communities? |
|  | How can climate adaptation and mitigation help WASH infrastructure? |
|  | How do climate-related disruptions to sanitation infrastructure impact the safe disposal of menstrual hygiene products and diapers, and what resilient waste management strategies can be implemented? |
|  | What are the most effective novel treatments for rainwater and floodwater in improving water quality, particularly in areas affected by climate-related disruptions? |
|  | How can WASH systems be designed for greater resilience to extreme weather events, and what strategies are most effective in ensuring rapid recovery after such events? |
|  | At what level of vulnerability in the areas of water, hygiene and sanitation should the impacts of critical events be attributed to structural conditions rather than to the effects of climate change? |
| Institutional WASH | How does institutional WASH influence health outcomes in the context of climate change? |
|  | How can WASH services in schools and health facilities be improved to better support climate-resilient health outcomes? |
|  | What is the potential of the 'build back better' approach to improve institutional WASH infrastructure and practices after extreme events or floods, particularly in climate-vulnerable areas? |
| Nature-based solutions | What are effective nature-based solutions for enhancing climate resilience? |
| One Health | What is the effect of climate change on animal health, and what impact does it have on the human population? |
| Urbanisation | What are the key challenges and research gaps in improving hygiene in rapidly urbanising areas, and what innovative approaches can be implemented to address these challenges? |

| **Table 2. Demographic characteristics of survey respondents (n=141)** | | |
| --- | --- | --- |
| **Gender** | **n** | **%** |
| Male | 73 | 52 |
| Female | 66 | 47 |
| Non-binary | 0 | 0 |
| Prefer not to say | 1 | 1 |
| **A language survey was taken in** |  |  |
| English | 123 | 87 |
| French | 13 | 9 |
| Arabic | 1 | 1 |
| Spanish | 3 | 2 |
| Portuguese | 1 | 1 |
| **Average experience (years)** | 14 (0–57) | |
| **Region of origin** |  |  |
| African Region (AFR) | 38 | 28 |
| European Region (EUR) | 57 | 41 |
| Eastern Mediterranean Region (EMR) | 6 | 4 |
| Region of the Americas (AMR) | 17 | 12 |
| South-East Asian Region (SEAR) | 13 | 9 |
| Western Pacific Region (WPR) | 7 | 5 |
| Not reported | 0 |  |
| **Region of focus** |  |  |
| African Region (AFR) | 82 | 58 |
| European Region (EUR) | 17 | 12 |
| Eastern Mediterranean Region (EMR) | 21 | 15 |
| Region of the Americas (AMR) | 18 | 13 |
| South-East Asian Region (SEAR) | 42 | 30 |
| Western Pacific Region (WPR) | 15 | 11 |
| Global | 56 | 40 |
| Prefer not to say | 2 | 1.4 |

| **Frequency Survey Respondents Countries of Origin** | | |
| --- | --- | --- |
| **WHO Region** | **Countries** | **Frequency** |
| African Region (AFR) | Angola | 1 |
|  | Burkina Faso | 2 |
|  | Burundi | 1 |
|  | Cameroon | 1 |
|  | Ethiopia | 5 |
|  | Gambia | 1 |
|  | Ghana | 2 |
|  | Kenya | 5 |
|  | Malawi | 1 |
|  | Niger | 1 |
|  | Nigeria | 1 |
|  | Sierra Leone | 2 |
|  | Somalia | 1 |
|  | South Africa | 3 |
|  | South Sudan | 1 |
|  | Sudan | 1 |
|  | Togo | 1 |
|  | Uganda | 5 |
|  | Zambia | 1 |
|  | Zimbabwe | 2 |
| European Region (EUR) | Austria | 2 |
|  | Belgium | 3 |
|  | Finland | 1 |
|  | France | 7 |
|  | Germany | 3 |
|  | Italy | 1 |
|  | Netherlands | 2 |
|  | Sweden | 1 |
|  | Switzerland | 13 |
|  | Ukraine | 1 |
|  | United Kingdom | 23 |
| Eastern Mediterranean Region (EMR) | Afghanistan | 1 |
|  | Jordan | 2 |
|  | Pakistan | 1 |
|  | Yemen | 2 |
| Region of the Americas (AMR) | Brazil | 2 |
|  | Canada | 3 |
|  | Colombia | 1 |
|  | Panama | 1 |
|  | United States of America | 10 |
| South-East Asian Region (SEAR) | Bangladesh | 7 |
|  | India | 1 |
|  | Indonesia | 1 |
|  | Nepal | 4 |
| Western Pacific Region (WPR) | Australia | 6 |
|  | Japan | 1 |

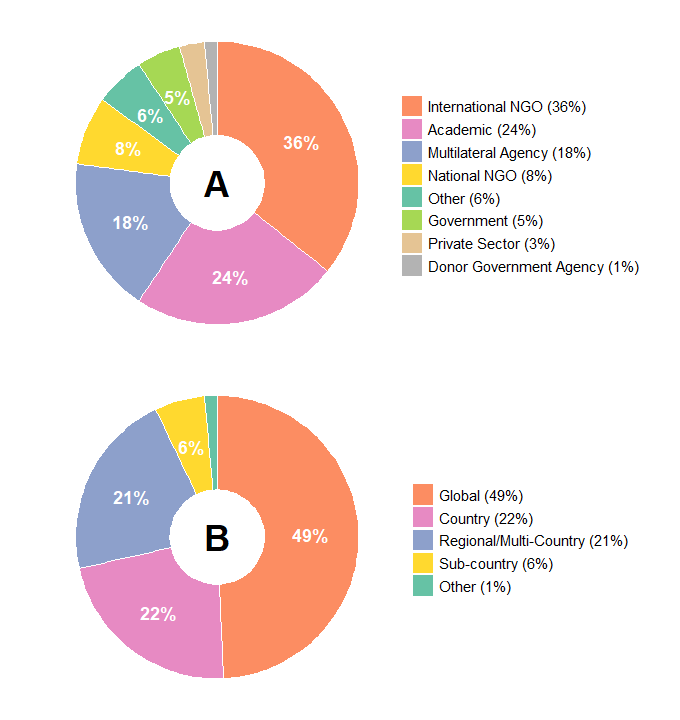

**Figure 2. Characteristics of survey respondents**

Key: (A) organisation type (n=140); (B) geographical level of focus; (C) map with region of focus by World Health Organization (WHO) region
