## Supplementary Material 3 for "Climate change, hygiene, and health: a research roadmap for climate adaptation"

| **Table 1. All 57 questions, in rank order, on climate change, hygiene and health** | | | | | | | | | | |
| --- | --- | --- | --- | --- | --- | --- | --- | --- | --- | --- |
| **#** | **Climate Hazards** | **4Ds category** | **Research question** | **Average number of respondents (n)** | **Scaled Weighted RPS (%)** | **Unscaled Weighted RPS (%)** | **Scaled Unweighted RPS (%)** | **Unscaled Unweighted RPS (%)** | **Scaled Prioritisation score from AEA (%)** | **Unscaled AEA (%)** |
| 1 | Climate-related hazards | Delivery | What hygiene promotion actions and preparedness strategies are needed before or during extreme weather events to prepare populations for increased risks to health? | 117 | 100.0% | 75.4% | 100.0% | 90.9% | 100.0% | 82.5% |
| 2 | Water-related climate drivers | Description | How do climate-induced water scarcity, drought or precipitation variability affect the infectious disease burden, and how is this mediated by changes to hygiene practices? | 120 | 96.5% | 74.7% | 96.3% | 90.0% | 86.3% | 76.7% |
| 3 | Climate-related hazards | Description | How do climate-induced water scarcity and drought affect an individual's use and consumption of water for hygiene practices? | 122 | 95.1% | 74.4% | 94.7% | 89.6% | 95.4% | 80.5% |
| 4 | Temperature-related climate drivers | Description | What is the association between higher ambient temperatures, precipitation variability and humidity with the incidence of diarrhoeal diseases, including cholera? | 121 | 93.3% | 74.0% | 92.8% | 89.2% | 80.9% | 74.4% |
| 5 | Climate-related hazards | Description | During extreme weather events, how does solid waste contaminate the domestic environment and increase the exposure to infectious diseases and/or chemical pollutants? | 129 | 90.6% | 73.5% | 90.4% | 88.6% | 83.7% | 75.6% |
| 6 | Climate-related hazards | Delivery | What support is needed for people with disabilities and their caregivers during extreme weather events to maintain personal hygiene? | 115 | 89.0% | 73.1% | 88.8% | 88.2% | 77.2% | 72.8% |
| 7 | Water-related climate drivers | Development | Are different approaches needed for hygiene promotion and hygiene behaviour change for populations living in water-scarce or drought conditions? | 122 | 85.1% | 72.4% | 84.8% | 87.2% | 85.7% | 76.4% |
| 8 | Climate-related hazards | Description | How do extreme weather events affect the capability of people with disabilities to maintain personal hygiene? | 123 | 83.3% | 72.0% | 83.0% | 86.8% | 77.1% | 72.8% |
| 9 | Climate-related hazards | Description | Does climate-induced water scarcity or drought affect an individual's capability, opportunity, or motivation to practice effective hygiene behaviours? | 122 | 83.2% | 72.0% | 82.9% | 86.7% | 83.3% | 75.4% |
| 10 | Temperature-related climate drivers | Description | How do extreme heat events affect an individual's use and consumption of water for hygiene practices? | 126 | 83.0% | 71.9% | 82.7% | 86.7% | 86.1% | 76.6% |
| 11 | Climate-related hazards | Delivery | Given the microbial and chemical contamination risks of floodwater, what messages should be included in health promotion and hygiene behaviour change campaigns? | 128.5 | 82.0% | 71.7% | 82.0% | 86.5% | 83.9% | 75.7% |
| 12 | Temperature-related climate drivers | Description | What effect will an increase in ambient temperature and humidity associated with climate change have on pathogen levels, with implications for human health, in water, food, soil, surfaces, and the environment? | 121 | 81.5% | 71.6% | 81.5% | 86.4% | 66.3% | 68.2% |
| 13 | Climate-related hazards | Description | What are the determinants of hygiene behaviours during extreme weather events? | 117.75 | 81.3% | 71.6% | 81.1% | 86.3% | 80.7% | 74.3% |
| 14 | Climate-related hazards | Description | What challenges are faced by women and girls concerning menstrual health and hygiene during extreme weather events, periods of climate-induced water scarcity, drought, and saltwater intrusion? | 119 | 80.1% | 71.3% | 79.9% | 86.0% | 64.5% | 67.4% |
| 15 | Climate-related hazards | Description | What is the association between extreme weather events, climate-induced water scarcity, drought, or saltwater intrusion with the incidence of neglected tropical diseases (NTDs), and how are these risks mediated by hygiene practices? | 119 | 79.6% | 71.2% | 79.6% | 85.9% | 48.7% | 60.7% |
| 16 | Climate-related hazards | Description | How does chronic disruption of water and sanitation services due to extreme weather events impact personal hygiene behaviours? | 123 | 77.8% | 70.9% | 77.4% | 85.4% | 80.5% | 74.2% |
| 17 | Climate-related hazards | Description | How will extreme heat affect water collection practices and the water available for hygiene practices? | 126 | 77.5% | 70.8% | 77.5% | 85.4% | 85.7% | 76.4% |
| 18 | Climate-related hazards | Description | What challenges are there in maintaining adequate personal hygiene during extreme weather events, periods of climate-induced water scarcity, drought, and saltwater intrusion? | 119 | 76.4% | 70.6% | 76.3% | 85.1% | 72.4% | 70.8% |
| 19 | Climate-related hazards | Description | How does acute disruption of water and sanitation services due to extreme weather events impact personal hygiene behaviours? | 123 | 75.9% | 70.5% | 75.8% | 85.0% | 82.4% | 75.0% |
| 20 | Temperature-related climate drivers | Delivery | Given the food contamination risks resulting from higher temperatures and extreme weather events, what messages should be included in the promotion of safe food hygiene practices? | 126 | 74.8% | 70.3% | 74.9% | 84.8% | 65.5% | 67.9% |
| 21 | Water-related climate drivers | Delivery | Given climate-induced water scarcity and drought conditions, what options are there to conserve or recycle water at the household level for hygiene purposes? | 122 | 74.5% | 70.2% | 74.5% | 84.7% | 76.1% | 72.3% |
| 22 | Climate-related hazards | Delivery | What conditions for vectors and pests (e.g., mosquitoes, mice, cockroaches, and rats) during floods can be mitigated with effective domestic hygiene? | 117 | 74.2% | 70.1% | 74.4% | 84.6% | 60.2% | 65.6% |
| 23 | Temperature-related climate drivers | Description | How do changes in ambient temperature, humidity and precipitation affect food safety and the incidence of foodborne illness? | 126 | 73.7% | 70.0% | 73.3% | 84.4% | 50.5% | 61.5% |
| 24 | Climate-related hazards | Delivery | How well are interventions addressing gender-specific hygiene needs during extreme weather events? | 123 | 72.6% | 69.8% | 72.4% | 84.1% | 70.9% | 70.1% |
| 25 | Climate-related hazards | Delivery | How can effective behaviour change programmes for hygiene-related risks of climate change (e.g., floods, cyclones, climate-change induced water scarcity and drought) best be designed and delivered? | 122 | 69.7% | 69.2% | 69.6% | 83.4% | 74.1% | 71.5% |
| 26 | Water-related climate drivers | Description | How does the effect of climate change on women's and girls' hygiene affect their participation in wider society? | 120 | 69.2% | 69.1% | 69.0% | 83.3% | 64.7% | 67.5% |
| 27 | Climate-related hazards | Description | How can the hygiene needs of people with incontinence be addressed during extreme weather events? | 128 | 68.0% | 68.9% | 68.0% | 83.0% | 61.6% | 66.2% |
| 28 | Climate-related hazards | Delivery | What are appropriate channels of communication to populations about hygiene-related health risks before, during, and post extreme weather events? | 118 | 67.8% | 68.8% | 67.8% | 83.0% | 67.4% | 68.6% |
| 29 | Climate-related hazards | Development | What culturally acceptable products are needed in disaster preparedness kits for personal hygiene? | 117 | 67.3% | 68.7% | 67.3% | 82.9% | 73.3% | 71.2% |
| 30 | Climate-related hazards | Development | How can the coordination and implementation of hygiene-related interventions during and post extreme weather events be improved? | 128 | 66.7% | 68.6% | 66.7% | 82.7% | 65.3% | 67.8% |
| 31 | Climate-related hazards | Description | How resilient are hygiene services (i.e., facilities, hardware, infrastructure) to the effects of climate change? | 123 | 65.5% | 68.4% | 65.2% | 82.4% | 41.4% | 57.6% |
| 32 | Temperature-related climate drivers | Description | How will higher ambient temperatures, humidity and precipitation change the contamination of food across the whole food chain, from preparation, processing, storage, and consumption? | 121 | 65.3% | 68.3% | 65.3% | 82.4% | 66.5% | 68.3% |
| 33 | Climate-related hazards | Description | How can individuals, and those who support them, maintain menstrual health during extreme weather events? | 127.75 | 65.3% | 68.3% | 65.1% | 82.3% | 66.1% | 68.1% |
| 34 | Climate-related hazards | Description | How do individuals prioritise hygiene in relation to other needs during extreme weather events? | 123 | 64.1% | 68.1% | 63.7% | 82.0% | 69.4% | 69.5% |
| 35 | Water-related climate drivers | Description | How will climate change affect the current Burden of Disease estimates related to hygiene? | 126 | 63.8% | 68.0% | 63.3% | 81.9% | 54.3% | 63.1% |
| 36 | Water-related climate drivers | Description | How do domestic hygiene practices during extreme weather events (droughts, heavy precipitation, floods, and cyclones) change the risk of mosquito-borne diseases? | 123 | 62.6% | 67.8% | 62.6% | 81.7% | 55.5% | 63.62% |
| 37 | Climate-related hazards | Delivery | Which approaches are effective in preparing and restoring the hygiene supply chain to the population during extreme weather events? | 115 | 62.4% | 67.7% | 62.3% | 81.6% | 58.3% | 64.78% |
| 38 | Climate-related hazards | Description | What are the health risks of using floodwater for personal and domestic hygiene? | 117 | 61.4% | 67.5% | 61.4% | 81.4% | 70.3% | 69.87% |
| 39 | Climate-related hazards | Description | Do people experiencing homelessness have specific hygiene-related challenges during extreme weather events? | 127 | 61.4% | 67.5% | 61.3% | 81.4% | 59.6% | 65.35% |
| 40 | Climate-related hazards | Description | What are the risks of vaginal or reproductive tract infections (RTIs), urinary tract infections (UTIs), fungal infections, bacterial vaginosis, rashes, or discomfort due to inadequate menstrual hygiene from climate change events? And does this differ concerning climate hazards, such as saltwater intrusion, floods or cyclone waters, or drought, and if so, why? | 119 | 60.1% | 67.3% | 60.1% | 81.1% | 26.4% | 51.26% |
| 41 | Water-related climate drivers | Description | What is the gender-specific hygiene needs affected by climate change? | 120 | 57.3% | 66.7% | 57.3% | 80.4% | 59.3% | 65.21% |
| 42 | Climate-related hazards | Description | How does the reduced opportunity to practice personal hygiene (washing, bathing, showering) affect mental health during extreme weather events or periods of climate-induced water scarcity and drought? | 120 | 56.5% | 66.5% | 56.5% | 80.2% | 49.9% | 61.25% |
| 43 | Water-related climate drivers | Description | How does climate change affect the food preparation and hygiene practices of individuals, and how does this differ by climate events, such as heatwaves, high humidity, extreme weather events and/or climate-induced water scarcity? | 125.75 | 50.7% | 65.3% | 50.5% | 78.7% | 43.8% | 58.63% |
| 44 | Climate-related hazards | Delivery | What microbiological and chemical contaminants from floods can be mitigated with effective domestic hygiene? | 115 | 49.3% | 65.1% | 49.3% | 78.4% | 33.1% | 54.13% |
| 45 | Water-related climate drivers | Description | What will be the effect on skin conditions from inadequate personal hygiene due to climate change? | 119 | 46.4% | 64.5% | 46.1% | 77.6% | 41.6% | 57.71% |
| 46 | Water-related climate drivers | Description | How does climate-induced water scarcity affect an individual's experience of shame, embarrassment, and humiliation due to their lack of opportunity for personal or menstrual hygiene? | 120 | 46.4% | 64.5% | 46.5% | 77.7% | 24.9% | 50.63% |
| 47 | Water-related climate drivers | Description | Do climate-induced water scarcity and droughts change individual preferences for disposable hygiene materials (e.g. nappies, menstrual pads, incontinence pads)? | 122 | 46.2% | 64.4% | 46.0% | 77.6% | 50.5% | 61.48% |
| 48 | Water-related climate drivers | Description | How will climate change alter the risk of exposure to domestic animals and their waste, and how can this be reduced by hygiene in the domestic environment? | 120 | 42.2% | 63.6% | 41.8% | 76.6% | 34.1% | 54.55% |
| 49 | Climate-related hazards | Delivery | How are budgets of national and local stakeholders being allocated for climate-resilient hygiene efforts and services? | 121 | 41.7% | 63.5% | 41.8% | 76.6% | 30.3% | 52.92% |
| 50 | Climate-related hazards | Development | How are private water service providers considering and planning for the potential increase in hygiene needs during extreme weather events? | 128 | 40.2% | 63.2% | 40.1% | 76.2% | 39.1% | 56.64% |
| 51 | Climate-related hazards | Delivery | What hygiene products are needed for post-flood cleaning of households and domestic spaces? | 117 | 39.6% | 63.1% | 39.8% | 76.1% | 47.6% | 60.26% |
| 52 | Climate-related hazards | Delivery | What surfaces are most critical for cleaning during heavy rains and floods to maintain hygiene in the domestic environment? | 115 | 39.5% | 63.1% | 39.5% | 76.0% | 43.4% | 58.48% |
| 53 | Temperature-related climate drivers | Delivery | What is the association between higher ambient temperatures, precipitation variability and the effectiveness of handwashing interventions against diarrhoeal disease? | 120 | 36.6% | 62.5% | 36.3% | 75.2% | 39.6% | 56.88% |
| 54 | Climate-related hazards | Description | Do climate-induced cold spells or freezes affect an individual's capability, opportunity, or motivation to practice effective hygiene behaviours? | 126 | 32.6% | 61.6% | 32.9% | 74.4% | 28.5% | 52.18% |
| 55 | Temperature-related climate drivers | Discovery | What is the stability and durability of hygiene products (e.g., soap, detergents, cleaning products) under higher ambient temperature and humidity throughout the process of manufacturing, transportation, and use? | 120 | 16.4% | 58.3% | 16.3% | 70.3% | 13.6% | 45.83% |
| 56 | Temperature-related climate drivers | Description | What is the association between higher ambient temperatures, precipitation variability and hand-washing behaviours? | 121 | 1.7% | 55.3% | 1.6% | 66.7% | 9.7% | 44.21% |
| 57 | Temperature-related climate drivers | Description | What is the association between higher ambient temperatures, precipitation variability and hand contamination? | 121 | 0.0% | 55.0% | 0.0% | 66.3% | 0.0% | 40.08% |
