## Supplementary Material 4 for "Climate change, hygiene, and health: a research roadmap for climate adaptation"

### Table S1 – Research priorities stratified by organisation type

| 1. International NGO | | | | |
| --- | --- | --- | --- | --- |
| **Climate Hazards** | **RQ ID #** | **Research Question** | **# of Respondents** | **Scaled Weighted RPS (%)** |
| Climate-related hazards | 21 | What hygiene promotion actions and preparedness strategies are needed before or during extreme weather events to prepare populations for increased risks to health? | 45 | 100.0% |
| Water-related climate drivers | 33 | How do climate-induced water scarcity, drought or precipitation variability affect the infectious disease burden, and how is this mediated by changes to hygiene practices? | 43 | 86.6% |
| Climate-related hazards | 2 | During extreme weather events, how does solid waste contaminate the domestic environment and increase the exposure to infectious diseases and/or chemical pollutants? | 47 | 81.6% |
| Temperature-related climate drivers | 47 | How do extreme heat events affect an individual's use and consumption of water for hygiene practices? | 48 | 80.6% |
| Climate-related hazards | 16 | What are the determinants of hygiene behaviours during extreme weather events? | 45 | 79.5% |
| Water-related climate drivers | 32 | How do climate-induced water scarcity and drought affect individual's use and consumption of water for hygiene practices? | 43 | 79.2% |
| Climate-related hazards | 23 | What support is needed for people with disabilities and their caregivers during extreme weather events to maintain personal hygiene? | 44 | 79.2% |
| Temperature-related climate drivers | 55 | What is the association between higher ambient temperatures, precipitation variability and humidity with the incidence of diarrhoeal diseases including cholera? | 44 | 78.8% |
| Climate-related hazards | 9 | How do extreme weather events affect the capability of people with disabilities to maintain personal hygiene? | 46 | 77.8% |
| Temperature-related climate drivers | 52 | What effect will an increase in ambient temperature and humidity associated with climate change have on pathogen levels, with implications for human health, in water, food, soil, surfaces, and the environment? | 44 | 74.8% |
| Temperature-related climate drivers | 50 | How will extreme heat affect water collecting practices and the water available for hygiene practices? | 48 | 74.0% |
| Water-related climate drivers | 40 | What challenges are faced by women and girls concerning menstrual health and hygiene during extreme weather events, periods of climate-induced water scarcity, drought and saltwater intrusion? | 43 | 73.5% |
| Water-related climate drivers | 42 | What is the association between extreme weather events, climate-induced water scarcity, drought or saltwater intrusion with the incidence of neglected tropical diseases (NTDs), and how are these risks mediated by hygiene practices? | 43 | 72.3% |
| Climate-related hazards | 15 | What are appropriate channels of communication to populations about hygiene-related health risks before, during and post extreme weather events? | 45 | 71.5% |
| Water-related climate drivers | 26 | Are different approaches needed for hygiene promotion and hygiene behaviour change for populations living in water-scarce or drought conditions? | 43 | 71.2% |
| Water-related climate drivers | 28 | Does climate-induced water scarcity or drought affect an individual's capability, opportunity or motivation to practice effective hygiene behaviours? | 43 | 69.9% |
| Climate-related hazards | 10 | How do individuals prioritise hygiene in relation to other needs during extreme weather events? | 46 | 68.8% |
| Climate-related hazards | 7 | How can the hygiene needs of people with incontinence be addressed during extreme weather events? | 47 | 68.3% |
| Water-related climate drivers | 41 | What challenges are there in maintaining adequate personal hygiene during extreme weather events, periods of climate-induced water scarcity, drought and saltwater intrusion? | 43 | 67.3% |
| Climate-related hazards | 18 | What conditions for vectors and pests (e.g., mosquitoes, mice, cockroaches, and rats) during floods can be mitigated with effective domestic hygiene? | 45 | 67.1% |
| Climate-related hazards | 8 | How do domestic hygiene practices during extreme weather events (droughts, heavy precipitation, floods and cyclones) change the risk of mosquito-borne diseases? | 46 | 65.9% |
| Climate-related hazards | 13 | How resilient are hygiene services (i.e., facilities, hardware, infrastructure) to the effects of climate change? | 46 | 64.9% |
| Climate-related hazards | 5 | How can coordination and implementation of hygiene-related interventions during and post extreme weather events be improved? | 47 | 64.8% |
| Water-related climate drivers | 35 | How does the effect of climate change on women's and girls' hygiene affect their participation in wider society? | 43 | 63.7% |
| Climate-related hazards | 12 | How does chronic disruption of water and sanitation services due to extreme weather events impact personal hygiene behaviours? | 46 | 62.8% |
| Climate-related hazards | 14 | How well are interventions addressing gender-specific hygiene needs during extreme weather events? | 46 | 62.4% |
| Climate-related hazards | 3 | Given the microbial and chemical contamination risks of floodwater, what messages should be included in health promotion and hygiene behaviour change campaigns? | 47 | 62.2% |
| Temperature-related climate drivers | 49 | How will climate change affect the current Burden of Disease estimates related to hygiene? | 48 | 58.5% |
| Temperature-related climate drivers | 48 | How does climate change affect the food preparation and hygiene practices of individuals, and how does this differ by climate events, such as heatwaves, high humidity, extreme weather events and/or climate-induced water scarcity? | 47 | 58.2% |
| Water-related climate drivers | 39 | What are the risks of vaginal or reproductive tract infections (RTIs), urinary tract infections (UTIs), fungal infections, bacterial vaginosis, rashes or discomfort due to inadequate menstrual hygiene from climate change events? And does this differ concerning particular climate hazards, such as saltwater intrusion, flood or cyclone waters, or drought, and if so, why? | 43 | 57.9% |
| Water-related climate drivers | 29 | Given climate-induced water scarcity and drought conditions, what options are there to conserve or recycle water at the household level for hygiene purposes? | 43 | 56.4% |
| Temperature-related climate drivers | 45 | Given the food contamination risks resulting from higher temperatures and extreme weather events, what messages should be included in the promotion of safe food hygiene practices? | 48 | 55.4% |
| Climate-related hazards | 11 | How does acute disruption of water and sanitation services due to extreme weather events impact personal hygiene behaviours? | 46 | 55.2% |
| Temperature-related climate drivers | 46 | How do changes in ambient temperature, humidity and precipitation affect food safety and incidence of foodborne illness? | 48 | 54.7% |
| Water-related climate drivers | 27 | Do climate-induced water scarcity and droughts change individual preferences for disposable hygiene materials (e.g. nappies, menstrual pads, incontinence pads)? | 43 | 50.0% |
| Climate-related hazards | 6 | How can individuals, and those that support them, maintain menstrual health during extreme weather events? | 47 | 49.3% |
| Climate-related hazards | 1 | Do people experiencing homelessness have specific hygiene-related challenges during extreme weather events? | 47 | 49.2% |
| Climate-related hazards | 19 | What culturally acceptable products are needed in disaster preparedness kits for personal hygiene? | 45 | 48.6% |
| Water-related climate drivers | 31 | How can effective behaviour change programmes for hygiene-related risks of climate change (e.g., floods, cyclones, climate-change induced water scarcity and drought) best be designed and delivered? | 43 | 47.8% |
| Climate-related hazards | 17 | What are the health risks of using floodwater for personal and domestic hygiene? | 45 | 47.3% |
| Water-related climate drivers | 38 | What are the gender-specific hygiene needs affected by climate change? | 43 | 45.5% |
| Water-related climate drivers | 30 | How are budgets of national and local stakeholders being allocated for climate-resilient hygiene efforts and services? | 43 | 45.0% |
| Temperature-related climate drivers | 51 | How will higher ambient temperatures, humidity and precipitation change the contamination of food across the whole food chain, from preparation, processing, storage and consumption? | 44 | 44.0% |
| Climate-related hazards | 22 | What microbiological and chemical contaminants from floods can be mitigated with effective domestic hygiene? | 44 | 43.8% |
| Water-related climate drivers | 36 | How does the reduced opportunity to practice personal hygiene (washing, bathing, showering) affect mental health during extreme weather events or periods of climate-induced water scarcity and drought? | 43 | 43.1% |
| Climate-related hazards | 4 | How are private water service providers considering and planning for the potential increase in hygiene needs during extreme weather events? | 47 | 40.4% |
| Climate-related hazards | 25 | Which approaches are effective in preparing and restoring the hygiene supply chain to the population during extreme weather events? | 44 | 38.8% |
| Water-related climate drivers | 34 | How does climate-induced water scarcity affect individual's experience of shame, embarrassment and humiliation due to their lack of opportunity for personal or menstrual hygiene? | 43 | 36.8% |
| Climate-related hazards | 24 | What surfaces are most critical for cleaning during heavy rains and floods to maintain hygiene in the domestic environment? | 44 | 35.9% |
| Temperature-related climate drivers | 44 | Do climate-induced cold spells or freezes affect an individual's capability, opportunity or motivation to practice effective hygiene behaviours? | 48 | 35.6% |
| Water-related climate drivers | 43 | What will be the effect on skin conditions from inadequate personal hygiene due to climate change? | 43 | 34.1% |
| Water-related climate drivers | 37 | How will climate change alter the risk of exposure to domestic animals and their waste, and how can this be reduced by hygiene in the domestic environment? | 43 | 31.8% |
| Climate-related hazards | 20 | What hygiene products are needed for post-flood cleaning of households and domestic spaces? | 45 | 27.7% |
| Temperature-related climate drivers | 56 | What is the association between higher ambient temperatures, precipitation variability and the effectiveness of handwashing interventions against diarrhoeal disease? | 44 | 13.5% |
| Temperature-related climate drivers | 57 | What is the stability and durability of hygiene products, (e.g., soap, detergents, cleaning products) under higher ambient temperature and humidity throughout the process of manufacturing, transportation and use? | 44 | 3.1% |
| Temperature-related climate drivers | 54 | What is the association between higher ambient temperatures, precipitation variability and hand-washing behaviours? | 44 | 0.9% |
| Temperature-related climate drivers | 53 | What is the association between higher ambient temperatures, precipitation variability and hand contamination? | 44 | 0.0% |

| 2. Academic | | | | |
| --- | --- | --- | --- | --- |
| **Climate Hazards** | **RQ ID #** | **Research Question** | **# of Respondents** | **Scaled Weighted RPS (%)** |
| Climate-related hazards | 19 | What culturally acceptable products are needed in disaster preparedness kits for personal hygiene? | 29 | 100.0% |
| Climate-related hazards | 6 | How can individuals, and those that support them, maintain menstrual health during extreme weather events? | 32 | 99.1% |
| Climate-related hazards | 2 | During extreme weather events, how does solid waste contaminate the domestic environment and increase the exposure to infectious diseases and/or chemical pollutants? | 32 | 97.3% |
| Climate-related hazards | 9 | How do extreme weather events affect the capability of people with disabilities to maintain personal hygiene? | 30 | 97.0% |
| Water-related climate drivers | 32 | How do climate-induced water scarcity and drought affect individual's use and consumption of water for hygiene practices? | 31 | 96.7% |
| Water-related climate drivers | 28 | Does climate-induced water scarcity or drought affect an individual's capability, opportunity or motivation to practice effective hygiene behaviours? | 31 | 96.0% |
| Water-related climate drivers | 40 | What challenges are faced by women and girls concerning menstrual health and hygiene during extreme weather events, periods of climate-induced water scarcity, drought and saltwater intrusion? | 30 | 95.9% |
| Temperature-related climate drivers | 46 | How do changes in ambient temperature, humidity and precipitation affect food safety and incidence of foodborne illness? | 30 | 95.9% |
| Water-related climate drivers | 29 | Given climate-induced water scarcity and drought conditions, what options are there to conserve or recycle water at the household level for hygiene purposes? | 31 | 95.7% |
| Water-related climate drivers | 26 | Are different approaches needed for hygiene promotion and hygiene behaviour change for populations living in water-scarce or drought conditions? | 31 | 94.4% |
| Climate-related hazards | 23 | What support is needed for people with disabilities and their caregivers during extreme weather events to maintain personal hygiene? | 29 | 93.6% |
| Water-related climate drivers | 33 | How do climate-induced water scarcity, drought or precipitation variability affect the infectious disease burden, and how is this mediated by changes to hygiene practices? | 31 | 93.2% |
| Temperature-related climate drivers | 47 | How do extreme heat events affect an individual's use and consumption of water for hygiene practices? | 30 | 93.0% |
| Climate-related hazards | 21 | What hygiene promotion actions and preparedness strategies are needed before or during extreme weather events to prepare populations for increased risks to health? | 29 | 91.6% |
| Climate-related hazards | 12 | How does chronic disruption of water and sanitation services due to extreme weather events impact personal hygiene behaviours? | 30 | 91.5% |
| Climate-related hazards | 3 | Given the microbial and chemical contamination risks of floodwater, what messages should be included in health promotion and hygiene behaviour change campaigns? | 32 | 89.6% |
| Climate-related hazards | 25 | Which approaches are effective in preparing and restoring the hygiene supply chain to the population during extreme weather events? | 29 | 88.6% |
| Temperature-related climate drivers | 55 | What is the association between higher ambient temperatures, precipitation variability and humidity with the incidence of diarrhoeal diseases including cholera? | 30 | 88.2% |
| Water-related climate drivers | 37 | How will climate change alter the risk of exposure to domestic animals and their waste, and how can this be reduced by hygiene in the domestic environment? | 31 | 88.1% |
| Climate-related hazards | 14 | How well are interventions addressing gender-specific hygiene needs during extreme weather events? | 30 | 87.7% |
| Climate-related hazards | 5 | How can coordination and implementation of hygiene-related interventions during and post extreme weather events be improved? | 32 | 87.4% |
| Temperature-related climate drivers | 45 | Given the food contamination risks resulting from higher temperatures and extreme weather events, what messages should be included in the promotion of safe food hygiene practices? | 30 | 87.2% |
| Water-related climate drivers | 42 | What is the association between extreme weather events, climate-induced water scarcity, drought or saltwater intrusion with the incidence of neglected tropical diseases (NTDs), and how are these risks mediated by hygiene practices? | 30 | 86.6% |
| Water-related climate drivers | 35 | How does the effect of climate change on women's and girls' hygiene affect their participation in wider society? | 31 | 86.5% |
| Water-related climate drivers | 36 | How does the reduced opportunity to practice personal hygiene (washing, bathing, showering) affect mental health during extreme weather events or periods of climate-induced water scarcity and drought? | 31 | 86.0% |
| Water-related climate drivers | 41 | What challenges are there in maintaining adequate personal hygiene during extreme weather events, periods of climate-induced water scarcity, drought and saltwater intrusion? | 30 | 85.9% |
| Temperature-related climate drivers | 52 | What effect will an increase in ambient temperature and humidity associated with climate change have on pathogen levels, with implications for human health, in water, food, soil, surfaces, and the environment? | 30 | 85.3% |
| Climate-related hazards | 11 | How does acute disruption of water and sanitation services due to extreme weather events impact personal hygiene behaviours? | 30 | 84.2% |
| Climate-related hazards | 1 | Do people experiencing homelessness have specific hygiene-related challenges during extreme weather events? | 32 | 83.8% |
| Temperature-related climate drivers | 51 | How will higher ambient temperatures, humidity and precipitation change the contamination of food across the whole food chain, from preparation, processing, storage and consumption? | 30 | 83.4% |
| Climate-related hazards | 17 | What are the health risks of using floodwater for personal and domestic hygiene? | 29 | 82.5% |
| Climate-related hazards | 13 | How resilient are hygiene services (i.e., facilities, hardware, infrastructure) to the effects of climate change? | 30 | 81.2% |
| Climate-related hazards | 20 | What hygiene products are needed for post-flood cleaning of households and domestic spaces? | 29 | 79.1% |
| Temperature-related climate drivers | 50 | How will extreme heat affect water collecting practices and the water available for hygiene practices? | 30 | 78.5% |
| Climate-related hazards | 22 | What microbiological and chemical contaminants from floods can be mitigated with effective domestic hygiene? | 29 | 77.9% |
| Climate-related hazards | 18 | What conditions for vectors and pests (e.g., mosquitoes, mice, cockroaches, and rats) during floods can be mitigated with effective domestic hygiene? | 29 | 74.8% |
| Water-related climate drivers | 31 | How can effective behaviour change programmes for hygiene-related risks of climate change (e.g., floods, cyclones, climate-change induced water scarcity and drought) best be designed and delivered? | 31 | 73.1% |
| Climate-related hazards | 15 | What are appropriate channels of communication to populations about hygiene-related health risks before, during and post extreme weather events? | 29 | 72.7% |
| Climate-related hazards | 16 | What are the determinants of hygiene behaviours during extreme weather events? | 28 | 72.7% |
| Water-related climate drivers | 38 | What are the gender-specific hygiene needs affected by climate change? | 31 | 71.0% |
| Climate-related hazards | 7 | How can the hygiene needs of people with incontinence be addressed during extreme weather events? | 32 | 67.9% |
| Water-related climate drivers | 30 | How are budgets of national and local stakeholders being allocated for climate-resilient hygiene efforts and services? | 31 | 66.9% |
| Water-related climate drivers | 39 | What are the risks of vaginal or reproductive tract infections (RTIs), urinary tract infections (UTIs), fungal infections, bacterial vaginosis, rashes or discomfort due to inadequate menstrual hygiene from climate change events? And does this differ concerning particular climate hazards, such as saltwater intrusion, flood or cyclone waters, or drought, and if so, why? | 30 | 64.3% |
| Water-related climate drivers | 43 | What will be the effect on skin conditions from inadequate personal hygiene due to climate change? | 30 | 63.2% |
| Climate-related hazards | 4 | How are private water service providers considering and planning for the potential increase in hygiene needs during extreme weather events? | 32 | 63.1% |
| Climate-related hazards | 10 | How do individuals prioritise hygiene in relation to other needs during extreme weather events? | 30 | 62.9% |
| Water-related climate drivers | 34 | How does climate-induced water scarcity affect individual's experience of shame, embarrassment and humiliation due to their lack of opportunity for personal or menstrual hygiene? | 31 | 62.8% |
| Temperature-related climate drivers | 49 | How will climate change affect the current Burden of Disease estimates related to hygiene? | 30 | 60.0% |
| Temperature-related climate drivers | 56 | What is the association between higher ambient temperatures, precipitation variability and the effectiveness of handwashing interventions against diarrhoeal disease? | 30 | 50.8% |
| Water-related climate drivers | 27 | Do climate-induced water scarcity and droughts change individual preferences for disposable hygiene materials (e.g. nappies, menstrual pads, incontinence pads)? | 31 | 49.7% |
| Temperature-related climate drivers | 57 | What is the stability and durability of hygiene products, (e.g., soap, detergents, cleaning products) under higher ambient temperature and humidity throughout the process of manufacturing, transportation and use? | 30 | 49.6% |
| Climate-related hazards | 8 | How do domestic hygiene practices during extreme weather events (droughts, heavy precipitation, floods and cyclones) change the risk of mosquito-borne diseases? | 30 | 47.5% |
| Temperature-related climate drivers | 48 | How does climate change affect the food preparation and hygiene practices of individuals, and how does this differ by climate events, such as heatwaves, high humidity, extreme weather events and/or climate-induced water scarcity? | 30 | 46.8% |
| Climate-related hazards | 24 | What surfaces are most critical for cleaning during heavy rains and floods to maintain hygiene in the domestic environment? | 29 | 37.2% |
| Temperature-related climate drivers | 44 | Do climate-induced cold spells or freezes affect an individual's capability, opportunity or motivation to practice effective hygiene behaviours? | 30 | 26.3% |
| Temperature-related climate drivers | 54 | What is the association between higher ambient temperatures, precipitation variability and hand-washing behaviours? | 30 | 4.1% |
| Temperature-related climate drivers | 53 | What is the association between higher ambient temperatures, precipitation variability and hand contamination? | 30 | 0.0% |

| 3. Multilateral Agency | | | | |
| --- | --- | --- | --- | --- |
| **Climate Hazards** | **RQ ID #** | **Research Question** | **# of Respondents** | **Scaled Weighted RPS (%)** |
| Temperature-related climate drivers | 52 | What effect will an increase in ambient temperature and humidity associated with climate change have on pathogen levels, with implications for human health, in water, food, soil, surfaces, and the environment? | 21 | 100.0% |
| Climate-related hazards | 3 | Given the microbial and chemical contamination risks of floodwater, what messages should be included in health promotion and hygiene behaviour change campaigns? | 21 | 99.8% |
| Climate-related hazards | 21 | What hygiene promotion actions and preparedness strategies are needed before or during extreme weather events to prepare populations for increased risks to health? | 18 | 95.1% |
| Water-related climate drivers | 33 | How do climate-induced water scarcity, drought or precipitation variability affect the infectious disease burden, and how is this mediated by changes to hygiene practices? | 20 | 93.0% |
| Temperature-related climate drivers | 55 | What is the association between higher ambient temperatures, precipitation variability and humidity with the incidence of diarrhoeal diseases including cholera? | 21 | 92.8% |
| Climate-related hazards | 16 | What are the determinants of hygiene behaviours during extreme weather events? | 19 | 86.6% |
| Water-related climate drivers | 31 | How can effective behaviour change programmes for hygiene-related risks of climate change (e.g., floods, cyclones, climate-change induced water scarcity and drought) best be designed and delivered? | 20 | 84.7% |
| Climate-related hazards | 13 | How resilient are hygiene services (i.e., facilities, hardware, infrastructure) to the effects of climate change? | 21 | 83.5% |
| Temperature-related climate drivers | 45 | Given the food contamination risks resulting from higher temperatures and extreme weather events, what messages should be included in the promotion of safe food hygiene practices? | 21 | 80.8% |
| Water-related climate drivers | 32 | How do climate-induced water scarcity and drought affect individual's use and consumption of water for hygiene practices? | 20 | 79.5% |
| Climate-related hazards | 11 | How does acute disruption of water and sanitation services due to extreme weather events impact personal hygiene behaviours? | 21 | 78.9% |
| Climate-related hazards | 25 | Which approaches are effective in preparing and restoring the hygiene supply chain to the population during extreme weather events? | 18 | 78.6% |
| Climate-related hazards | 10 | How do individuals prioritise hygiene in relation to other needs during extreme weather events? | 21 | 74.2% |
| Climate-related hazards | 18 | What conditions for vectors and pests (e.g., mosquitoes, mice, cockroaches, and rats) during floods can be mitigated with effective domestic hygiene? | 18 | 72.6% |
| Water-related climate drivers | 34 | How does climate-induced water scarcity affect individual's experience of shame, embarrassment and humiliation due to their lack of opportunity for personal or menstrual hygiene? | 20 | 71.1% |
| Water-related climate drivers | 26 | Are different approaches needed for hygiene promotion and hygiene behaviour change for populations living in water-scarce or drought conditions? | 20 | 69.4% |
| Climate-related hazards | 2 | During extreme weather events, how does solid waste contaminate the domestic environment and increase the exposure to infectious diseases and/or chemical pollutants? | 21 | 67.8% |
| Climate-related hazards | 12 | How does chronic disruption of water and sanitation services due to extreme weather events impact personal hygiene behaviours? | 21 | 67.4% |
| Climate-related hazards | 6 | How can individuals, and those that support them, maintain menstrual health during extreme weather events? | 21 | 66.2% |
| Climate-related hazards | 23 | What support is needed for people with disabilities and their caregivers during extreme weather events to maintain personal hygiene? | 18 | 63.0% |
| Climate-related hazards | 8 | How do domestic hygiene practices during extreme weather events (droughts, heavy precipitation, floods and cyclones) change the risk of mosquito-borne diseases? | 21 | 62.7% |
| Water-related climate drivers | 29 | Given climate-induced water scarcity and drought conditions, what options are there to conserve or recycle water at the household level for hygiene purposes? | 20 | 62.6% |
| Climate-related hazards | 7 | How can the hygiene needs of people with incontinence be addressed during extreme weather events? | 21 | 62.6% |
| Temperature-related climate drivers | 50 | How will extreme heat affect water collecting practices and the water available for hygiene practices? | 21 | 62.2% |
| Water-related climate drivers | 41 | What challenges are there in maintaining adequate personal hygiene during extreme weather events, periods of climate-induced water scarcity, drought and saltwater intrusion? | 20 | 60.5% |
| Temperature-related climate drivers | 56 | What is the association between higher ambient temperatures, precipitation variability and the effectiveness of handwashing interventions against diarrhoeal disease? | 20 | 59.5% |
| Temperature-related climate drivers | 51 | How will higher ambient temperatures, humidity and precipitation change the contamination of food across the whole food chain, from preparation, processing, storage and consumption? | 21 | 56.0% |
| Climate-related hazards | 5 | How can coordination and implementation of hygiene-related interventions during and post extreme weather events be improved? | 21 | 55.7% |
| Temperature-related climate drivers | 47 | How do extreme heat events affect an individual's use and consumption of water for hygiene practices? | 21 | 54.5% |
| Temperature-related climate drivers | 46 | How do changes in ambient temperature, humidity and precipitation affect food safety and incidence of foodborne illness? | 21 | 53.4% |
| Temperature-related climate drivers | 49 | How will climate change affect the current Burden of Disease estimates related to hygiene? | 21 | 51.8% |
| Climate-related hazards | 9 | How do extreme weather events affect the capability of people with disabilities to maintain personal hygiene? | 21 | 47.4% |
| Climate-related hazards | 15 | What are appropriate channels of communication to populations about hygiene-related health risks before, during and post extreme weather events? | 19 | 47.3% |
| Temperature-related climate drivers | 57 | What is the stability and durability of hygiene products, (e.g., soap, detergents, cleaning products) under higher ambient temperature and humidity throughout the process of manufacturing, transportation and use? | 20 | 44.6% |
| Climate-related hazards | 14 | How well are interventions addressing gender-specific hygiene needs during extreme weather events? | 21 | 43.3% |
| Climate-related hazards | 1 | Do people experiencing homelessness have specific hygiene-related challenges during extreme weather events? | 21 | 42.7% |
| Water-related climate drivers | 28 | Does climate-induced water scarcity or drought affect an individual's capability, opportunity or motivation to practice effective hygiene behaviours? | 20 | 41.6% |
| Climate-related hazards | 17 | What are the health risks of using floodwater for personal and domestic hygiene? | 18 | 41.1% |
| Water-related climate drivers | 40 | What challenges are faced by women and girls concerning menstrual health and hygiene during extreme weather events, periods of climate-induced water scarcity, drought and saltwater intrusion? | 20 | 40.7% |
| Climate-related hazards | 19 | What culturally acceptable products are needed in disaster preparedness kits for personal hygiene? | 18 | 39.6% |
| Water-related climate drivers | 42 | What is the association between extreme weather events, climate-induced water scarcity, drought or saltwater intrusion with the incidence of neglected tropical diseases (NTDs), and how are these risks mediated by hygiene practices? | 20 | 39.2% |
| Water-related climate drivers | 38 | What are the gender-specific hygiene needs affected by climate change? | 20 | 38.8% |
| Water-related climate drivers | 39 | What are the risks of vaginal or reproductive tract infections (RTIs), urinary tract infections (UTIs), fungal infections, bacterial vaginosis, rashes or discomfort due to inadequate menstrual hygiene from climate change events? And does this differ concerning particular climate hazards, such as saltwater intrusion, flood or cyclone waters, or drought, and if so, why? | 20 | 36.6% |
| Water-related climate drivers | 36 | How does the reduced opportunity to practice personal hygiene (washing, bathing, showering) affect mental health during extreme weather events or periods of climate-induced water scarcity and drought? | 20 | 34.6% |
| Climate-related hazards | 4 | How are private water service providers considering and planning for the potential increase in hygiene needs during extreme weather events? | 21 | 31.6% |
| Climate-related hazards | 24 | What surfaces are most critical for cleaning during heavy rains and floods to maintain hygiene in the domestic environment? | 18 | 31.2% |
| Water-related climate drivers | 35 | How does the effect of climate change on women's and girls' hygiene affect their participation in wider society? | 20 | 26.9% |
| Water-related climate drivers | 30 | How are budgets of national and local stakeholders being allocated for climate-resilient hygiene efforts and services? | 20 | 21.2% |
| Temperature-related climate drivers | 48 | How does climate change affect the food preparation and hygiene practices of individuals, and how does this differ by climate events, such as heatwaves, high humidity, extreme weather events and/or climate-induced water scarcity? | 21 | 20.6% |
| Climate-related hazards | 22 | What microbiological and chemical contaminants from floods can be mitigated with effective domestic hygiene? | 18 | 16.9% |
| Water-related climate drivers | 27 | Do climate-induced water scarcity and droughts change individual preferences for disposable hygiene materials (e.g. nappies, menstrual pads, incontinence pads)? | 20 | 15.1% |
| Temperature-related climate drivers | 54 | What is the association between higher ambient temperatures, precipitation variability and hand-washing behaviours? | 21 | 8.0% |
| Climate-related hazards | 20 | What hygiene products are needed for post-flood cleaning of households and domestic spaces? | 18 | 6.6% |
| Temperature-related climate drivers | 53 | What is the association between higher ambient temperatures, precipitation variability and hand contamination? | 21 | 6.0% |
| Water-related climate drivers | 43 | What will be the effect on skin conditions from inadequate personal hygiene due to climate change? | 20 | 0.7% |
| Temperature-related climate drivers | 44 | Do climate-induced cold spells or freezes affect an individual's capability, opportunity or motivation to practice effective hygiene behaviours? | 21 | 0.6% |
| Water-related climate drivers | 37 | How will climate change alter the risk of exposure to domestic animals and their waste, and how can this be reduced by hygiene in the domestic environment? | 20 | 0.0% |

| 4. National NGO | | | | |
| --- | --- | --- | --- | --- |
| **Climate Hazards** | **RQ ID #** | **Research Question** | **# of Respondents** | **Scaled Weighted RPS (%)** |
| Climate-related hazards | 19 | What culturally acceptable products are needed in disaster preparedness kits for personal hygiene? | 9 | 100.0% |
| Water-related climate drivers | 28 | Does climate-induced water scarcity or drought affect an individual's capability, opportunity or motivation to practice effective hygiene behaviours? | 11 | 99.2% |
| Water-related climate drivers | 38 | What are the gender-specific hygiene needs affected by climate change? | 10 | 98.1% |
| Water-related climate drivers | 43 | What will be the effect on skin conditions from inadequate personal hygiene due to climate change? | 10 | 96.2% |
| Temperature-related climate drivers | 46 | How do changes in ambient temperature, humidity and precipitation affect food safety and incidence of foodborne illness? | 10 | 96.2% |
| Water-related climate drivers | 40 | What challenges are faced by women and girls concerning menstrual health and hygiene during extreme weather events, periods of climate-induced water scarcity, drought and saltwater intrusion? | 10 | 94.9% |
| Temperature-related climate drivers | 56 | What is the association between higher ambient temperatures, precipitation variability and the effectiveness of handwashing interventions against diarrhoeal disease? | 10 | 93.8% |
| Temperature-related climate drivers | 52 | What effect will an increase in ambient temperature and humidity associated with climate change have on pathogen levels, with implications for human health, in water, food, soil, surfaces, and the environment? | 10 | 93.6% |
| Temperature-related climate drivers | 55 | What is the association between higher ambient temperatures, precipitation variability and humidity with the incidence of diarrhoeal diseases including cholera? | 10 | 93.2% |
| Temperature-related climate drivers | 51 | How will higher ambient temperatures, humidity and precipitation change the contamination of food across the whole food chain, from preparation, processing, storage and consumption? | 10 | 89.7% |
| Climate-related hazards | 20 | What hygiene products are needed for post-flood cleaning of households and domestic spaces? | 9 | 87.2% |
| Water-related climate drivers | 33 | How do climate-induced water scarcity, drought or precipitation variability affect the infectious disease burden, and how is this mediated by changes to hygiene practices? | 10 | 86.7% |
| Water-related climate drivers | 41 | What challenges are there in maintaining adequate personal hygiene during extreme weather events, periods of climate-induced water scarcity, drought and saltwater intrusion? | 10 | 85.2% |
| Water-related climate drivers | 35 | How does the effect of climate change on women's and girls' hygiene affect their participation in wider society? | 10 | 84.6% |
| Climate-related hazards | 23 | What support is needed for people with disabilities and their caregivers during extreme weather events to maintain personal hygiene? | 9 | 84.2% |
| Water-related climate drivers | 32 | How do climate-induced water scarcity and drought affect individual's use and consumption of water for hygiene practices? | 11 | 83.8% |
| Climate-related hazards | 25 | Which approaches are effective in preparing and restoring the hygiene supply chain to the population during extreme weather events? | 9 | 83.1% |
| Climate-related hazards | 17 | What are the health risks of using floodwater for personal and domestic hygiene? | 9 | 82.4% |
| Climate-related hazards | 21 | What hygiene promotion actions and preparedness strategies are needed before or during extreme weather events to prepare populations for increased risks to health? | 9 | 82.2% |
| Temperature-related climate drivers | 49 | How will climate change affect the current Burden of Disease estimates related to hygiene? | 10 | 77.1% |
| Temperature-related climate drivers | 45 | Given the food contamination risks resulting from higher temperatures and extreme weather events, what messages should be included in the promotion of safe food hygiene practices? | 10 | 76.7% |
| Water-related climate drivers | 31 | How can effective behaviour change programmes for hygiene-related risks of climate change (e.g., floods, cyclones, climate-change induced water scarcity and drought) best be designed and delivered? | 11 | 75.9% |
| Climate-related hazards | 9 | How do extreme weather events affect the capability of people with disabilities to maintain personal hygiene? | 8 | 74.0% |
| Climate-related hazards | 11 | How does acute disruption of water and sanitation services due to extreme weather events impact personal hygiene behaviours? | 8 | 74.0% |
| Climate-related hazards | 10 | How do individuals prioritise hygiene in relation to other needs during extreme weather events? | 8 | 73.8% |
| Climate-related hazards | 15 | What are appropriate channels of communication to populations about hygiene-related health risks before, during and post extreme weather events? | 9 | 72.7% |
| Temperature-related climate drivers | 48 | How does climate change affect the food preparation and hygiene practices of individuals, and how does this differ by climate events, such as heatwaves, high humidity, extreme weather events and/or climate-induced water scarcity? | 10 | 72.6% |
| Climate-related hazards | 2 | During extreme weather events, how does solid waste contaminate the domestic environment and increase the exposure to infectious diseases and/or chemical pollutants? | 10 | 72.2% |
| Climate-related hazards | 22 | What microbiological and chemical contaminants from floods can be mitigated with effective domestic hygiene? | 9 | 71.6% |
| Climate-related hazards | 8 | How do domestic hygiene practices during extreme weather events (droughts, heavy precipitation, floods and cyclones) change the risk of mosquito-borne diseases? | 8 | 71.3% |
| Water-related climate drivers | 37 | How will climate change alter the risk of exposure to domestic animals and their waste, and how can this be reduced by hygiene in the domestic environment? | 10 | 68.4% |
| Water-related climate drivers | 26 | Are different approaches needed for hygiene promotion and hygiene behaviour change for populations living in water-scarce or drought conditions? | 11 | 68.1% |
| Water-related climate drivers | 29 | Given climate-induced water scarcity and drought conditions, what options are there to conserve or recycle water at the household level for hygiene purposes? | 11 | 68.1% |
| Water-related climate drivers | 42 | What is the association between extreme weather events, climate-induced water scarcity, drought or saltwater intrusion with the incidence of neglected tropical diseases (NTDs), and how are these risks mediated by hygiene practices? | 10 | 67.7% |
| Temperature-related climate drivers | 53 | What is the association between higher ambient temperatures, precipitation variability and hand contamination? | 10 | 64.0% |
| Temperature-related climate drivers | 54 | What is the association between higher ambient temperatures, precipitation variability and hand-washing behaviours? | 10 | 64.0% |
| Climate-related hazards | 3 | Given the microbial and chemical contamination risks of floodwater, what messages should be included in health promotion and hygiene behaviour change campaigns? | 10 | 63.4% |
| Climate-related hazards | 24 | What surfaces are most critical for cleaning during heavy rains and floods to maintain hygiene in the domestic environment? | 9 | 62.9% |
| Climate-related hazards | 14 | How well are interventions addressing gender-specific hygiene needs during extreme weather events? | 8 | 62.1% |
| Water-related climate drivers | 27 | Do climate-induced water scarcity and droughts change individual preferences for disposable hygiene materials (e.g. nappies, menstrual pads, incontinence pads)? | 11 | 60.9% |
| Climate-related hazards | 1 | Do people experiencing homelessness have specific hygiene-related challenges during extreme weather events? | 10 | 59.8% |
| Temperature-related climate drivers | 44 | Do climate-induced cold spells or freezes affect an individual's capability, opportunity or motivation to practice effective hygiene behaviours? | 10 | 59.2% |
| Temperature-related climate drivers | 47 | How do extreme heat events affect an individual's use and consumption of water for hygiene practices? | 10 | 59.2% |
| Climate-related hazards | 16 | What are the determinants of hygiene behaviours during extreme weather events? | 9 | 58.9% |
| Climate-related hazards | 12 | How does chronic disruption of water and sanitation services due to extreme weather events impact personal hygiene behaviours? | 8 | 57.7% |
| Water-related climate drivers | 36 | How does the reduced opportunity to practice personal hygiene (washing, bathing, showering) affect mental health during extreme weather events or periods of climate-induced water scarcity and drought? | 10 | 54.6% |
| Water-related climate drivers | 39 | What are the risks of vaginal or reproductive tract infections (RTIs), urinary tract infections (UTIs), fungal infections, bacterial vaginosis, rashes or discomfort due to inadequate menstrual hygiene from climate change events? And does this differ concerning particular climate hazards, such as saltwater intrusion, flood or cyclone waters, or drought, and if so, why? | 10 | 52.4% |
| Climate-related hazards | 18 | What conditions for vectors and pests (e.g., mosquitoes, mice, cockroaches, and rats) during floods can be mitigated with effective domestic hygiene? | 9 | 47.3% |
| Temperature-related climate drivers | 50 | How will extreme heat affect water collecting practices and the water available for hygiene practices? | 10 | 47.1% |
| Climate-related hazards | 7 | How can the hygiene needs of people with incontinence be addressed during extreme weather events? | 10 | 42.9% |
| Climate-related hazards | 5 | How can coordination and implementation of hygiene-related interventions during and post extreme weather events be improved? | 10 | 38.2% |
| Climate-related hazards | 4 | How are private water service providers considering and planning for the potential increase in hygiene needs during extreme weather events? | 10 | 34.9% |
| Climate-related hazards | 13 | How resilient are hygiene services (i.e., facilities, hardware, infrastructure) to the effects of climate change? | 8 | 31.0% |
| Temperature-related climate drivers | 57 | What is the stability and durability of hygiene products, (e.g., soap, detergents, cleaning products) under higher ambient temperature and humidity throughout the process of manufacturing, transportation and use? | 10 | 26.8% |
| Water-related climate drivers | 30 | How are budgets of national and local stakeholders being allocated for climate-resilient hygiene efforts and services? | 10 | 21.5% |
| Water-related climate drivers | 34 | How does climate-induced water scarcity affect individual's experience of shame, embarrassment and humiliation due to their lack of opportunity for personal or menstrual hygiene? | 10 | 6.3% |
| Climate-related hazards | 6 | How can individuals, and those that support them, maintain menstrual health during extreme weather events? | 9 | 0.0% |

| 5. Other | | | | |
| --- | --- | --- | --- | --- |
| **Climate Hazards** | **RQ ID #** | **Research Question** | **# of Respondents** | **Scaled Weighted RPS (%)** |
| Climate-related hazards | 9 | How do extreme weather events affect the capability of people with disabilities to maintain personal hygiene? | 6 | 100.0% |
| Climate-related hazards | 23 | What support is needed for people with disabilities and their caregivers during extreme weather events to maintain personal hygiene? | 6 | 95.0% |
| Water-related climate drivers | 28 | Does climate-induced water scarcity or drought affect an individual's capability, opportunity or motivation to practice effective hygiene behaviours? | 7 | 87.0% |
| Climate-related hazards | 19 | What culturally acceptable products are needed in disaster preparedness kits for personal hygiene? | 6 | 84.4% |
| Climate-related hazards | 18 | What conditions for vectors and pests (e.g., mosquitoes, mice, cockroaches, and rats) during floods can be mitigated with effective domestic hygiene? | 6 | 83.7% |
| Water-related climate drivers | 32 | How do climate-induced water scarcity and drought affect individual's use and consumption of water for hygiene practices? | 7 | 82.0% |
| Climate-related hazards | 6 | How can individuals, and those that support them, maintain menstrual health during extreme weather events? | 6 | 81.2% |
| Water-related climate drivers | 42 | What is the association between extreme weather events, climate-induced water scarcity, drought or saltwater intrusion with the incidence of neglected tropical diseases (NTDs), and how are these risks mediated by hygiene practices? | 7 | 80.2% |
| Water-related climate drivers | 43 | What will be the effect on skin conditions from inadequate personal hygiene due to climate change? | 7 | 80.2% |
| Water-related climate drivers | 33 | How do climate-induced water scarcity, drought or precipitation variability affect the infectious disease burden, and how is this mediated by changes to hygiene practices? | 7 | 80.2% |
| Climate-related hazards | 8 | How do domestic hygiene practices during extreme weather events (droughts, heavy precipitation, floods and cyclones) change the risk of mosquito-borne diseases? | 6 | 78.6% |
| Climate-related hazards | 2 | During extreme weather events, how does solid waste contaminate the domestic environment and increase the exposure to infectious diseases and/or chemical pollutants? | 7 | 78.4% |
| Climate-related hazards | 12 | How does chronic disruption of water and sanitation services due to extreme weather events impact personal hygiene behaviours? | 6 | 78.4% |
| Temperature-related climate drivers | 46 | How do changes in ambient temperature, humidity and precipitation affect food safety and incidence of foodborne illness? | 7 | 78.2% |
| Water-related climate drivers | 31 | How can effective behaviour change programmes for hygiene-related risks of climate change (e.g., floods, cyclones, climate-change induced water scarcity and drought) best be designed and delivered? | 7 | 77.0% |
| Temperature-related climate drivers | 49 | How will climate change affect the current Burden of Disease estimates related to hygiene? | 7 | 74.0% |
| Climate-related hazards | 16 | What are the determinants of hygiene behaviours during extreme weather events? | 6 | 73.8% |
| Climate-related hazards | 21 | What hygiene promotion actions and preparedness strategies are needed before or during extreme weather events to prepare populations for increased risks to health? | 6 | 73.8% |
| Climate-related hazards | 14 | How well are interventions addressing gender-specific hygiene needs during extreme weather events? | 6 | 69.2% |
| Temperature-related climate drivers | 48 | How does climate change affect the food preparation and hygiene practices of individuals, and how does this differ by climate events, such as heatwaves, high humidity, extreme weather events and/or climate-induced water scarcity? | 7 | 69.2% |
| Water-related climate drivers | 41 | What challenges are there in maintaining adequate personal hygiene during extreme weather events, periods of climate-induced water scarcity, drought and saltwater intrusion? | 7 | 68.8% |
| Climate-related hazards | 1 | Do people experiencing homelessness have specific hygiene-related challenges during extreme weather events? | 5 | 68.6% |
| Climate-related hazards | 7 | How can the hygiene needs of people with incontinence be addressed during extreme weather events? | 6 | 68.3% |
| Climate-related hazards | 20 | What hygiene products are needed for post-flood cleaning of households and domestic spaces? | 6 | 68.3% |
| Climate-related hazards | 11 | How does acute disruption of water and sanitation services due to extreme weather events impact personal hygiene behaviours? | 6 | 68.1% |
| Temperature-related climate drivers | 44 | Do climate-induced cold spells or freezes affect an individual's capability, opportunity or motivation to practice effective hygiene behaviours? | 7 | 67.7% |
| Temperature-related climate drivers | 55 | What is the association between higher ambient temperatures, precipitation variability and humidity with the incidence of diarrhoeal diseases including cholera? | 6 | 67.3% |
| Water-related climate drivers | 39 | What are the risks of vaginal or reproductive tract infections (RTIs), urinary tract infections (UTIs), fungal infections, bacterial vaginosis, rashes or discomfort due to inadequate menstrual hygiene from climate change events? And does this differ concerning particular climate hazards, such as saltwater intrusion, flood or cyclone waters, or drought, and if so, why? | 7 | 67.0% |
| Water-related climate drivers | 27 | Do climate-induced water scarcity and droughts change individual preferences for disposable hygiene materials (e.g. nappies, menstrual pads, incontinence pads)? | 7 | 66.4% |
| Temperature-related climate drivers | 45 | Given the food contamination risks resulting from higher temperatures and extreme weather events, what messages should be included in the promotion of safe food hygiene practices? | 7 | 65.5% |
| Water-related climate drivers | 30 | How are budgets of national and local stakeholders being allocated for climate-resilient hygiene efforts and services? | 7 | 65.2% |
| Water-related climate drivers | 40 | What challenges are faced by women and girls concerning menstrual health and hygiene during extreme weather events, periods of climate-induced water scarcity, drought and saltwater intrusion? | 7 | 65.0% |
| Climate-related hazards | 15 | What are appropriate channels of communication to populations about hygiene-related health risks before, during and post extreme weather events? | 6 | 65.0% |
| Temperature-related climate drivers | 51 | How will higher ambient temperatures, humidity and precipitation change the contamination of food across the whole food chain, from preparation, processing, storage and consumption? | 6 | 64.3% |
| Water-related climate drivers | 38 | What are the gender-specific hygiene needs affected by climate change? | 7 | 62.7% |
| Temperature-related climate drivers | 47 | How do extreme heat events affect an individual's use and consumption of water for hygiene practices? | 7 | 62.5% |
| Water-related climate drivers | 26 | Are different approaches needed for hygiene promotion and hygiene behaviour change for populations living in water-scarce or drought conditions? | 7 | 62.3% |
| Climate-related hazards | 24 | What surfaces are most critical for cleaning during heavy rains and floods to maintain hygiene in the domestic environment? | 6 | 60.1% |
| Water-related climate drivers | 35 | How does the effect of climate change on women's and girls' hygiene affect their participation in wider society? | 7 | 60.0% |
| Water-related climate drivers | 34 | How does climate-induced water scarcity affect individual's experience of shame, embarrassment and humiliation due to their lack of opportunity for personal or menstrual hygiene? | 7 | 57.3% |
| Temperature-related climate drivers | 50 | How will extreme heat affect water collecting practices and the water available for hygiene practices? | 7 | 57.2% |
| Climate-related hazards | 25 | Which approaches are effective in preparing and restoring the hygiene supply chain to the population during extreme weather events? | 6 | 52.3% |
| Water-related climate drivers | 29 | Given climate-induced water scarcity and drought conditions, what options are there to conserve or recycle water at the household level for hygiene purposes? | 7 | 51.8% |
| Climate-related hazards | 3 | Given the microbial and chemical contamination risks of floodwater, what messages should be included in health promotion and hygiene behaviour change campaigns? | 6 | 45.5% |
| Climate-related hazards | 13 | How resilient are hygiene services (i.e., facilities, hardware, infrastructure) to the effects of climate change? | 6 | 42.5% |
| Temperature-related climate drivers | 52 | What effect will an increase in ambient temperature and humidity associated with climate change have on pathogen levels, with implications for human health, in water, food, soil, surfaces, and the environment? | 6 | 38.7% |
| Climate-related hazards | 10 | How do individuals prioritise hygiene in relation to other needs during extreme weather events? | 6 | 36.9% |
| Water-related climate drivers | 36 | How does the reduced opportunity to practice personal hygiene (washing, bathing, showering) affect mental health during extreme weather events or periods of climate-induced water scarcity and drought? | 7 | 34.0% |
| Climate-related hazards | 4 | How are private water service providers considering and planning for the potential increase in hygiene needs during extreme weather events? | 6 | 32.9% |
| Climate-related hazards | 5 | How can coordination and implementation of hygiene-related interventions during and post extreme weather events be improved? | 6 | 32.2% |
| Climate-related hazards | 17 | What are the health risks of using floodwater for personal and domestic hygiene? | 6 | 31.4% |
| Climate-related hazards | 22 | What microbiological and chemical contaminants from floods can be mitigated with effective domestic hygiene? | 6 | 29.6% |
| Temperature-related climate drivers | 53 | What is the association between higher ambient temperatures, precipitation variability and hand contamination? | 6 | 28.4% |
| Temperature-related climate drivers | 54 | What is the association between higher ambient temperatures, precipitation variability and hand-washing behaviours? | 6 | 27.1% |
| Temperature-related climate drivers | 57 | What is the stability and durability of hygiene products, (e.g., soap, detergents, cleaning products) under higher ambient temperature and humidity throughout the process of manufacturing, transportation and use? | 6 | 22.1% |
| Temperature-related climate drivers | 56 | What is the association between higher ambient temperatures, precipitation variability and the effectiveness of handwashing interventions against diarrhoeal disease? | 6 | 8.3% |
| Water-related climate drivers | 37 | How will climate change alter the risk of exposure to domestic animals and their waste, and how can this be reduced by hygiene in the domestic environment? | 7 | 0.0% |

| 6. Government | | | | |
| --- | --- | --- | --- | --- |
| **Climate Hazards** | **RQ ID #** | **Research Question** | **# of Respondents** | **Scaled Weighted RPS (%)** |
| Water-related climate drivers | 28 | Does climate-induced water scarcity or drought affect an individual's capability, opportunity or motivation to practice effective hygiene behaviours? | 5 | 100.0% |
| Climate-related hazards | 14 | How well are interventions addressing gender-specific hygiene needs during extreme weather events? | 5 | 99.6% |
| Water-related climate drivers | 26 | Are different approaches needed for hygiene promotion and hygiene behaviour change for populations living in water-scarce or drought conditions? | 5 | 99.3% |
| Climate-related hazards | 3 | Given the microbial and chemical contamination risks of floodwater, what messages should be included in health promotion and hygiene behaviour change campaigns? | 5 | 95.6% |
| Climate-related hazards | 9 | How do extreme weather events affect the capability of people with disabilities to maintain personal hygiene? | 5 | 95.0% |
| Water-related climate drivers | 42 | What is the association between extreme weather events, climate-induced water scarcity, drought or saltwater intrusion with the incidence of neglected tropical diseases (NTDs), and how are these risks mediated by hygiene practices? | 4 | 91.3% |
| Climate-related hazards | 21 | What hygiene promotion actions and preparedness strategies are needed before or during extreme weather events to prepare populations for increased risks to health? | 4 | 85.2% |
| Climate-related hazards | 8 | How do domestic hygiene practices during extreme weather events (droughts, heavy precipitation, floods and cyclones) change the risk of mosquito-borne diseases? | 5 | 83.1% |
| Temperature-related climate drivers | 46 | How do changes in ambient temperature, humidity and precipitation affect food safety and incidence of foodborne illness? | 5 | 82.9% |
| Water-related climate drivers | 32 | How do climate-induced water scarcity and drought affect individual's use and consumption of water for hygiene practices? | 5 | 82.7% |
| Temperature-related climate drivers | 50 | How will extreme heat affect water collecting practices and the water available for hygiene practices? | 5 | 82.7% |
| Water-related climate drivers | 29 | Given climate-induced water scarcity and drought conditions, what options are there to conserve or recycle water at the household level for hygiene purposes? | 5 | 82.2% |
| Temperature-related climate drivers | 49 | How will climate change affect the current Burden of Disease estimates related to hygiene? | 5 | 80.1% |
| Temperature-related climate drivers | 55 | What is the association between higher ambient temperatures, precipitation variability and humidity with the incidence of diarrhoeal diseases including cholera? | 5 | 78.8% |
| Climate-related hazards | 2 | During extreme weather events, how does solid waste contaminate the domestic environment and increase the exposure to infectious diseases and/or chemical pollutants? | 5 | 78.6% |
| Water-related climate drivers | 33 | How do climate-induced water scarcity, drought or precipitation variability affect the infectious disease burden, and how is this mediated by changes to hygiene practices? | 4 | 75.2% |
| Climate-related hazards | 17 | What are the health risks of using floodwater for personal and domestic hygiene? | 4 | 74.4% |
| Water-related climate drivers | 31 | How can effective behaviour change programmes for hygiene-related risks of climate change (e.g., floods, cyclones, climate-change induced water scarcity and drought) best be designed and delivered? | 5 | 74.4% |
| Climate-related hazards | 11 | How does acute disruption of water and sanitation services due to extreme weather events impact personal hygiene behaviours? | 5 | 73.1% |
| Temperature-related climate drivers | 47 | How do extreme heat events affect an individual's use and consumption of water for hygiene practices? | 5 | 70.0% |
| Climate-related hazards | 5 | How can coordination and implementation of hygiene-related interventions during and post extreme weather events be improved? | 5 | 69.3% |
| Climate-related hazards | 12 | How does chronic disruption of water and sanitation services due to extreme weather events impact personal hygiene behaviours? | 5 | 69.3% |
| Temperature-related climate drivers | 45 | Given the food contamination risks resulting from higher temperatures and extreme weather events, what messages should be included in the promotion of safe food hygiene practices? | 5 | 69.3% |
| Temperature-related climate drivers | 56 | What is the association between higher ambient temperatures, precipitation variability and the effectiveness of handwashing interventions against diarrhoeal disease? | 5 | 69.2% |
| Climate-related hazards | 23 | What support is needed for people with disabilities and their caregivers during extreme weather events to maintain personal hygiene? | 4 | 69.1% |
| Water-related climate drivers | 40 | What challenges are faced by women and girls concerning menstrual health and hygiene during extreme weather events, periods of climate-induced water scarcity, drought and saltwater intrusion? | 4 | 69.1% |
| Climate-related hazards | 7 | How can the hygiene needs of people with incontinence be addressed during extreme weather events? | 5 | 64.0% |
| Climate-related hazards | 19 | What culturally acceptable products are needed in disaster preparedness kits for personal hygiene? | 4 | 63.6% |
| Water-related climate drivers | 36 | How does the reduced opportunity to practice personal hygiene (washing, bathing, showering) affect mental health during extreme weather events or periods of climate-induced water scarcity and drought? | 4 | 62.1% |
| Temperature-related climate drivers | 44 | Do climate-induced cold spells or freezes affect an individual's capability, opportunity or motivation to practice effective hygiene behaviours? | 5 | 61.8% |
| Climate-related hazards | 6 | How can individuals, and those that support them, maintain menstrual health during extreme weather events? | 5 | 60.2% |
| Climate-related hazards | 22 | What microbiological and chemical contaminants from floods can be mitigated with effective domestic hygiene? | 4 | 58.5% |
| Climate-related hazards | 1 | Do people experiencing homelessness have specific hygiene-related challenges during extreme weather events? | 5 | 55.8% |
| Water-related climate drivers | 39 | What are the risks of vaginal or reproductive tract infections (RTIs), urinary tract infections (UTIs), fungal infections, bacterial vaginosis, rashes or discomfort due to inadequate menstrual hygiene from climate change events? And does this differ concerning particular climate hazards, such as saltwater intrusion, flood or cyclone waters, or drought, and if so, why? | 4 | 54.7% |
| Climate-related hazards | 16 | What are the determinants of hygiene behaviours during extreme weather events? | 4 | 53.0% |
| Temperature-related climate drivers | 51 | How will higher ambient temperatures, humidity and precipitation change the contamination of food across the whole food chain, from preparation, processing, storage and consumption? | 5 | 51.8% |
| Water-related climate drivers | 35 | How does the effect of climate change on women's and girls' hygiene affect their participation in wider society? | 4 | 51.0% |
| Climate-related hazards | 18 | What conditions for vectors and pests (e.g., mosquitoes, mice, cockroaches, and rats) during floods can be mitigated with effective domestic hygiene? | 4 | 49.0% |
| Temperature-related climate drivers | 48 | How does climate change affect the food preparation and hygiene practices of individuals, and how does this differ by climate events, such as heatwaves, high humidity, extreme weather events and/or climate-induced water scarcity? | 5 | 48.2% |
| Water-related climate drivers | 43 | What will be the effect on skin conditions from inadequate personal hygiene due to climate change? | 4 | 47.2% |
| Water-related climate drivers | 37 | How will climate change alter the risk of exposure to domestic animals and their waste, and how can this be reduced by hygiene in the domestic environment? | 4 | 47.2% |
| Temperature-related climate drivers | 52 | What effect will an increase in ambient temperature and humidity associated with climate change have on pathogen levels, with implications for human health, in water, food, soil, surfaces, and the environment? | 5 | 44.4% |
| Climate-related hazards | 10 | How do individuals prioritise hygiene in relation to other needs during extreme weather events? | 5 | 43.5% |
| Climate-related hazards | 4 | How are private water service providers considering and planning for the potential increase in hygiene needs during extreme weather events? | 5 | 43.3% |
| Temperature-related climate drivers | 54 | What is the association between higher ambient temperatures, precipitation variability and hand-washing behaviours? | 5 | 41.6% |
| Water-related climate drivers | 41 | What challenges are there in maintaining adequate personal hygiene during extreme weather events, periods of climate-induced water scarcity, drought and saltwater intrusion? | 4 | 36.3% |
| Water-related climate drivers | 38 | What are the gender-specific hygiene needs affected by climate change? | 4 | 36.1% |
| Water-related climate drivers | 34 | How does climate-induced water scarcity affect individual's experience of shame, embarrassment and humiliation due to their lack of opportunity for personal or menstrual hygiene? | 4 | 31.4% |
| Water-related climate drivers | 30 | How are budgets of national and local stakeholders being allocated for climate-resilient hygiene efforts and services? | 5 | 30.6% |
| Climate-related hazards | 24 | What surfaces are most critical for cleaning during heavy rains and floods to maintain hygiene in the domestic environment? | 4 | 30.5% |
| Climate-related hazards | 15 | What are appropriate channels of communication to populations about hygiene-related health risks before, during and post extreme weather events? | 4 | 28.2% |
| Water-related climate drivers | 27 | Do climate-induced water scarcity and droughts change individual preferences for disposable hygiene materials (e.g. nappies, menstrual pads, incontinence pads)? | 5 | 26.2% |
| Temperature-related climate drivers | 57 | What is the stability and durability of hygiene products, (e.g., soap, detergents, cleaning products) under higher ambient temperature and humidity throughout the process of manufacturing, transportation and use? | 5 | 25.5% |
| Climate-related hazards | 13 | How resilient are hygiene services (i.e., facilities, hardware, infrastructure) to the effects of climate change? | 5 | 25.3% |
| Climate-related hazards | 25 | Which approaches are effective in preparing and restoring the hygiene supply chain to the population during extreme weather events? | 4 | 18.3% |
| Climate-related hazards | 20 | What hygiene products are needed for post-flood cleaning of households and domestic spaces? | 4 | 3.5% |
| Temperature-related climate drivers | 53 | What is the association between higher ambient temperatures, precipitation variability and hand contamination? | 5 | 0.0% |

| 7. Private Sector* | | | | |
| --- | --- | --- | --- | --- |
| **Scaled Weighted RPS Percentages are overlapping due to low frequency of survey takers from this organisation type* | | | | |
| **Climate Hazards** | **RQ ID #** | **Research Question** | **# of Respondents** | **Scaled Weighted RPS (%)** |
| Climate-related hazards | 11 | How does acute disruption of water and sanitation services due to extreme weather events impact personal hygiene behaviours? | 4 | 100.0% |
| Climate-related hazards | 17 | What are the health risks of using floodwater for personal and domestic hygiene? | 4 | 100.0% |
| Climate-related hazards | 18 | What conditions for vectors and pests (e.g., mosquitoes, mice, cockroaches, and rats) during floods can be mitigated with effective domestic hygiene? | 4 | 100.0% |
| Water-related climate drivers | 32 | How do climate-induced water scarcity and drought affect individual's use and consumption of water for hygiene practices? | 4 | 100.0% |
| Temperature-related climate drivers | 50 | How will extreme heat affect water collecting practices and the water available for hygiene practices? | 4 | 100.0% |
| Climate-related hazards | 12 | How does chronic disruption of water and sanitation services due to extreme weather events impact personal hygiene behaviours? | 4 | 90.0% |
| Climate-related hazards | 24 | What surfaces are most critical for cleaning during heavy rains and floods to maintain hygiene in the domestic environment? | 4 | 90.0% |
| Temperature-related climate drivers | 55 | What is the association between higher ambient temperatures, precipitation variability and humidity with the incidence of diarrhoeal diseases including cholera? | 4 | 90.0% |
| Temperature-related climate drivers | 51 | How will higher ambient temperatures, humidity and precipitation change the contamination of food across the whole food chain, from preparation, processing, storage and consumption? | 4 | 81.0% |
| Temperature-related climate drivers | 56 | What is the association between higher ambient temperatures, precipitation variability and the effectiveness of handwashing interventions against diarrhoeal disease? | 4 | 80.5% |
| Climate-related hazards | 1 | Do people experiencing homelessness have specific hygiene-related challenges during extreme weather events? | 4 | 80.0% |
| Climate-related hazards | 3 | Given the microbial and chemical contamination risks of floodwater, what messages should be included in health promotion and hygiene behaviour change campaigns? | 4 | 80.0% |
| Water-related climate drivers | 42 | What is the association between extreme weather events, climate-induced water scarcity, drought or saltwater intrusion with the incidence of neglected tropical diseases (NTDs), and how are these risks mediated by hygiene practices? | 4 | 73.3% |
| Climate-related hazards | 14 | How well are interventions addressing gender-specific hygiene needs during extreme weather events? | 4 | 70.5% |
| Temperature-related climate drivers | 47 | How do extreme heat events affect an individual's use and consumption of water for hygiene practices? | 4 | 70.5% |
| Water-related climate drivers | 26 | Are different approaches needed for hygiene promotion and hygiene behaviour change for populations living in water-scarce or drought conditions? | 4 | 70.0% |
| Climate-related hazards | 2 | During extreme weather events, how does solid waste contaminate the domestic environment and increase the exposure to infectious diseases and/or chemical pollutants? | 4 | 69.5% |
| Temperature-related climate drivers | 45 | Given the food contamination risks resulting from higher temperatures and extreme weather events, what messages should be included in the promotion of safe food hygiene practices? | 4 | 69.5% |
| Water-related climate drivers | 39 | What are the risks of vaginal or reproductive tract infections (RTIs), urinary tract infections (UTIs), fungal infections, bacterial vaginosis, rashes or discomfort due to inadequate menstrual hygiene from climate change events? And, does this differ concerning particular climate hazards, such as saltwater intrusion, flood or cyclone waters, or drought, and if so, why? | 4 | 60.6% |
| Climate-related hazards | 6 | How can individuals, and those that support them, maintain menstrual health during extreme weather events? | 4 | 60.5% |
| Climate-related hazards | 25 | Which approaches are effective in preparing and restoring the hygiene supply chain to the population during extreme weather events? | 4 | 60.5% |
| Water-related climate drivers | 27 | Do climate-induced water scarcity and droughts change individual preferences for disposable hygiene materials (e.g. nappies, menstrual pads, incontinence pads)? | 4 | 60.5% |
| Water-related climate drivers | 28 | Does climate-induced water scarcity or drought affect an individual's capability, opportunity or motivation to practice effective hygiene behaviours? | 4 | 60.5% |
| Climate-related hazards | 16 | What are the determinants of hygiene behaviours during extreme weather events? | 4 | 60.0% |
| Water-related climate drivers | 29 | Given climate-induced water scarcity and drought conditions, what options are there to conserve or recycle water at the household level for hygiene purposes? | 4 | 60.0% |
| Water-related climate drivers | 41 | What challenges are there in maintaining adequate personal hygiene during extreme weather events, periods of climate-induced water scarcity, drought and saltwater intrusion? | 4 | 60.0% |
| Water-related climate drivers | 43 | What will be the effect on skin conditions from inadequate personal hygiene due to climate change? | 4 | 59.4% |
| Temperature-related climate drivers | 44 | Do climate-induced cold spells or freezes affect an individual's capability, opportunity or motivation to practice effective hygiene behaviours? | 4 | 59.4% |
| Climate-related hazards | 20 | What hygiene products are needed for post-flood cleaning of households and domestic spaces? | 4 | 58.7% |
| Climate-related hazards | 5 | How can coordination and implementation of hygiene-related interventions during and post extreme weather events be improved? | 4 | 50.5% |
| Climate-related hazards | 7 | How can the hygiene needs of people with incontinence be addressed during extreme weather events? | 4 | 50.5% |
| Water-related climate drivers | 31 | How can effective behaviour change programmes for hygiene-related risks of climate change (e.g., floods, cyclones, climate-change induced water scarcity and drought) best be designed and delivered? | 4 | 50.5% |
| Water-related climate drivers | 37 | How will climate change alter the risk of exposure to domestic animals and their waste, and how can this be reduced by hygiene in the domestic environment? | 4 | 50.5% |
| Temperature-related climate drivers | 57 | What is the stability and durability of hygiene products, (e.g., soap, detergents, cleaning products) under higher ambient temperature and humidity throughout the process of manufacturing, transportation and use? | 4 | 50.5% |
| Climate-related hazards | 21 | What hygiene promotion actions and preparedness strategies are needed before or during extreme weather events to prepare populations for increased risks to health? | 4 | 49.5% |
| Climate-related hazards | 10 | How do individuals prioritise hygiene in relation to other needs during extreme weather events? | 4 | 49.0% |
| Water-related climate drivers | 34 | How does climate-induced water scarcity affect individual's experience of shame, embarrassment and humiliation due to their lack of opportunity for personal or menstrual hygiene? | 4 | 41.0% |
| Climate-related hazards | 15 | What are appropriate channels of communication to populations about hygiene-related health risks before, during and post extreme weather events? | 4 | 49.0% |
| Climate-related hazards | 23 | What support is needed for people with disabilities and their caregivers during extreme weather events to maintain personal hygiene? | 4 | 48.2% |
| Water-related climate drivers | 36 | How does the reduced opportunity to practice personal hygiene (washing, bathing, showering) affect mental health during extreme weather events or periods of climate-induced water scarcity and drought? | 4 | 47.2% |
| Water-related climate drivers | 33 | How do climate-induced water scarcity, drought or precipitation variability affect the infectious disease burden, and how is this mediated by changes to hygiene practices? | 4 | 47.2% |
| Climate-related hazards | 8 | How do domestic hygiene practices during extreme weather events (droughts, heavy precipitation, floods and cyclones) change the risk of mosquito-borne diseases? | 4 | 44.4% |
| Temperature-related climate drivers | 46 | How do changes in ambient temperature, humidity and precipitation affect food safety and incidence of foodborne illness? | 4 | 43.5% |
| Climate-related hazards | 4 | How are private water service providers considering and planning for the potential increase in hygiene needs during extreme weather events? | 4 | 43.3% |
| Water-related climate drivers | 35 | How does the effect of climate change on women's and girls' hygiene affect their participation in wider society? | 4 | 41.6% |
| Climate-related hazards | 22 | What microbiological and chemical contaminants from floods can be mitigated with effective domestic hygiene? | 4 | 36.3% |
| Temperature-related climate drivers | 48 | How does climate change affect the food preparation and hygiene practices of individuals, and how does this differ by climate events, such as heatwaves, high humidity, extreme weather events and/or climate-induced water scarcity? | 4 | 36.1% |
| Temperature-related climate drivers | 49 | How will climate change affect the current Burden of Disease estimates related to hygiene? | 4 | 31.4% |
| Temperature-related climate drivers | 54 | What is the association between higher ambient temperatures, precipitation variability and hand-washing behaviours? | 4 | 30.6% |
| Climate-related hazards | 13 | How resilient are hygiene services (i.e., facilities, hardware, infrastructure) to the effects of climate change? | 4 | 30.5% |
| Water-related climate drivers | 38 | What are the gender-specific hygiene needs affected by climate change? | 4 | 28.2% |
| Climate-related hazards | 9 | How do extreme weather events affect the capability of people with disabilities to maintain personal hygiene? | 4 | 26.2% |
| Temperature-related climate drivers | 52 | What effect will an increase in ambient temperature and humidity associated with climate change have on pathogen levels, with implications for human health, in water, food, soil, surfaces, and the environment? | 4 | 25.5% |
| Water-related climate drivers | 30 | How are budgets of national and local stakeholders being allocated for climate-resilient hygiene efforts and services? | 4 | 25.3% |
| Water-related climate drivers | 40 | What challenges are faced by women and girls concerning menstrual health and hygiene during extreme weather events, periods of climate-induced water scarcity, drought and saltwater intrusion? | 4 | 18.3% |
| Temperature-related climate drivers | 53 | What is the association between higher ambient temperatures, precipitation variability and hand contamination? | 4 | 3.5% |
| Climate-related hazards | 19 | What culturally acceptable products are needed in disaster preparedness kits for personal hygiene? | 4 | 0.0% |

**Donor government agency is unable to produce scaled weighted RPS (%) and accordingly, ranking of research question priorities due to low frequency of count for translation (n=2)

### Table S2 – Research priorities stratified by region of focus

| 1. African Region (AFRO) | | | | |
| --- | --- | --- | --- | --- |
| **Climate Hazards** | **RQ ID #** | **Research Question** | **# of Respondents** | **Scaled Weighted RPS (%)** |
| Water-related climate drivers | 33 | How do climate-induced water scarcity, drought or precipitation variability affect the infectious disease burden, and how is this mediated by changes to hygiene practices? | 67 | 100.0% |
| Climate-related hazards | 21 | What hygiene promotion actions and preparedness strategies are needed before or during extreme weather events to prepare populations for increased risks to health? | 65 | 90.2% |
| Water-related climate drivers | 32 | How do climate-induced water scarcity and drought affect individual's use and consumption of water for hygiene practices? | 69 | 90.1% |
| Climate-related hazards | 23 | What support is needed for people with disabilities and their caregivers during extreme weather events to maintain personal hygiene? | 63 | 89.7% |
| Temperature-related climate drivers | 55 | What is the association between higher ambient temperatures, precipitation variability and humidity with the incidence of diarrhoeal diseases including cholera? | 69 | 87.7% |
| Water-related climate drivers | 40 | What challenges are faced by women and girls concerning menstrual health and hygiene during extreme weather events, periods of climate-induced water scarcity, drought and saltwater intrusion? | 66 | 86.2% |
| Temperature-related climate drivers | 46 | How do changes in ambient temperature, humidity and precipitation affect food safety and incidence of foodborne illness? | 73 | 85.8% |
| Climate-related hazards | 3 | Given the microbial and chemical contamination risks of floodwater, what messages should be included in health promotion and hygiene behaviour change campaigns? | 74 | 85.2% |
| Temperature-related climate drivers | 47 | How do extreme heat events affect an individual's use and consumption of water for hygiene practices? | 73 | 84.6% |
| Water-related climate drivers | 28 | Does climate-induced water scarcity or drought affect an individual's capability, opportunity or motivation to practice effective hygiene behaviours? | 69 | 82.0% |
| Temperature-related climate drivers | 45 | Given the food contamination risks resulting from higher temperatures and extreme weather events, what messages should be included in the promotion of safe food hygiene practices? | 73 | 82.0% |
| Climate-related hazards | 16 | What are the determinants of hygiene behaviours during extreme weather events? | 64 | 80.5% |
| Water-related climate drivers | 35 | How does the effect of climate change on women's and girls' hygiene affect their participation in wider society? | 67 | 79.5% |
| Water-related climate drivers | 29 | Given climate-induced water scarcity and drought conditions, what options are there to conserve or recycle water at the household level for hygiene purposes? | 69 | 79.5% |
| Climate-related hazards | 9 | How do extreme weather events affect the capability of people with disabilities to maintain personal hygiene? | 69 | 78.8% |
| Water-related climate drivers | 26 | Are different approaches needed for hygiene promotion and hygiene behaviour change for populations living in water-scarce or drought conditions? | 69 | 78.8% |
| Water-related climate drivers | 42 | What is the association between extreme weather events, climate-induced water scarcity, drought or saltwater intrusion with the incidence of neglected tropical diseases (NTDs), and how are these risks mediated by hygiene practices? | 66 | 78.4% |
| Temperature-related climate drivers | 52 | What effect will an increase in ambient temperature and humidity associated with climate change have on pathogen levels, with implications for human health, in water, food, soil, surfaces, and the environment? | 69 | 76.9% |
| Water-related climate drivers | 36 | How does the reduced opportunity to practice personal hygiene (washing, bathing, showering) affect mental health during extreme weather events or periods of climate-induced water scarcity and drought? | 67 | 76.1% |
| Climate-related hazards | 14 | How well are interventions addressing gender-specific hygiene needs during extreme weather events? | 69 | 75.8% |
| Climate-related hazards | 2 | During extreme weather events, how does solid waste contaminate the domestic environment and increase the exposure to infectious diseases and/or chemical pollutants? | 74 | 75.4% |
| Climate-related hazards | 12 | How does chronic disruption of water and sanitation services due to extreme weather events impact personal hygiene behaviours? | 69 | 75.2% |
| Climate-related hazards | 11 | How does acute disruption of water and sanitation services due to extreme weather events impact personal hygiene behaviours? | 69 | 71.8% |
| Climate-related hazards | 6 | How can individuals, and those that support them, maintain menstrual health during extreme weather events? | 73 | 71.2% |
| Climate-related hazards | 15 | What are appropriate channels of communication to populations about hygiene-related health risks before, during and post extreme weather events? | 65 | 69.8% |
| Temperature-related climate drivers | 50 | How will extreme heat affect water collecting practices and the water available for hygiene practices? | 73 | 69.7% |
| Climate-related hazards | 19 | What culturally acceptable products are needed in disaster preparedness kits for personal hygiene? | 65 | 67.5% |
| Water-related climate drivers | 41 | What challenges are there in maintaining adequate personal hygiene during extreme weather events, periods of climate-induced water scarcity, drought and saltwater intrusion? | 66 | 67.2% |
| Climate-related hazards | 7 | How can the hygiene needs of people with incontinence be addressed during extreme weather events? | 74 | 67.0% |
| Climate-related hazards | 13 | How resilient are hygiene services (i.e., facilities, hardware, infrastructure) to the effects of climate change? | 69 | 64.4% |
| Climate-related hazards | 5 | How can coordination and implementation of hygiene-related interventions during and post extreme weather events be improved? | 74 | 61.6% |
| Climate-related hazards | 25 | Which approaches are effective in preparing and restoring the hygiene supply chain to the population during extreme weather events? | 63 | 61.3% |
| Water-related climate drivers | 39 | What are the risks of vaginal or reproductive tract infections (RTIs), urinary tract infections (UTIs), fungal infections, bacterial vaginosis, rashes or discomfort due to inadequate menstrual hygiene from climate change events? And does this differ concerning particular climate hazards, such as saltwater intrusion, flood or cyclone waters, or drought, and if so, why? | 66 | 60.8% |
| Temperature-related climate drivers | 49 | How will climate change affect the current Burden of Disease estimates related to hygiene? | 73 | 60.8% |
| Climate-related hazards | 8 | How do domestic hygiene practices during extreme weather events (droughts, heavy precipitation, floods and cyclones) change the risk of mosquito-borne diseases? | 69 | 60.4% |
| Climate-related hazards | 18 | What conditions for vectors and pests (e.g., mosquitoes, mice, cockroaches, and rats) during floods can be mitigated with effective domestic hygiene? | 65 | 60.1% |
| Water-related climate drivers | 31 | How can effective behaviour change programmes for hygiene-related risks of climate change (e.g., floods, cyclones, climate-change induced water scarcity and drought) best be designed and delivered? | 69 | 59.6% |
| Temperature-related climate drivers | 51 | How will higher ambient temperatures, humidity and precipitation change the contamination of food across the whole food chain, from preparation, processing, storage and consumption? | 69 | 58.4% |
| Climate-related hazards | 10 | How do individuals prioritise hygiene in relation to other needs during extreme weather events? | 69 | 58.0% |
| Water-related climate drivers | 43 | What will be the effect on skin conditions from inadequate personal hygiene due to climate change? | 66 | 56.0% |
| Climate-related hazards | 17 | What are the health risks of using floodwater for personal and domestic hygiene? | 65 | 52.4% |
| Water-related climate drivers | 38 | What are the gender-specific hygiene needs affected by climate change? | 67 | 51.3% |
| Climate-related hazards | 1 | Do people experiencing homelessness have specific hygiene-related challenges during extreme weather events? | 74 | 49.3% |
| Temperature-related climate drivers | 48 | How does climate change affect the food preparation and hygiene practices of individuals, and how does this differ by climate events, such as heatwaves, high humidity, extreme weather events and/or climate-induced water scarcity? | 72 | 49.2% |
| Climate-related hazards | 22 | What microbiological and chemical contaminants from floods can be mitigated with effective domestic hygiene? | 63 | 41.8% |
| Water-related climate drivers | 34 | How does climate-induced water scarcity affect individual's experience of shame, embarrassment and humiliation due to their lack of opportunity for personal or menstrual hygiene? | 67 | 38.8% |
| Temperature-related climate drivers | 44 | Do climate-induced cold spells or freezes affect an individual's capability, opportunity or motivation to practice effective hygiene behaviours? | 73 | 38.0% |
| Temperature-related climate drivers | 56 | What is the association between higher ambient temperatures, precipitation variability and the effectiveness of handwashing interventions against diarrhoeal disease? | 68 | 34.1% |
| Water-related climate drivers | 30 | How are budgets of national and local stakeholders being allocated for climate-resilient hygiene efforts and services? | 68 | 34.0% |
| Water-related climate drivers | 37 | How will climate change alter the risk of exposure to domestic animals and their waste, and how can this be reduced by hygiene in the domestic environment? | 67 | 31.9% |
| Water-related climate drivers | 27 | Do climate-induced water scarcity and droughts change individual preferences for disposable hygiene materials (e.g. nappies, menstrual pads, incontinence pads)? | 69 | 31.5% |
| Temperature-related climate drivers | 57 | What is the stability and durability of hygiene products, (e.g., soap, detergents, cleaning products) under higher ambient temperature and humidity throughout the process of manufacturing, transportation and use? | 68 | 22.6% |
| Climate-related hazards | 24 | What surfaces are most critical for cleaning during heavy rains and floods to maintain hygiene in the domestic environment? | 63 | 20.2% |
| Climate-related hazards | 20 | What hygiene products are needed for post-flood cleaning of households and domestic spaces? | 65 | 19.4% |
| Climate-related hazards | 4 | How are private water service providers considering and planning for the potential increase in hygiene needs during extreme weather events? | 74 | 18.1% |
| Temperature-related climate drivers | 54 | What is the association between higher ambient temperatures, precipitation variability and hand-washing behaviours? | 69 | 6.4% |
| Temperature-related climate drivers | 53 | What is the association between higher ambient temperatures, precipitation variability and hand contamination? | 69 | 0.0% |

| 2. Region of the Americas (AMRO) | | | | |
| --- | --- | --- | --- | --- |
| **Climate Hazards** | **RQ ID #** | **Research Question** | **# of Respondents** | **Scaled Weighted RPS (%)** |
| Water-related climate drivers | 40 | What challenges are faced by women and girls concerning menstrual health and hygiene during extreme weather events, periods of climate-induced water scarcity, drought and saltwater intrusion? | 17 | 100.0% |
| Climate-related hazards | 21 | What hygiene promotion actions and preparedness strategies are needed before or during extreme weather events to prepare populations for increased risks to health? | 16 | 98.4% |
| Water-related climate drivers | 41 | What challenges are there in maintaining adequate personal hygiene during extreme weather events, periods of climate-induced water scarcity, drought and saltwater intrusion? | 17 | 96.3% |
| Water-related climate drivers | 33 | How do climate-induced water scarcity, drought or precipitation variability affect the infectious disease burden, and how is this mediated by changes to hygiene practices? | 17 | 95.9% |
| Water-related climate drivers | 26 | Are different approaches needed for hygiene promotion and hygiene behaviour change for populations living in water-scarce or drought conditions? | 17 | 95.6% |
| Climate-related hazards | 3 | Given the microbial and chemical contamination risks of floodwater, what messages should be included in health promotion and hygiene behaviour change campaigns? | 16 | 95.5% |
| Temperature-related climate drivers | 50 | How will extreme heat affect water collecting practices and the water available for hygiene practices? | 17 | 93.7% |
| Climate-related hazards | 10 | How do individuals prioritise hygiene in relation to other needs during extreme weather events? | 16 | 93.2% |
| Climate-related hazards | 1 | Do people experiencing homelessness have specific hygiene-related challenges during extreme weather events? | 16 | 92.8% |
| Climate-related hazards | 11 | How does acute disruption of water and sanitation services due to extreme weather events impact personal hygiene behaviours? | 16 | 92.6% |
| Climate-related hazards | 16 | What are the determinants of hygiene behaviours during extreme weather events? | 16 | 89.3% |
| Water-related climate drivers | 32 | How do climate-induced water scarcity and drought affect individual's use and consumption of water for hygiene practices? | 17 | 89.0% |
| Temperature-related climate drivers | 47 | How do extreme heat events affect an individual's use and consumption of water for hygiene practices? | 17 | 87.2% |
| Water-related climate drivers | 28 | Does climate-induced water scarcity or drought affect an individual's capability, opportunity or motivation to practice effective hygiene behaviours? | 17 | 86.6% |
| Water-related climate drivers | 36 | How does the reduced opportunity to practice personal hygiene (washing, bathing, showering) affect mental health during extreme weather events or periods of climate-induced water scarcity and drought? | 17 | 85.0% |
| Climate-related hazards | 13 | How resilient are hygiene services (i.e., facilities, hardware, infrastructure) to the effects of climate change? | 16 | 83.1% |
| Climate-related hazards | 19 | What culturally acceptable products are needed in disaster preparedness kits for personal hygiene? | 16 | 82.4% |
| Temperature-related climate drivers | 55 | What is the association between higher ambient temperatures, precipitation variability and humidity with the incidence of diarrhoeal diseases including cholera? | 16 | 82.4% |
| Water-related climate drivers | 42 | What is the association between extreme weather events, climate-induced water scarcity, drought or saltwater intrusion with the incidence of neglected tropical diseases (NTDs), and how are these risks mediated by hygiene practices? | 17 | 82.0% |
| Climate-related hazards | 2 | During extreme weather events, how does solid waste contaminate the domestic environment and increase the exposure to infectious diseases and/or chemical pollutants? | 16 | 81.5% |
| Climate-related hazards | 9 | How do extreme weather events affect the capability of people with disabilities to maintain personal hygiene? | 16 | 80.5% |
| Climate-related hazards | 12 | How does chronic disruption of water and sanitation services due to extreme weather events impact personal hygiene behaviours? | 16 | 79.7% |
| Water-related climate drivers | 29 | Given climate-induced water scarcity and drought conditions, what options are there to conserve or recycle water at the household level for hygiene purposes? | 17 | 78.7% |
| Temperature-related climate drivers | 51 | How will higher ambient temperatures, humidity and precipitation change the contamination of food across the whole food chain, from preparation, processing, storage and consumption? | 16 | 77.5% |
| Water-related climate drivers | 35 | How does the effect of climate change on women's and girls' hygiene affect their participation in wider society? | 17 | 76.4% |
| Temperature-related climate drivers | 52 | What effect will an increase in ambient temperature and humidity associated with climate change have on pathogen levels, with implications for human health, in water, food, soil, surfaces, and the environment? | 16 | 76.0% |
| Temperature-related climate drivers | 46 | How do changes in ambient temperature, humidity and precipitation affect food safety and incidence of foodborne illness? | 17 | 74.4% |
| Climate-related hazards | 15 | What are appropriate channels of communication to populations about hygiene-related health risks before, during and post extreme weather events? | 16 | 74.3% |
| Climate-related hazards | 25 | Which approaches are effective in preparing and restoring the hygiene supply chain to the population during extreme weather events? | 16 | 72.1% |
| Climate-related hazards | 23 | What support is needed for people with disabilities and their caregivers during extreme weather events to maintain personal hygiene? | 16 | 69.7% |
| Climate-related hazards | 17 | What are the health risks of using floodwater for personal and domestic hygiene? | 16 | 68.9% |
| Climate-related hazards | 14 | How well are interventions addressing gender-specific hygiene needs during extreme weather events? | 16 | 67.3% |
| Climate-related hazards | 6 | How can individuals, and those that support them, maintain menstrual health during extreme weather events? | 16 | 67.2% |
| Water-related climate drivers | 39 | What are the risks of vaginal or reproductive tract infections (RTIs), urinary tract infections (UTIs), fungal infections, bacterial vaginosis, rashes or discomfort due to inadequate menstrual hygiene from climate change events? And does this differ concerning particular climate hazards, such as saltwater intrusion, flood or cyclone waters, or drought, and if so, why? | 17 | 64.5% |
| Water-related climate drivers | 31 | How can effective behaviour change programmes for hygiene-related risks of climate change (e.g., floods, cyclones, climate-change induced water scarcity and drought) best be designed and delivered? | 17 | 63.9% |
| Climate-related hazards | 20 | What hygiene products are needed for post-flood cleaning of households and domestic spaces? | 16 | 61.0% |
| Temperature-related climate drivers | 45 | Given the food contamination risks resulting from higher temperatures and extreme weather events, what messages should be included in the promotion of safe food hygiene practices? | 17 | 61.0% |
| Climate-related hazards | 7 | How can the hygiene needs of people with incontinence be addressed during extreme weather events? | 16 | 59.6% |
| Water-related climate drivers | 34 | How does climate-induced water scarcity affect individual's experience of shame, embarrassment and humiliation due to their lack of opportunity for personal or menstrual hygiene? | 17 | 58.3% |
| Climate-related hazards | 18 | What conditions for vectors and pests (e.g., mosquitoes, mice, cockroaches, and rats) during floods can be mitigated with effective domestic hygiene? | 16 | 58.0% |
| Water-related climate drivers | 37 | How will climate change alter the risk of exposure to domestic animals and their waste, and how can this be reduced by hygiene in the domestic environment? | 17 | 57.2% |
| Climate-related hazards | 8 | How do domestic hygiene practices during extreme weather events (droughts, heavy precipitation, floods and cyclones) change the risk of mosquito-borne diseases? | 16 | 54.9% |
| Climate-related hazards | 5 | How can coordination and implementation of hygiene-related interventions during and post extreme weather events be improved? | 16 | 52.7% |
| Climate-related hazards | 22 | What microbiological and chemical contaminants from floods can be mitigated with effective domestic hygiene? | 16 | 51.6% |
| Water-related climate drivers | 38 | What are the gender-specific hygiene needs affected by climate change? | 17 | 51.5% |
| Temperature-related climate drivers | 48 | How does climate change affect the food preparation and hygiene practices of individuals, and how does this differ by climate events, such as heatwaves, high humidity, extreme weather events and/or climate-induced water scarcity? | 17 | 50.4% |
| Water-related climate drivers | 27 | Do climate-induced water scarcity and droughts change individual preferences for disposable hygiene materials (e.g. nappies, menstrual pads, incontinence pads)? | 17 | 41.2% |
| Climate-related hazards | 4 | How are private water service providers considering and planning for the potential increase in hygiene needs during extreme weather events? | 16 | 41.0% |
| Temperature-related climate drivers | 56 | What is the association between higher ambient temperatures, precipitation variability and the effectiveness of handwashing interventions against diarrhoeal disease? | 16 | 39.1% |
| Temperature-related climate drivers | 49 | How will climate change affect the current Burden of Disease estimates related to hygiene? | 17 | 33.3% |
| Climate-related hazards | 24 | What surfaces are most critical for cleaning during heavy rains and floods to maintain hygiene in the domestic environment? | 16 | 31.5% |
| Water-related climate drivers | 43 | What will be the effect on skin conditions from inadequate personal hygiene due to climate change? | 17 | 30.9% |
| Temperature-related climate drivers | 44 | Do climate-induced cold spells or freezes affect an individual's capability, opportunity or motivation to practice effective hygiene behaviours? | 17 | 28.7% |
| Water-related climate drivers | 30 | How are budgets of national and local stakeholders being allocated for climate-resilient hygiene efforts and services? | 17 | 22.4% |
| Temperature-related climate drivers | 57 | What is the stability and durability of hygiene products, (e.g., soap, detergents, cleaning products) under higher ambient temperature and humidity throughout the process of manufacturing, transportation and use? | 16 | 21.7% |
| Temperature-related climate drivers | 54 | What is the association between higher ambient temperatures, precipitation variability and hand-washing behaviours? | 16 | 3.8% |
| Temperature-related climate drivers | 53 | What is the association between higher ambient temperatures, precipitation variability and hand contamination? | 16 | 0.0% |

| 3. South-East Asia Region (SEARO) | | | | |
| --- | --- | --- | --- | --- |
| **Climate Hazards** | **RQ ID #** | **Research Question** | **# of Respondents** | **Scaled Weighted RPS (%)** |
| Water-related climate drivers | 33 | How do climate-induced water scarcity, drought or precipitation variability affect the infectious disease burden, and how is this mediated by changes to hygiene practices? | 36 | 100.0% |
| Water-related climate drivers | 42 | What is the association between extreme weather events, climate-induced water scarcity, drought or saltwater intrusion with the incidence of neglected tropical diseases (NTDs), and how are these risks mediated by hygiene practices? | 36 | 95.1% |
| Climate-related hazards | 23 | What support is needed for people with disabilities and their caregivers during extreme weather events to maintain personal hygiene? | 37 | 94.5% |
| Temperature-related climate drivers | 55 | What is the association between higher ambient temperatures, precipitation variability and humidity with the incidence of diarrhoeal diseases including cholera? | 36 | 92.3% |
| Water-related climate drivers | 28 | Does climate-induced water scarcity or drought affect an individual's capability, opportunity or motivation to practice effective hygiene behaviours? | 36 | 90.0% |
| Climate-related hazards | 21 | What hygiene promotion actions and preparedness strategies are needed before or during extreme weather events to prepare populations for increased risks to health? | 38 | 88.5% |
| Temperature-related climate drivers | 47 | How do extreme heat events affect an individual's use and consumption of water for hygiene practices? | 38 | 88.0% |
| Water-related climate drivers | 41 | What challenges are there in maintaining adequate personal hygiene during extreme weather events, periods of climate-induced water scarcity, drought and saltwater intrusion? | 36 | 88.0% |
| Water-related climate drivers | 26 | Are different approaches needed for hygiene promotion and hygiene behaviour change for populations living in water-scarce or drought conditions? | 36 | 87.7% |
| Temperature-related climate drivers | 46 | How do changes in ambient temperature, humidity and precipitation affect food safety and incidence of foodborne illness? | 38 | 87.3% |
| Water-related climate drivers | 40 | What challenges are faced by women and girls concerning menstrual health and hygiene during extreme weather events, periods of climate-induced water scarcity, drought and saltwater intrusion? | 36 | 86.7% |
| Water-related climate drivers | 32 | How do climate-induced water scarcity and drought affect individual's use and consumption of water for hygiene practices? | 36 | 85.7% |
| Climate-related hazards | 14 | How well are interventions addressing gender-specific hygiene needs during extreme weather events? | 39 | 83.7% |
| Temperature-related climate drivers | 52 | What effect will an increase in ambient temperature and humidity associated with climate change have on pathogen levels, with implications for human health, in water, food, soil, surfaces, and the environment? | 36 | 82.7% |
| Climate-related hazards | 9 | How do extreme weather events affect the capability of people with disabilities to maintain personal hygiene? | 39 | 82.5% |
| Climate-related hazards | 13 | How resilient are hygiene services (i.e., facilities, hardware, infrastructure) to the effects of climate change? | 39 | 81.7% |
| Temperature-related climate drivers | 50 | How will extreme heat affect water collecting practices and the water available for hygiene practices? | 38 | 81.0% |
| Climate-related hazards | 2 | During extreme weather events, how does solid waste contaminate the domestic environment and increase the exposure to infectious diseases and/or chemical pollutants? | 40 | 80.9% |
| Climate-related hazards | 3 | Given the microbial and chemical contamination risks of floodwater, what messages should be included in health promotion and hygiene behaviour change campaigns? | 39 | 78.4% |
| Water-related climate drivers | 35 | How does the effect of climate change on women's and girls' hygiene affect their participation in wider society? | 36 | 78.1% |
| Climate-related hazards | 11 | How does acute disruption of water and sanitation services due to extreme weather events impact personal hygiene behaviours? | 39 | 75.9% |
| Climate-related hazards | 12 | How does chronic disruption of water and sanitation services due to extreme weather events impact personal hygiene behaviours? | 39 | 75.0% |
| Climate-related hazards | 16 | What are the determinants of hygiene behaviours during extreme weather events? | 37 | 74.9% |
| Water-related climate drivers | 29 | Given climate-induced water scarcity and drought conditions, what options are there to conserve or recycle water at the household level for hygiene purposes? | 36 | 74.5% |
| Temperature-related climate drivers | 45 | Given the food contamination risks resulting from higher temperatures and extreme weather events, what messages should be included in the promotion of safe food hygiene practices? | 38 | 73.1% |
| Climate-related hazards | 6 | How can individuals, and those that support them, maintain menstrual health during extreme weather events? | 39 | 73.0% |
| Climate-related hazards | 8 | How do domestic hygiene practices during extreme weather events (droughts, heavy precipitation, floods and cyclones) change the risk of mosquito-borne diseases? | 39 | 71.7% |
| Climate-related hazards | 18 | What conditions for vectors and pests (e.g., mosquitoes, mice, cockroaches, and rats) during floods can be mitigated with effective domestic hygiene? | 38 | 70.4% |
| Climate-related hazards | 19 | What culturally acceptable products are needed in disaster preparedness kits for personal hygiene? | 38 | 68.8% |
| Climate-related hazards | 15 | What are appropriate channels of communication to populations about hygiene-related health risks before, during and post extreme weather events? | 38 | 67.9% |
| Climate-related hazards | 25 | Which approaches are effective in preparing and restoring the hygiene supply chain to the population during extreme weather events? | 37 | 67.6% |
| Water-related climate drivers | 38 | What are the gender-specific hygiene needs affected by climate change? | 36 | 67.3% |
| Climate-related hazards | 1 | Do people experiencing homelessness have specific hygiene-related challenges during extreme weather events? | 38 | 65.8% |
| Climate-related hazards | 7 | How can the hygiene needs of people with incontinence be addressed during extreme weather events? | 39 | 65.7% |
| Water-related climate drivers | 36 | How does the reduced opportunity to practice personal hygiene (washing, bathing, showering) affect mental health during extreme weather events or periods of climate-induced water scarcity and drought? | 36 | 65.0% |
| Climate-related hazards | 5 | How can coordination and implementation of hygiene-related interventions during and post extreme weather events be improved? | 39 | 64.5% |
| Water-related climate drivers | 37 | How will climate change alter the risk of exposure to domestic animals and their waste, and how can this be reduced by hygiene in the domestic environment? | 36 | 64.4% |
| Water-related climate drivers | 31 | How can effective behaviour change programmes for hygiene-related risks of climate change (e.g., floods, cyclones, climate-change induced water scarcity and drought) best be designed and delivered? | 36 | 62.9% |
| Temperature-related climate drivers | 49 | How will climate change affect the current Burden of Disease estimates related to hygiene? | 38 | 61.3% |
| Water-related climate drivers | 39 | What are the risks of vaginal or reproductive tract infections (RTIs), urinary tract infections (UTIs), fungal infections, bacterial vaginosis, rashes or discomfort due to inadequate menstrual hygiene from climate change events? And does this differ concerning particular climate hazards, such as saltwater intrusion, flood or cyclone waters, or drought, and if so, why? | 36 | 57.1% |
| Temperature-related climate drivers | 51 | How will higher ambient temperatures, humidity and precipitation change the contamination of food across the whole food chain, from preparation, processing, storage and consumption? | 36 | 55.6% |
| Temperature-related climate drivers | 48 | How does climate change affect the food preparation and hygiene practices of individuals, and how does this differ by climate events, such as heatwaves, high humidity, extreme weather events and/or climate-induced water scarcity? | 38 | 54.9% |
| Climate-related hazards | 22 | What microbiological and chemical contaminants from floods can be mitigated with effective domestic hygiene? | 37 | 53.0% |
| Climate-related hazards | 17 | What are the health risks of using floodwater for personal and domestic hygiene? | 38 | 49.8% |
| Climate-related hazards | 10 | How do individuals prioritise hygiene in relation to other needs during extreme weather events? | 39 | 47.8% |
| Water-related climate drivers | 27 | Do climate-induced water scarcity and droughts change individual preferences for disposable hygiene materials (e.g. nappies, menstrual pads, incontinence pads)? | 36 | 47.6% |
| Water-related climate drivers | 43 | What will be the effect on skin conditions from inadequate personal hygiene due to climate change? | 36 | 45.0% |
| Water-related climate drivers | 30 | How are budgets of national and local stakeholders being allocated for climate-resilient hygiene efforts and services? | 36 | 42.0% |
| Climate-related hazards | 24 | What surfaces are most critical for cleaning during heavy rains and floods to maintain hygiene in the domestic environment? | 37 | 39.0% |
| Temperature-related climate drivers | 44 | Do climate-induced cold spells or freezes affect an individual's capability, opportunity or motivation to practice effective hygiene behaviours? | 38 | 38.9% |
| Climate-related hazards | 20 | What hygiene products are needed for post-flood cleaning of households and domestic spaces? | 38 | 37.4% |
| Temperature-related climate drivers | 57 | What is the stability and durability of hygiene products, (e.g., soap, detergents, cleaning products) under higher ambient temperature and humidity throughout the process of manufacturing, transportation and use? | 36 | 36.5% |
| Temperature-related climate drivers | 56 | What is the association between higher ambient temperatures, precipitation variability and the effectiveness of handwashing interventions against diarrhoeal disease? | 36 | 36.0% |
| Water-related climate drivers | 34 | How does climate-induced water scarcity affect individual's experience of shame, embarrassment and humiliation due to their lack of opportunity for personal or menstrual hygiene? | 36 | 32.3% |
| Climate-related hazards | 4 | How are private water service providers considering and planning for the potential increase in hygiene needs during extreme weather events? | 39 | 32.0% |
| Temperature-related climate drivers | 54 | What is the association between higher ambient temperatures, precipitation variability and hand-washing behaviours? | 36 | 2.1% |
| Temperature-related climate drivers | 53 | What is the association between higher ambient temperatures, precipitation variability and hand contamination? | 36 | 0.0% |

| 4. European Region (EURO) | | | | |
| --- | --- | --- | --- | --- |
| **Climate Hazards** | **RQ ID #** | **Research Question** | **# of Respondents** | **Scaled Weighted RPS (%)** |
| Climate-related hazards | 21 | What hygiene promotion actions and preparedness strategies are needed before or during extreme weather events to prepare populations for increased risks to health? | 16 | 100.0% |
| Climate-related hazards | 18 | What conditions for vectors and pests (e.g., mosquitoes, mice, cockroaches, and rats) during floods can be mitigated with effective domestic hygiene? | 16 | 99.6% |
| Temperature-related climate drivers | 51 | How will higher ambient temperatures, humidity and precipitation change the contamination of food across the whole food chain, from preparation, processing, storage and consumption? | 16 | 98.6% |
| Temperature-related climate drivers | 55 | What is the association between higher ambient temperatures, precipitation variability and humidity with the incidence of diarrhoeal diseases including cholera? | 16 | 98.1% |
| Water-related climate drivers | 29 | Given climate-induced water scarcity and drought conditions, what options are there to conserve or recycle water at the household level for hygiene purposes? | 14 | 94.7% |
| Climate-related hazards | 12 | How does chronic disruption of water and sanitation services due to extreme weather events impact personal hygiene behaviours? | 16 | 93.4% |
| Water-related climate drivers | 40 | What challenges are faced by women and girls concerning menstrual health and hygiene during extreme weather events, periods of climate-induced water scarcity, drought and saltwater intrusion? | 14 | 92.7% |
| Water-related climate drivers | 28 | Does climate-induced water scarcity or drought affect an individual's capability, opportunity or motivation to practice effective hygiene behaviours? | 14 | 90.6% |
| Climate-related hazards | 23 | What support is needed for people with disabilities and their caregivers during extreme weather events to maintain personal hygiene? | 16 | 88.0% |
| Water-related climate drivers | 37 | How will climate change alter the risk of exposure to domestic animals and their waste, and how can this be reduced by hygiene in the domestic environment? | 14 | 86.3% |
| Water-related climate drivers | 26 | Are different approaches needed for hygiene promotion and hygiene behaviour change for populations living in water-scarce or drought conditions? | 14 | 85.4% |
| Climate-related hazards | 3 | Given the microbial and chemical contamination risks of floodwater, what messages should be included in health promotion and hygiene behaviour change campaigns? | 16 | 85.0% |
| Climate-related hazards | 5 | How can coordination and implementation of hygiene-related interventions during and post extreme weather events be improved? | 16 | 84.2% |
| Climate-related hazards | 11 | How does acute disruption of water and sanitation services due to extreme weather events impact personal hygiene behaviours? | 16 | 83.8% |
| Climate-related hazards | 16 | What are the determinants of hygiene behaviours during extreme weather events? | 15 | 83.8% |
| Climate-related hazards | 1 | Do people experiencing homelessness have specific hygiene-related challenges during extreme weather events? | 16 | 83.7% |
| Water-related climate drivers | 41 | What challenges are there in maintaining adequate personal hygiene during extreme weather events, periods of climate-induced water scarcity, drought and saltwater intrusion? | 14 | 83.1% |
| Temperature-related climate drivers | 46 | How do changes in ambient temperature, humidity and precipitation affect food safety and incidence of foodborne illness? | 16 | 82.9% |
| Water-related climate drivers | 33 | How do climate-induced water scarcity, drought or precipitation variability affect the infectious disease burden, and how is this mediated by changes to hygiene practices? | 14 | 82.7% |
| Temperature-related climate drivers | 50 | How will extreme heat affect water collecting practices and the water available for hygiene practices? | 16 | 82.2% |
| Water-related climate drivers | 36 | How does the reduced opportunity to practice personal hygiene (washing, bathing, showering) affect mental health during extreme weather events or periods of climate-induced water scarcity and drought? | 14 | 82.2% |
| Climate-related hazards | 14 | How well are interventions addressing gender-specific hygiene needs during extreme weather events? | 16 | 82.1% |
| Climate-related hazards | 6 | How can individuals, and those that support them, maintain menstrual health during extreme weather events? | 16 | 80.8% |
| Temperature-related climate drivers | 52 | What effect will an increase in ambient temperature and humidity associated with climate change have on pathogen levels, with implications for human health, in water, food, soil, surfaces, and the environment? | 16 | 80.2% |
| Temperature-related climate drivers | 47 | How do extreme heat events affect an individual's use and consumption of water for hygiene practices? | 16 | 79.5% |
| Water-related climate drivers | 35 | How does the effect of climate change on women's and girls' hygiene affect their participation in wider society? | 14 | 78.7% |
| Climate-related hazards | 25 | Which approaches are effective in preparing and restoring the hygiene supply chain to the population during extreme weather events? | 16 | 77.4% |
| Climate-related hazards | 19 | What culturally acceptable products are needed in disaster preparedness kits for personal hygiene? | 16 | 77.2% |
| Climate-related hazards | 13 | How resilient are hygiene services (i.e., facilities, hardware, infrastructure) to the effects of climate change? | 16 | 76.3% |
| Climate-related hazards | 2 | During extreme weather events, how does solid waste contaminate the domestic environment and increase the exposure to infectious diseases and/or chemical pollutants? | 17 | 76.3% |
| Water-related climate drivers | 32 | How do climate-induced water scarcity and drought affect individual's use and consumption of water for hygiene practices? | 14 | 76.0% |
| Climate-related hazards | 9 | How do extreme weather events affect the capability of people with disabilities to maintain personal hygiene? | 16 | 75.5% |
| Water-related climate drivers | 39 | What are the risks of vaginal or reproductive tract infections (RTIs), urinary tract infections (UTIs), fungal infections, bacterial vaginosis, rashes or discomfort due to inadequate menstrual hygiene from climate change events? And does this differ concerning particular climate hazards, such as saltwater intrusion, flood or cyclone waters, or drought, and if so, why? | 14 | 75.5% |
| Temperature-related climate drivers | 45 | Given the food contamination risks resulting from higher temperatures and extreme weather events, what messages should be included in the promotion of safe food hygiene practices? | 16 | 75.3% |
| Climate-related hazards | 15 | What are appropriate channels of communication to populations about hygiene-related health risks before, during and post extreme weather events? | 16 | 73.8% |
| Climate-related hazards | 17 | What are the health risks of using floodwater for personal and domestic hygiene? | 16 | 73.1% |
| Water-related climate drivers | 43 | What will be the effect on skin conditions from inadequate personal hygiene due to climate change? | 14 | 71.5% |
| Climate-related hazards | 10 | How do individuals prioritise hygiene in relation to other needs during extreme weather events? | 16 | 68.4% |
| Climate-related hazards | 22 | What microbiological and chemical contaminants from floods can be mitigated with effective domestic hygiene? | 16 | 67.8% |
| Climate-related hazards | 20 | What hygiene products are needed for post-flood cleaning of households and domestic spaces? | 16 | 65.3% |
| Water-related climate drivers | 30 | How are budgets of national and local stakeholders being allocated for climate-resilient hygiene efforts and services? | 14 | 61.9% |
| Climate-related hazards | 7 | How can the hygiene needs of people with incontinence be addressed during extreme weather events? | 16 | 61.8% |
| Climate-related hazards | 8 | How do domestic hygiene practices during extreme weather events (droughts, heavy precipitation, floods and cyclones) change the risk of mosquito-borne diseases? | 16 | 61.5% |
| Water-related climate drivers | 38 | What are the gender-specific hygiene needs affected by climate change? | 14 | 57.8% |
| Climate-related hazards | 4 | How are private water service providers considering and planning for the potential increase in hygiene needs during extreme weather events? | 16 | 56.5% |
| Temperature-related climate drivers | 44 | Do climate-induced cold spells or freezes affect an individual's capability, opportunity or motivation to practice effective hygiene behaviours? | 16 | 55.6% |
| Water-related climate drivers | 31 | How can effective behaviour change programmes for hygiene-related risks of climate change (e.g., floods, cyclones, climate-change induced water scarcity and drought) best be designed and delivered? | 14 | 52.9% |
| Temperature-related climate drivers | 57 | What is the stability and durability of hygiene products, (e.g., soap, detergents, cleaning products) under higher ambient temperature and humidity throughout the process of manufacturing, transportation and use? | 16 | 51.9% |
| Temperature-related climate drivers | 56 | What is the association between higher ambient temperatures, precipitation variability and the effectiveness of handwashing interventions against diarrhoeal disease? | 16 | 51.2% |
| Temperature-related climate drivers | 49 | How will climate change affect the current Burden of Disease estimates related to hygiene? | 16 | 51.0% |
| Water-related climate drivers | 42 | What is the association between extreme weather events, climate-induced water scarcity, drought or saltwater intrusion with the incidence of neglected tropical diseases (NTDs), and how are these risks mediated by hygiene practices? | 14 | 43.0% |
| Temperature-related climate drivers | 48 | How does climate change affect the food preparation and hygiene practices of individuals, and how does this differ by climate events, such as heatwaves, high humidity, extreme weather events and/or climate-induced water scarcity? | 16 | 37.2% |
| Water-related climate drivers | 34 | How does climate-induced water scarcity affect individual's experience of shame, embarrassment and humiliation due to their lack of opportunity for personal or menstrual hygiene? | 14 | 35.0% |
| Water-related climate drivers | 27 | Do climate-induced water scarcity and droughts change individual preferences for disposable hygiene materials (e.g. nappies, menstrual pads, incontinence pads)? | 14 | 28.1% |
| Temperature-related climate drivers | 54 | What is the association between higher ambient temperatures, precipitation variability and hand-washing behaviours? | 16 | 17.3% |
| Temperature-related climate drivers | 53 | What is the association between higher ambient temperatures, precipitation variability and hand contamination? | 16 | 12.1% |
| Climate-related hazards | 24 | What surfaces are most critical for cleaning during heavy rains and floods to maintain hygiene in the domestic environment? | 16 | 0.0% |

| 5. Eastern Mediterranean Region (EMRO) | | | | |
| --- | --- | --- | --- | --- |
| **Climate Hazards** | **RQ ID #** | **Research Question** | **# of Respondents** | **Scaled Weighted RPS (%)** |
| Climate-related hazards | 21 | What hygiene promotion actions and preparedness strategies are needed before or during extreme weather events to prepare populations for increased risks to health? | 19 | 100.0% |
| Climate-related hazards | 18 | What conditions for vectors and pests (e.g., mosquitoes, mice, cockroaches, and rats) during floods can be mitigated with effective domestic hygiene? | 19 | 88.6% |
| Water-related climate drivers | 32 | How do climate-induced water scarcity and drought affect individual's use and consumption of water for hygiene practices? | 18 | 88.1% |
| Water-related climate drivers | 33 | How do climate-induced water scarcity, drought or precipitation variability affect the infectious disease burden, and how is this mediated by changes to hygiene practices? | 18 | 87.8% |
| Water-related climate drivers | 28 | Does climate-induced water scarcity or drought affect an individual's capability, opportunity or motivation to practice effective hygiene behaviours? | 18 | 86.6% |
| Climate-related hazards | 9 | How do extreme weather events affect the capability of people with disabilities to maintain personal hygiene? | 19 | 86.2% |
| Climate-related hazards | 17 | What are the health risks of using floodwater for personal and domestic hygiene? | 19 | 85.6% |
| Climate-related hazards | 12 | How does chronic disruption of water and sanitation services due to extreme weather events impact personal hygiene behaviours? | 19 | 85.1% |
| Temperature-related climate drivers | 46 | How do changes in ambient temperature, humidity and precipitation affect food safety and incidence of foodborne illness? | 21 | 83.7% |
| Climate-related hazards | 23 | What support is needed for people with disabilities and their caregivers during extreme weather events to maintain personal hygiene? | 19 | 81.8% |
| Temperature-related climate drivers | 55 | What is the association between higher ambient temperatures, precipitation variability and humidity with the incidence of diarrhoeal diseases including cholera? | 20 | 81.5% |
| Water-related climate drivers | 40 | What challenges are faced by women and girls concerning menstrual health and hygiene during extreme weather events, periods of climate-induced water scarcity, drought and saltwater intrusion? | 18 | 77.5% |
| Climate-related hazards | 7 | How can the hygiene needs of people with incontinence be addressed during extreme weather events? | 20 | 77.3% |
| Water-related climate drivers | 39 | What are the risks of vaginal or reproductive tract infections (RTIs), urinary tract infections (UTIs), fungal infections, bacterial vaginosis, rashes or discomfort due to inadequate menstrual hygiene from climate change events? And does this differ concerning particular climate hazards, such as saltwater intrusion, flood or cyclone waters, or drought, and if so, why? | 18 | 76.5% |
| Temperature-related climate drivers | 51 | How will higher ambient temperatures, humidity and precipitation change the contamination of food across the whole food chain, from preparation, processing, storage and consumption? | 20 | 74.9% |
| Temperature-related climate drivers | 47 | How do extreme heat events affect an individual's use and consumption of water for hygiene practices? | 21 | 73.4% |
| Climate-related hazards | 6 | How can individuals, and those that support them, maintain menstrual health during extreme weather events? | 20 | 72.7% |
| Climate-related hazards | 3 | Given the microbial and chemical contamination risks of floodwater, what messages should be included in health promotion and hygiene behaviour change campaigns? | 20 | 71.2% |
| Water-related climate drivers | 35 | How does the effect of climate change on women's and girls' hygiene affect their participation in wider society? | 18 | 70.7% |
| Water-related climate drivers | 41 | What challenges are there in maintaining adequate personal hygiene during extreme weather events, periods of climate-induced water scarcity, drought and saltwater intrusion? | 18 | 70.6% |
| Climate-related hazards | 1 | Do people experiencing homelessness have specific hygiene-related challenges during extreme weather events? | 19 | 70.4% |
| Water-related climate drivers | 29 | Given climate-induced water scarcity and drought conditions, what options are there to conserve or recycle water at the household level for hygiene purposes? | 18 | 70.2% |
| Water-related climate drivers | 30 | How are budgets of national and local stakeholders being allocated for climate-resilient hygiene efforts and services? | 18 | 67.3% |
| Temperature-related climate drivers | 50 | How will extreme heat affect water collecting practices and the water available for hygiene practices? | 21 | 66.0% |
| Water-related climate drivers | 26 | Are different approaches needed for hygiene promotion and hygiene behaviour change for populations living in water-scarce or drought conditions? | 18 | 65.7% |
| Water-related climate drivers | 42 | What is the association between extreme weather events, climate-induced water scarcity, drought or saltwater intrusion with the incidence of neglected tropical diseases (NTDs), and how are these risks mediated by hygiene practices? | 18 | 64.5% |
| Temperature-related climate drivers | 45 | Given the food contamination risks resulting from higher temperatures and extreme weather events, what messages should be included in the promotion of safe food hygiene practices? | 21 | 64.2% |
| Climate-related hazards | 14 | How well are interventions addressing gender-specific hygiene needs during extreme weather events? | 19 | 62.3% |
| Climate-related hazards | 8 | How do domestic hygiene practices during extreme weather events (droughts, heavy precipitation, floods and cyclones) change the risk of mosquito-borne diseases? | 19 | 61.9% |
| Climate-related hazards | 11 | How does acute disruption of water and sanitation services due to extreme weather events impact personal hygiene behaviours? | 19 | 61.9% |
| Climate-related hazards | 2 | During extreme weather events, how does solid waste contaminate the domestic environment and increase the exposure to infectious diseases and/or chemical pollutants? | 20 | 61.4% |
| Water-related climate drivers | 31 | How can effective behaviour change programmes for hygiene-related risks of climate change (e.g., floods, cyclones, climate-change induced water scarcity and drought) best be designed and delivered? | 18 | 61.3% |
| Temperature-related climate drivers | 52 | What effect will an increase in ambient temperature and humidity associated with climate change have on pathogen levels, with implications for human health, in water, food, soil, surfaces, and the environment? | 20 | 59.8% |
| Climate-related hazards | 5 | How can coordination and implementation of hygiene-related interventions during and post extreme weather events be improved? | 20 | 59.3% |
| Water-related climate drivers | 43 | What will be the effect on skin conditions from inadequate personal hygiene due to climate change? | 18 | 58.2% |
| Climate-related hazards | 15 | What are appropriate channels of communication to populations about hygiene-related health risks before, during and post extreme weather events? | 19 | 58.1% |
| Water-related climate drivers | 34 | How does climate-induced water scarcity affect individual's experience of shame, embarrassment and humiliation due to their lack of opportunity for personal or menstrual hygiene? | 18 | 57.8% |
| Climate-related hazards | 13 | How resilient are hygiene services (i.e., facilities, hardware, infrastructure) to the effects of climate change? | 19 | 56.7% |
| Climate-related hazards | 25 | Which approaches are effective in preparing and restoring the hygiene supply chain to the population during extreme weather events? | 19 | 55.8% |
| Water-related climate drivers | 36 | How does the reduced opportunity to practice personal hygiene (washing, bathing, showering) affect mental health during extreme weather events or periods of climate-induced water scarcity and drought? | 18 | 54.5% |
| Climate-related hazards | 10 | How do individuals prioritise hygiene in relation to other needs during extreme weather events? | 19 | 52.6% |
| Temperature-related climate drivers | 49 | How will climate change affect the current Burden of Disease estimates related to hygiene? | 21 | 51.4% |
| Climate-related hazards | 20 | What hygiene products are needed for post-flood cleaning of households and domestic spaces? | 19 | 50.5% |
| Climate-related hazards | 16 | What are the determinants of hygiene behaviours during extreme weather events? | 19 | 50.1% |
| Water-related climate drivers | 37 | How will climate change alter the risk of exposure to domestic animals and their waste, and how can this be reduced by hygiene in the domestic environment? | 18 | 49.9% |
| Water-related climate drivers | 38 | What are the gender-specific hygiene needs affected by climate change? | 18 | 46.9% |
| Climate-related hazards | 4 | How are private water service providers considering and planning for the potential increase in hygiene needs during extreme weather events? | 20 | 46.8% |
| Climate-related hazards | 22 | What microbiological and chemical contaminants from floods can be mitigated with effective domestic hygiene? | 19 | 46.4% |
| Climate-related hazards | 19 | What culturally acceptable products are needed in disaster preparedness kits for personal hygiene? | 19 | 45.6% |
| Water-related climate drivers | 27 | Do climate-induced water scarcity and droughts change individual preferences for disposable hygiene materials (e.g. nappies, menstrual pads, incontinence pads)? | 18 | 40.9% |
| Temperature-related climate drivers | 48 | How does climate change affect the food preparation and hygiene practices of individuals, and how does this differ by climate events, such as heatwaves, high humidity, extreme weather events and/or climate-induced water scarcity? | 21 | 26.2% |
| Temperature-related climate drivers | 44 | Do climate-induced cold spells or freezes affect an individual's capability, opportunity or motivation to practice effective hygiene behaviours? | 21 | 24.4% |
| Temperature-related climate drivers | 56 | What is the association between higher ambient temperatures, precipitation variability and the effectiveness of handwashing interventions against diarrhoeal disease? | 20 | 23.9% |
| Temperature-related climate drivers | 57 | What is the stability and durability of hygiene products, (e.g., soap, detergents, cleaning products) under higher ambient temperature and humidity throughout the process of manufacturing, transportation and use? | 20 | 22.7% |
| Temperature-related climate drivers | 54 | What is the association between higher ambient temperatures, precipitation variability and hand-washing behaviours? | 20 | 10.3% |
| Climate-related hazards | 24 | What surfaces are most critical for cleaning during heavy rains and floods to maintain hygiene in the domestic environment? | 19 | 1.9% |
| Temperature-related climate drivers | 53 | What is the association between higher ambient temperatures, precipitation variability and hand contamination? | 20 | 0.0% |

| 6. Western Pacific Region (WPRO) | | | | |
| --- | --- | --- | --- | --- |
| **Climate Hazards** | **RQ ID #** | **Research Question** | **# of Respondents** | **Scaled Weighted RPS (%)** |
| Water-related climate drivers | 33 | How do climate-induced water scarcity, drought or precipitation variability affect the infectious disease burden, and how is this mediated by changes to hygiene practices? | 14 | 100.0% |
| Water-related climate drivers | 28 | Does climate-induced water scarcity or drought affect an individual's capability, opportunity or motivation to practice effective hygiene behaviours? | 14 | 99.5% |
| Climate-related hazards | 18 | What conditions for vectors and pests (e.g., mosquitoes, mice, cockroaches, and rats) during floods can be mitigated with effective domestic hygiene? | 14 | 95.7% |
| Climate-related hazards | 23 | What support is needed for people with disabilities and their caregivers during extreme weather events to maintain personal hygiene? | 14 | 94.4% |
| Temperature-related climate drivers | 55 | What is the association between higher ambient temperatures, precipitation variability and humidity with the incidence of diarrhoeal diseases including cholera? | 14 | 92.6% |
| Water-related climate drivers | 26 | Are different approaches needed for hygiene promotion and hygiene behaviour change for populations living in water-scarce or drought conditions? | 14 | 91.2% |
| Climate-related hazards | 21 | What hygiene promotion actions and preparedness strategies are needed before or during extreme weather events to prepare populations for increased risks to health? | 14 | 89.0% |
| Water-related climate drivers | 32 | How do climate-induced water scarcity and drought affect individual's use and consumption of water for hygiene practices? | 14 | 86.9% |
| Temperature-related climate drivers | 52 | What effect will an increase in ambient temperature and humidity associated with climate change have on pathogen levels, with implications for human health, in water, food, soil, surfaces, and the environment? | 14 | 85.7% |
| Water-related climate drivers | 38 | What are the gender-specific hygiene needs affected by climate change? | 14 | 85.4% |
| Water-related climate drivers | 40 | What challenges are faced by women and girls concerning menstrual health and hygiene during extreme weather events, periods of climate-induced water scarcity, drought and saltwater intrusion? | 14 | 85.3% |
| Water-related climate drivers | 42 | What is the association between extreme weather events, climate-induced water scarcity, drought or saltwater intrusion with the incidence of neglected tropical diseases (NTDs), and how are these risks mediated by hygiene practices? | 14 | 83.6% |
| Climate-related hazards | 8 | How do domestic hygiene practices during extreme weather events (droughts, heavy precipitation, floods and cyclones) change the risk of mosquito-borne diseases? | 14 | 83.1% |
| Temperature-related climate drivers | 45 | Given the food contamination risks resulting from higher temperatures and extreme weather events, what messages should be included in the promotion of safe food hygiene practices? | 15 | 82.8% |
| Climate-related hazards | 14 | How well are interventions addressing gender-specific hygiene needs during extreme weather events? | 14 | 82.7% |
| Temperature-related climate drivers | 47 | How do extreme heat events affect an individual's use and consumption of water for hygiene practices? | 15 | 82.5% |
| Climate-related hazards | 19 | What culturally acceptable products are needed in disaster preparedness kits for personal hygiene? | 14 | 82.4% |
| Temperature-related climate drivers | 46 | How do changes in ambient temperature, humidity and precipitation affect food safety and incidence of foodborne illness? | 15 | 81.8% |
| Climate-related hazards | 12 | How does chronic disruption of water and sanitation services due to extreme weather events impact personal hygiene behaviours? | 14 | 80.5% |
| Climate-related hazards | 11 | How does acute disruption of water and sanitation services due to extreme weather events impact personal hygiene behaviours? | 14 | 80.3% |
| Climate-related hazards | 9 | How do extreme weather events affect the capability of people with disabilities to maintain personal hygiene? | 14 | 79.7% |
| Temperature-related climate drivers | 51 | How will higher ambient temperatures, humidity and precipitation change the contamination of food across the whole food chain, from preparation, processing, storage and consumption? | 14 | 79.7% |
| Climate-related hazards | 2 | During extreme weather events, how does solid waste contaminate the domestic environment and increase the exposure to infectious diseases and/or chemical pollutants? | 14 | 78.7% |
| Climate-related hazards | 13 | How resilient are hygiene services (i.e., facilities, hardware, infrastructure) to the effects of climate change? | 14 | 76.5% |
| Climate-related hazards | 15 | What are appropriate channels of communication to populations about hygiene-related health risks before, during and post extreme weather events? | 14 | 76.3% |
| Water-related climate drivers | 41 | What challenges are there in maintaining adequate personal hygiene during extreme weather events, periods of climate-induced water scarcity, drought and saltwater intrusion? | 14 | 76.2% |
| Climate-related hazards | 3 | Given the microbial and chemical contamination risks of floodwater, what messages should be included in health promotion and hygiene behaviour change campaigns? | 14 | 75.9% |
| Climate-related hazards | 25 | Which approaches are effective in preparing and restoring the hygiene supply chain to the population during extreme weather events? | 14 | 74.3% |
| Water-related climate drivers | 35 | How does the effect of climate change on women's and girls' hygiene affect their participation in wider society? | 14 | 72.7% |
| Water-related climate drivers | 29 | Given climate-induced water scarcity and drought conditions, what options are there to conserve or recycle water at the household level for hygiene purposes? | 14 | 72.1% |
| Climate-related hazards | 6 | How can individuals, and those that support them, maintain menstrual health during extreme weather events? | 14 | 69.1% |
| Water-related climate drivers | 37 | How will climate change alter the risk of exposure to domestic animals and their waste, and how can this be reduced by hygiene in the domestic environment? | 14 | 67.7% |
| Climate-related hazards | 7 | How can the hygiene needs of people with incontinence be addressed during extreme weather events? | 14 | 67.0% |
| Water-related climate drivers | 31 | How can effective behaviour change programmes for hygiene-related risks of climate change (e.g., floods, cyclones, climate-change induced water scarcity and drought) best be designed and delivered? | 14 | 66.0% |
| Temperature-related climate drivers | 49 | How will climate change affect the current Burden of Disease estimates related to hygiene? | 15 | 64.8% |
| Climate-related hazards | 16 | What are the determinants of hygiene behaviours during extreme weather events? | 13 | 62.8% |
| Climate-related hazards | 10 | How do individuals prioritise hygiene in relation to other needs during extreme weather events? | 14 | 62.0% |
| Temperature-related climate drivers | 50 | How will extreme heat affect water collecting practices and the water available for hygiene practices? | 15 | 61.5% |
| Water-related climate drivers | 39 | What are the risks of vaginal or reproductive tract infections (RTIs), urinary tract infections (UTIs), fungal infections, bacterial vaginosis, rashes or discomfort due to inadequate menstrual hygiene from climate change events? And does this differ concerning particular climate hazards, such as saltwater intrusion, flood or cyclone waters, or drought, and if so, why? | 14 | 58.7% |
| Climate-related hazards | 22 | What microbiological and chemical contaminants from floods can be mitigated with effective domestic hygiene? | 14 | 57.8% |
| Climate-related hazards | 24 | What surfaces are most critical for cleaning during heavy rains and floods to maintain hygiene in the domestic environment? | 14 | 57.8% |
| Climate-related hazards | 17 | What are the health risks of using floodwater for personal and domestic hygiene? | 14 | 55.4% |
| Water-related climate drivers | 36 | How does the reduced opportunity to practice personal hygiene (washing, bathing, showering) affect mental health during extreme weather events or periods of climate-induced water scarcity and drought? | 14 | 55.3% |
| Climate-related hazards | 5 | How can coordination and implementation of hygiene-related interventions during and post extreme weather events be improved? | 14 | 52.9% |
| Temperature-related climate drivers | 48 | How does climate change affect the food preparation and hygiene practices of individuals, and how does this differ by climate events, such as heatwaves, high humidity, extreme weather events and/or climate-induced water scarcity? | 15 | 52.0% |
| Climate-related hazards | 1 | Do people experiencing homelessness have specific hygiene-related challenges during extreme weather events? | 13 | 51.0% |
| Climate-related hazards | 20 | What hygiene products are needed for post-flood cleaning of households and domestic spaces? | 14 | 48.5% |
| Water-related climate drivers | 43 | What will be the effect on skin conditions from inadequate personal hygiene due to climate change? | 14 | 48.5% |
| Climate-related hazards | 4 | How are private water service providers considering and planning for the potential increase in hygiene needs during extreme weather events? | 14 | 48.2% |
| Water-related climate drivers | 27 | Do climate-induced water scarcity and droughts change individual preferences for disposable hygiene materials (e.g. nappies, menstrual pads, incontinence pads)? | 14 | 47.0% |
| Temperature-related climate drivers | 44 | Do climate-induced cold spells or freezes affect an individual's capability, opportunity or motivation to practice effective hygiene behaviours? | 15 | 46.5% |
| Water-related climate drivers | 30 | How are budgets of national and local stakeholders being allocated for climate-resilient hygiene efforts and services? | 14 | 45.9% |
| Water-related climate drivers | 34 | How does climate-induced water scarcity affect individual's experience of shame, embarrassment and humiliation due to their lack of opportunity for personal or menstrual hygiene? | 14 | 29.7% |
| Temperature-related climate drivers | 57 | What is the stability and durability of hygiene products, (e.g., soap, detergents, cleaning products) under higher ambient temperature and humidity throughout the process of manufacturing, transportation and use? | 14 | 28.1% |
| Temperature-related climate drivers | 56 | What is the association between higher ambient temperatures, precipitation variability and the effectiveness of handwashing interventions against diarrhoeal disease? | 14 | 26.9% |
| Temperature-related climate drivers | 54 | What is the association between higher ambient temperatures, precipitation variability and hand-washing behaviours? | 14 | 18.3% |
| Water-related climate drivers | 33 | What is the association between higher ambient temperatures, precipitation variability and hand contamination? | 14 | 0.0% |

| 7. Prefer not to say* | | | | |
| --- | --- | --- | --- | --- |
| **Scaled Weighted RPS Percentages are overlapping due to low frequency of survey takers from this organisation type* | | | | |
| **Climate Hazards** | **RQ ID #** | **Research Question** | **# of Respondents** | **Scaled Weighted RPS (%)** |
| Climate-related hazards | 2 | During extreme weather events, how does solid waste contaminate the domestic environment and increase the exposure to infectious diseases and/or chemical pollutants? | 2 | 100.0% |
| Climate-related hazards | 3 | Given the microbial and chemical contamination risks of floodwater, what messages should be included in health promotion and hygiene behaviour change campaigns? | 2 | 100.0% |
| Climate-related hazards | 7 | How can the hygiene needs of people with incontinence be addressed during extreme weather events? | 2 | 100.0% |
| Climate-related hazards | 15 | What are appropriate channels of communication to populations about hygiene-related health risks before, during and post extreme weather events? | 2 | 100.0% |
| Climate-related hazards | 23 | What support is needed for people with disabilities and their caregivers during extreme weather events to maintain personal hygiene? | 2 | 100.0% |
| Water-related climate drivers | 34 | How does climate-induced water scarcity affect individual's experience of shame, embarrassment and humiliation due to their lack of opportunity for personal or menstrual hygiene? | 2 | 100.0% |
| Water-related climate drivers | 35 | How does the effect of climate change on women's and girls' hygiene affect their participation in wider society? | 2 | 100.0% |
| Water-related climate drivers | 36 | How does the reduced opportunity to practice personal hygiene (washing, bathing, showering) affect mental health during extreme weather events or periods of climate-induced water scarcity and drought? | 2 | 100.0% |
| Temperature-related climate drivers | 49 | How will climate change affect the current Burden of Disease estimates related to hygiene? | 2 | 100.0% |
| Temperature-related climate drivers | 51 | How will higher ambient temperatures, humidity and precipitation change the contamination of food across the whole food chain, from preparation, processing, storage and consumption? | 2 | 100.0% |
| Climate-related hazards | 16 | What are the determinants of hygiene behaviours during extreme weather events? | 2 | 93.4% |
| Climate-related hazards | 17 | What are the health risks of using floodwater for personal and domestic hygiene? | 2 | 93.4% |
| Climate-related hazards | 18 | What conditions for vectors and pests (e.g., mosquitoes, mice, cockroaches, and rats) during floods can be mitigated with effective domestic hygiene? | 2 | 93.4% |
| Climate-related hazards | 21 | What hygiene promotion actions and preparedness strategies are needed before or during extreme weather events to prepare populations for increased risks to health? | 2 | 93.4% |
| Climate-related hazards | 1 | Do people experiencing homelessness have specific hygiene-related challenges during extreme weather events? | 2 | 87.3% |
| Water-related climate drivers | 30 | How are budgets of national and local stakeholders being allocated for climate-resilient hygiene efforts and services? | 2 | 87.3% |
| Water-related climate drivers | 33 | How do climate-induced water scarcity, drought or precipitation variability affect the infectious disease burden, and how is this mediated by changes to hygiene practices? | 2 | 87.3% |
| Water-related climate drivers | 37 | How will climate change alter the risk of exposure to domestic animals and their waste, and how can this be reduced by hygiene in the domestic environment? | 2 | 87.3% |
| Water-related climate drivers | 39 | What are the risks of vaginal or reproductive tract infections (RTIs), urinary tract infections (UTIs), fungal infections, bacterial vaginosis, rashes or discomfort due to inadequate menstrual hygiene from climate change events? And, does this differ concerning particular climate hazards, such as saltwater intrusion, flood or cyclone waters, or drought, and if so, why? | 2 | 87.3% |
| Water-related climate drivers | 40 | What challenges are faced by women and girls concerning menstrual health and hygiene during extreme weather events, periods of climate-induced water scarcity, drought and saltwater intrusion? | 2 | 87.3% |
| Water-related climate drivers | 42 | What is the association between extreme weather events, climate-induced water scarcity, drought or saltwater intrusion with the incidence of neglected tropical diseases (NTDs), and how are these risks mediated by hygiene practices? | 2 | 86.7% |
| Temperature-related climate drivers | 52 | What effect will an increase in ambient temperature and humidity associated with climate change have on pathogen levels, with implications for human health, in water, food, soil, surfaces, and the environment? | 2 | 86.7% |
| Climate-related hazards | 4 | How are private water service providers considering and planning for the potential increase in hygiene needs during extreme weather events? | 2 | 81.0% |
| Water-related climate drivers | 26 | Are different approaches needed for hygiene promotion and hygiene behaviour change for populations living in water-scarce or drought conditions? | 2 | 81.0% |
| Water-related climate drivers | 28 | Does climate-induced water scarcity or drought affect an individual's capability, opportunity or motivation to practice effective hygiene behaviours? | 2 | 81.0% |
| Climate-related hazards | 10 | How do individuals prioritise hygiene in relation to other needs during extreme weather events? | 2 | 75.0% |
| Climate-related hazards | 24 | What surfaces are most critical for cleaning during heavy rains and floods to maintain hygiene in the domestic environment? | 2 | 75.0% |
| Temperature-related climate drivers | 55 | What is the association between higher ambient temperatures, precipitation variability and humidity with the incidence of diarrhoeal diseases including cholera? | 2 | 75.0% |
| Temperature-related climate drivers | 56 | What is the association between higher ambient temperatures, precipitation variability and the effectiveness of handwashing interventions against diarrhoeal disease? | 2 | 75.0% |
| Water-related climate drivers | 27 | Do climate-induced water scarcity and droughts change individual preferences for disposable hygiene materials (e.g. nappies, menstrual pads, incontinence pads)? | 2 | 69.0% |
| Temperature-related climate drivers | 54 | What is the association between higher ambient temperatures, precipitation variability and hand-washing behaviours? | 2 | 68.4% |
| Water-related climate drivers | 32 | How do climate-induced water scarcity and drought affect individual's use and consumption of water for hygiene practices? | 2 | 62.3% |
| Temperature-related climate drivers | 57 | What is the stability and durability of hygiene products, (e.g., soap, detergents, cleaning products) under higher ambient temperature and humidity throughout the process of manufacturing, transportation and use? | 2 | 62.3% |
| Water-related climate drivers | 29 | Given climate-induced water scarcity and drought conditions, what options are there to conserve or recycle water at the household level for hygiene purposes? | 2 | 56.3% |
| Temperature-related climate drivers | 44 | Do climate-induced cold spells or freezes affect an individual's capability, opportunity or motivation to practice effective hygiene behaviours? | 2 | 55.7% |
| Climate-related hazards | 8 | How do domestic hygiene practices during extreme weather events (droughts, heavy precipitation, floods and cyclones) change the risk of mosquito-borne diseases? | 2 | 50.0% |
| Climate-related hazards | 9 | How do extreme weather events affect the capability of people with disabilities to maintain personal hygiene? | 2 | 50.0% |
| Temperature-related climate drivers | 45 | Given the food contamination risks resulting from higher temperatures and extreme weather events, what messages should be included in the promotion of safe food hygiene practices? | 2 | 50.0% |
| Temperature-related climate drivers | 46 | How do changes in ambient temperature, humidity and precipitation affect food safety and incidence of foodborne illness? | 2 | 50.0% |
| Temperature-related climate drivers | 50 | How will extreme heat affect water collecting practices and the water available for hygiene practices? | 2 | 50.0% |
| Temperature-related climate drivers | 53 | What is the association between higher ambient temperatures, precipitation variability and hand contamination? | 2 | 50.0% |
| Climate-related hazards | 14 | How well are interventions addressing gender-specific hygiene needs during extreme weather events? | 2 | 49.7% |
| Climate-related hazards | 22 | What microbiological and chemical contaminants from floods can be mitigated with effective domestic hygiene? | 2 | 44.0% |
| Climate-related hazards | 6 | How can individuals, and those that support them, maintain menstrual health during extreme weather events? | 2 | 43.4% |
| Water-related climate drivers | 31 | How can effective behaviour change programmes for hygiene-related risks of climate change (e.g., floods, cyclones, climate-change induced water scarcity and drought) best be designed and delivered? | 2 | 43.4% |
| Temperature-related climate drivers | 48 | How does climate change affect the food preparation and hygiene practices of individuals, and how does this differ by climate events, such as heatwaves, high humidity, extreme weather events and/or climate-induced water scarcity? | 2 | 43.4% |
| Climate-related hazards | 11 | How does acute disruption of water and sanitation services due to extreme weather events impact personal hygiene behaviours? | 2 | 37.3% |
| Climate-related hazards | 13 | How resilient are hygiene services (i.e., facilities, hardware, infrastructure) to the effects of climate change? | 2 | 37.3% |
| Water-related climate drivers | 38 | What are the gender-specific hygiene needs affected by climate change? | 2 | 37.3% |
| Climate-related hazards | 19 | What culturally acceptable products are needed in disaster preparedness kits for personal hygiene? | 2 | 37.0% |
| Temperature-related climate drivers | 47 | How do extreme heat events affect an individual's use and consumption of water for hygiene practices? | 2 | 31.3% |
| Climate-related hazards | 5 | How can coordination and implementation of hygiene-related interventions during and post extreme weather events be improved? | 2 | 31.0% |
| Climate-related hazards | 12 | How does chronic disruption of water and sanitation services due to extreme weather events impact personal hygiene behaviours? | 2 | 31.0% |
| Water-related climate drivers | 41 | What challenges are there in maintaining adequate personal hygiene during extreme weather events, periods of climate-induced water scarcity, drought and saltwater intrusion? | 2 | 31.0% |
| Climate-related hazards | 25 | Which approaches are effective in preparing and restoring the hygiene supply chain to the population during extreme weather events? | 2 | 12.7% |
| Climate-related hazards | 20 | What hygiene products are needed for post-flood cleaning of households and domestic spaces? | 2 | 0.0% |
| Water-related climate drivers | 43 | What will be the effect on skin conditions from inadequate personal hygiene due to climate change? | 2 | 0.0% |

| 8. Global | | | | |
| --- | --- | --- | --- | --- |
| **Climate Hazards** | **RQ ID #** | **Research Question** | **# of Respondents** | **Scaled Weighted RPS (%)** |
| Climate-related hazards | 9 | How do extreme weather events affect the capability of people with disabilities to maintain personal hygiene? | 50 | 100.0% |
| Climate-related hazards | 21 | What hygiene promotion actions and preparedness strategies are needed before or during extreme weather events to prepare populations for increased risks to health? | 49 | 92.9% |
| Water-related climate drivers | 32 | How do climate-induced water scarcity and drought affect individual's use and consumption of water for hygiene practices? | 52 | 92.9% |
| Climate-related hazards | 2 | During extreme weather events, how does solid waste contaminate the domestic environment and increase the exposure to infectious diseases and/or chemical pollutants? | 51 | 92.8% |
| Water-related climate drivers | 33 | How do climate-induced water scarcity, drought or precipitation variability affect the infectious disease burden, and how is this mediated by changes to hygiene practices? | 51 | 91.2% |
| Climate-related hazards | 23 | What support is needed for people with disabilities and their caregivers during extreme weather events to maintain personal hygiene? | 49 | 88.8% |
| Temperature-related climate drivers | 47 | How do extreme heat events affect an individual's use and consumption of water for hygiene practices? | 53 | 88.6% |
| Temperature-related climate drivers | 55 | What is the association between higher ambient temperatures, precipitation variability and humidity with the incidence of diarrhoeal diseases including cholera? | 51 | 87.7% |
| Water-related climate drivers | 42 | What is the association between extreme weather events, climate-induced water scarcity, drought or saltwater intrusion with the incidence of neglected tropical diseases (NTDs), and how are these risks mediated by hygiene practices? | 51 | 86.5% |
| Water-related climate drivers | 28 | Does climate-induced water scarcity or drought affect an individual's capability, opportunity or motivation to practice effective hygiene behaviours? | 52 | 86.0% |
| Climate-related hazards | 16 | What are the determinants of hygiene behaviours during extreme weather events? | 49 | 84.1% |
| Temperature-related climate drivers | 50 | How will extreme heat affect water collecting practices and the water available for hygiene practices? | 53 | 81.9% |
| Water-related climate drivers | 26 | Are different approaches needed for hygiene promotion and hygiene behaviour change for populations living in water-scarce or drought conditions? | 52 | 81.8% |
| Water-related climate drivers | 41 | What challenges are there in maintaining adequate personal hygiene during extreme weather events, periods of climate-induced water scarcity, drought and saltwater intrusion? | 51 | 81.8% |
| Climate-related hazards | 12 | How does chronic disruption of water and sanitation services due to extreme weather events impact personal hygiene behaviours? | 50 | 81.8% |
| Temperature-related climate drivers | 52 | What effect will an increase in ambient temperature and humidity associated with climate change have on pathogen levels, with implications for human health, in water, food, soil, surfaces, and the environment? | 51 | 80.6% |
| Climate-related hazards | 3 | Given the microbial and chemical contamination risks of floodwater, what messages should be included in health promotion and hygiene behaviour change campaigns? | 51 | 80.2% |
| Climate-related hazards | 6 | How can individuals, and those that support them, maintain menstrual health during extreme weather events? | 51 | 80.0% |
| Climate-related hazards | 5 | How can coordination and implementation of hygiene-related interventions during and post extreme weather events be improved? | 51 | 78.6% |
| Climate-related hazards | 11 | How does acute disruption of water and sanitation services due to extreme weather events impact personal hygiene behaviours? | 50 | 77.8% |
| Climate-related hazards | 19 | What culturally acceptable products are needed in disaster preparedness kits for personal hygiene? | 49 | 77.1% |
| Climate-related hazards | 14 | How well are interventions addressing gender-specific hygiene needs during extreme weather events? | 50 | 77.0% |
| Water-related climate drivers | 31 | How can effective behaviour change programmes for hygiene-related risks of climate change (e.g., floods, cyclones, climate-change induced water scarcity and drought) best be designed and delivered? | 52 | 76.7% |
| Temperature-related climate drivers | 45 | Given the food contamination risks resulting from higher temperatures and extreme weather events, what messages should be included in the promotion of safe food hygiene practices? | 53 | 76.3% |
| Climate-related hazards | 13 | How resilient are hygiene services (i.e., facilities, hardware, infrastructure) to the effects of climate change? | 50 | 75.7% |
| Water-related climate drivers | 40 | What challenges are faced by women and girls concerning menstrual health and hygiene during extreme weather events, periods of climate-induced water scarcity, drought and saltwater intrusion? | 51 | 72.1% |
| Climate-related hazards | 18 | What conditions for vectors and pests (e.g., mosquitoes, mice, cockroaches, and rats) during floods can be mitigated with effective domestic hygiene? | 49 | 72.0% |
| Temperature-related climate drivers | 46 | How do changes in ambient temperature, humidity and precipitation affect food safety and incidence of foodborne illness? | 53 | 71.5% |
| Climate-related hazards | 1 | Do people experiencing homelessness have specific hygiene-related challenges during extreme weather events? | 51 | 67.6% |
| Climate-related hazards | 8 | How do domestic hygiene practices during extreme weather events (droughts, heavy precipitation, floods and cyclones) change the risk of mosquito-borne diseases? | 50 | 67.4% |
| Water-related climate drivers | 39 | What are the risks of vaginal or reproductive tract infections (RTIs), urinary tract infections (UTIs), fungal infections, bacterial vaginosis, rashes or discomfort due to inadequate menstrual hygiene from climate change events? And does this differ concerning particular climate hazards, such as saltwater intrusion, flood or cyclone waters, or drought, and if so, why? | 51 | 66.3% |
| Climate-related hazards | 22 | What microbiological and chemical contaminants from floods can be mitigated with effective domestic hygiene? | 49 | 66.3% |
| Temperature-related climate drivers | 49 | How will climate change affect the current Burden of Disease estimates related to hygiene? | 53 | 65.5% |
| Climate-related hazards | 7 | How can the hygiene needs of people with incontinence be addressed during extreme weather events? | 51 | 64.8% |
| Temperature-related climate drivers | 51 | How will higher ambient temperatures, humidity and precipitation change the contamination of food across the whole food chain, from preparation, processing, storage and consumption? | 51 | 63.7% |
| Climate-related hazards | 15 | What are appropriate channels of communication to populations about hygiene-related health risks before, during and post extreme weather events? | 50 | 63.2% |
| Water-related climate drivers | 29 | Given climate-induced water scarcity and drought conditions, what options are there to conserve or recycle water at the household level for hygiene purposes? | 52 | 63.0% |
| Water-related climate drivers | 36 | How does the reduced opportunity to practice personal hygiene (washing, bathing, showering) affect mental health during extreme weather events or periods of climate-induced water scarcity and drought? | 51 | 61.6% |
| Climate-related hazards | 25 | Which approaches are effective in preparing and restoring the hygiene supply chain to the population during extreme weather events? | 49 | 58.6% |
| Water-related climate drivers | 38 | What are the gender-specific hygiene needs affected by climate change? | 51 | 57.6% |
| Climate-related hazards | 10 | How do individuals prioritise hygiene in relation to other needs during extreme weather events? | 50 | 57.4% |
| Water-related climate drivers | 35 | How does the effect of climate change on women's and girls' hygiene affect their participation in wider society? | 51 | 57.2% |
| Climate-related hazards | 4 | How are private water service providers considering and planning for the potential increase in hygiene needs during extreme weather events? | 51 | 54.5% |
| Temperature-related climate drivers | 48 | How does climate change affect the food preparation and hygiene practices of individuals, and how does this differ by climate events, such as heatwaves, high humidity, extreme weather events and/or climate-induced water scarcity? | 53 | 53.1% |
| Climate-related hazards | 20 | What hygiene products are needed for post-flood cleaning of households and domestic spaces? | 49 | 51.3% |
| Water-related climate drivers | 27 | Do climate-induced water scarcity and droughts change individual preferences for disposable hygiene materials (e.g. nappies, menstrual pads, incontinence pads)? | 52 | 48.4% |
| Climate-related hazards | 17 | What are the health risks of using floodwater for personal and domestic hygiene? | 49 | 47.0% |
| Water-related climate drivers | 43 | What will be the effect on skin conditions from inadequate personal hygiene due to climate change? | 51 | 46.1% |
| Water-related climate drivers | 37 | How will climate change alter the risk of exposure to domestic animals and their waste, and how can this be reduced by hygiene in the domestic environment? | 51 | 45.1% |
| Water-related climate drivers | 34 | How does climate-induced water scarcity affect individual's experience of shame, embarrassment and humiliation due to their lack of opportunity for personal or menstrual hygiene? | 51 | 41.5% |
| Temperature-related climate drivers | 44 | Do climate-induced cold spells or freezes affect an individual's capability, opportunity or motivation to practice effective hygiene behaviours? | 53 | 38.2% |
| Water-related climate drivers | 30 | How are budgets of national and local stakeholders being allocated for climate-resilient hygiene efforts and services? | 52 | 37.8% |
| Temperature-related climate drivers | 56 | What is the association between higher ambient temperatures, precipitation variability and the effectiveness of handwashing interventions against diarrhoeal disease? | 51 | 34.1% |
| Climate-related hazards | 24 | What surfaces are most critical for cleaning during heavy rains and floods to maintain hygiene in the domestic environment? | 49 | 29.2% |
| Temperature-related climate drivers | 57 | What is the stability and durability of hygiene products, (e.g., soap, detergents, cleaning products) under higher ambient temperature and humidity throughout the process of manufacturing, transportation and use? | 51 | 25.3% |
| Temperature-related climate drivers | 54 | What is the association between higher ambient temperatures, precipitation variability and hand-washing behaviours? | 51 | 3.0% |
| Temperature-related climate drivers | 53 | What is the association between higher ambient temperatures, precipitation variability and hand contamination? | 51 | 0.0% |
